# The association between socioeconomic status and pandemic influenza: systematic review and meta-analysis

**DOI:** 10.1101/2020.12.09.20246496

**Authors:** Svenn-Erik Mamelund, Clare Shelley-Egan, Ole Rogeberg

## Abstract

**Background:** The objective was to document whether and to what extent there was an association between socioeconomic status (SES) and disease outcomes in the last five influenza pandemics.

**Methods/Principle Findings:** The review included studies published in English, Danish, Norwegian and Swedish. Records were identified through systematic literature searches in six databases. Results are summarized narratively and using meta-analytic strategies. We found studies only for the 1918 and 2009 pandemics. Of 14 studies on the 2009 pandemic including data on both medical and social risk factors, after controlling for medical risk factors 8 demonstrated independent impact of SES. A random effect analysis of 46 estimates from 35 studies found a pooled mean odds ratio of 1.4 (95% CI: 1.2 – 1.7), comparing the lowest to the highest SES, but with substantial effect heterogeneity across studies –reflecting differences in outcome measures and definitions of case and control samples. Analyses by pandemic period (1918 or 2009) and by level of SES measure (individual or ecological) indicate no differences along these dimensions. Studies using healthy controls tend to find low SES associated with worse influenza outcome, and studies using infected controls find low SES associated with more severe outcomes. Studies comparing severe outcomes (ICU or death) to hospital admissions are few but indicate no clear association. Studies with more unusual comparisons (e.g., pandemic vs seasonal influenza, seasonal influenza vs other patient groups) report no or negative associations.

**Conclusions/Significance:** Results show that social risk factors help to explain pandemic outcomes in 1918 and in 2009 although the mechanisms and types of social vulnerabilities leading to disparities in outcomes may differ over time. Studies of the 2009 pandemic also showed that social vulnerability could not always be explained by medical risk factors. To prepare for future pandemics, we must consider social along with medical vulnerability.

The protocol for this study has been registered in PROSPERO (ref. no 87922) and has been published (1).

## Introduction

It used to be believed that pandemic and infectious disease risks are the same for all, irrespective of social background or socioeconomic status (SES). But when 61-year old superstar Madonna shared this belief on Instagram on the 23^rd^ of March 2020, calling COVID-19 “the great equalizer” from a milky bath sprinkled with rose-petals (2), fans and others quickly pointed to the disproportionate pandemic burden and suffering of the poor. Their criticism is supported by a number of studies showing that certain indigenous people, people of colour, immigrants and the poor experienced disproportionate harm from COVID-19 as measured by infection rates, hospitalizations, the need for intensive care unit treatment, and death (3-6).

The idea that outcomes from infectious disease pandemics are socially neutral has an old history among lay people, researchers and policy makers responsible for pandemic preparedness plans. Literature on SES and 1918 influenza outcomes published by social historians between 1970 and 1990 argued that the disease was so highly transmissible that everybody was equally affected (7-11), pointing to anecdotal evidence such as the president and King of Spain falling ill and the Swedish Prince Erik dying at age 29(12). The studies typically lacked empirical analysis interpreting high quality data within statistical models, however. Empirical studies appearing from the mid-2000s often reported evidence inconsistent with the socially neutral hypothesis: SES seemed to be linked to exposure, susceptibility and access to care, and SES indicators were statistically associated with mortality (13-15). Although several studies of the 2009 pandemic also found SES associated with various pandemic outcomes (16-18), this social inequality in risk is still ignored in international pandemic preparedness plans (19). A systematic assessment of the evidence for such risk inequalities has been lacking, however, apart from a systematic review and meta-analysis of how the risk of 2009 influenza pandemic outcomes differ for disadvantaged populations (mainly indigenous people) (20).

In this paper, we present the first systematic review and meta-analysis on the association between SES and disease outcomes in the last 5 influenza pandemics. The objective was to document whether and to what extent there was an association between indicators of socioeconomic status (e.g. income, education) and pandemic outcomes (infection, hospitalizations, mortality) in the last five influenza pandemics (1889, 1918, 1957, 1968, 2009). In terms of PICOS criteria, the Population (P) consists of groups defined by socioeconomic status, the intervention (I) or exposure or risk factor is pandemic influenza, the comparison (C) or alternative interventions is not relevant, while the outcomes (O) are morbidity, hospitalization, or death associated with influenza pandemics. All types of study designs were considered (S). As described in our pre-registered analysis plan, we hypothesized that the association between SES and pandemic outcomes would increase with outcome severity, as higher income and SES tend to be associated with access to resources and protective factors that reduce the risk of progression to more severe outcomes.

Our review identified studies on the 1918 and 2009 pandemics only, with evidence of a social gradient in the disease burden of both these pandemics. Associations with SES were statistically significant in 8 of the 14 studies on the 2009 pandemic that adjusted estimates for medical risk factors, indicating that both sets of risk factors are needed to understand pandemic disease severity. We did not find support for the hypothesis that social risk factors were more important for severe than for less severe outcomes.

## Materials and Methods

### Bibliographic database search

A systematic search of Medline, Embase, Cinahl, SocIndex, Scopus and Web of Science was performed to identify all relevant articles published on socioeconomic factors and pandemic influenza. The strategy for the literature search was developed by two information specialists in cooperation with the research group, starting 5 October 2017. Several pilot searches were conducted in Web of Science and Medline respectively, on 12 and 19 October 2017, to ensure a sensitive search. The search strategy combined relevant terms, both controlled vocabulary terms (i.e. MeSH) and text words. The main search strategy used in Medline is available in PROSPERO 87922 and in the appendix, and the final search was carried out on 17 November 2017. The strategy was modified to fit the other databases listed above. To generate manageable results, restrictions on language (English, Danish, Norwegian and Swedish) and publication type (article/research article) were added to the searches in the other databases. The searches in Medline and Embase were performed without publication type restrictions. The search strategy was peer-reviewed by a third information specialist using a structured tool based on the PRESS-framework (21). Reference lists of relevant known studies were also screened and experts in the field consulted in order to identify other additional sources. Finally, we also contacted authors of published studies to ask for relevant data not presented in the papers or in appendices. However, we did not get any responses that made it into the paper and our analysis

### Inclusion criteria for title and abstract screening

After adding all identified records to an Endnote library and removing duplicates, the remaining results were imported to the program Covidence. Here, additional duplicates were removed. Each article’s title and abstract were screened by two of the authors (SEM and CSE), according to the selection criteria. After screening of titles and abstracts, we added full-text versions of articles in Covidence. Divergences in the inclusion of studies were re-assessed by the same researchers until consensus was reached in terms of inclusion or exclusion. The criteria for inclusion were:

1. The study period 1889-2009 includes the five pandemics in 1889, 1918, 1957, 1968 and 2009
2. Studies looking at the association between SES and pandemic outcome (morbidity, severe disease and mortality). SES was captured by key words such as education, income, occupational social class etc. (see search history for more examples). Morbidity was captured by key words such as infection rates, transmission rates, lab confirmed influenza, flu like illness, and influenza like illness (ILI). Severe disease was captured by key words such as disease severity, critical illness, critical disease, severe illness, severe disease, hospitalization, patient admission, hospital admission, intensive care unit (ICU) admission, and ICU treatment. Mortality was captured by key words such as fatal outcome, fatal illness, fatal disease, fatality, lethal outcome, lethal illness, lethal disease, terminal outcome, terminal illness, terminal disease, lethality, death, death rate, and mortality rate. All of these key-words were used in both pilots and the final search as described above. The search strategy also covered studies of ethnic and disadvantaged populations, as some of these included covariates for socioeconomic confounders that fell within our inclusion criteria.
3. Studies covering both seasonal and pandemic influenza distinguishing between non-pandemic and pandemic years.
4. Studies covering all regions/countries, type of studies (interventional, observational, etc.) and populations (age, gender, pregnant women, soldiers etc.).

### Exclusion criteria for title and abstract screening

The following criteria excluded studies from the systematic literature review:

1. Studies on pandemic diseases other than influenza
2. Studies on seasonal influenza only
3. Studies on both seasonal and pandemic influenza that *<u>did not</u>* distinguish between non-pandemic and pandemic years
4. Studies on influenza vaccine uptake, attitudes towards influenza vaccination and compliance with (non)pharmaceutical interventions during influenza pandemics
5. Case studies or qualitative studies on the associations between socioeconomic factors and pandemic outcomes
6. Studies on social justice and pandemic influenza
7. Studies of pandemic influenza preparedness plans
8. Studies on ethnic and disadvantaged minorities that *<u>did not</u>* report controls for socioeconomic confounders

### Data selection and extraction

We drafted a data abstraction form, pilot tested it and modified it, where necessary. Two reviewers (SEM and CSE) independently extracted data from all included studies. Any disagreements were resolved via discussion or by involving a third reviewer for arbitration. 1-5 and 6 below were entered into separate spreadsheets for each article. The following information was extracted:

1. Article info
  a. First author
  b. Year published
  c. Journal
2. Data sample
  a. Country or region of analysis
  b. Pandemic years (1889, 1918, 1957, 1968, 2009)
  c. Sample inclusion criteria – i.e. characteristics of sample/population (civilian, military, gender, pregnant, age-group/median/average age, patient group etc).
  d. Sample size
  e. Unit of analysis (individuals, households, regions, hospitals etc)
  f. Data aggregation level (observations of individual units, aggregated units, etc.). e.g., if hospitals are the unit of analysis, does the data used occur at the hospital level or is it pooled across hospitals?
  g. Source of outcome data, e.g., census, routine notification data (e.g. influenza cases reported to a doctor), survey data, register data
    i. If survey or population data had incomplete coverage
      1. Response rate/coverage
      2. Representativity: Is the sample shown to be representative for the population? i.e. has a non-response analysis been carried out?
3. Outcome variable - Pandemic outcome (a. morbidity, b. hospitalization, c. mortality)
  a. Definition of morbidity: influenza-like illness (ILI), Lab-confirmed Infection rates (PCR), transmission rates (reproduction number, R0), immunity/antibodies towards influenza (HI titer above a certain threshold) due to exposure to the disease and not vaccination
  b. Definition of hospitalization; Hospitalized inpatients with (PCR) or without confirmed influenza; patients admitted to intensive care unit (ICU) or not; mechanically ventilated patients (“lung machines”) or not; inpatients vs outpatients
  c. Definition of cause of mortality: Influenza and pneumonia (PI), excess mortality (PI, all causes of death etc.), respiratory diseases, pneumonia etc.
4. Baseline outcomes (control type), i.e. what was the control group or baseline outcome comparison? (general healthy population, infected patients, the hospitalized, patients with lab-confirmed seasonal influenza)
5. Independent variables of interest – relating to SES
  a. Type of SES indices (education, income, crowding, density, deprivation index, unemployment, occupational social class, poverty status, % below poverty level)
  b. Definition or brief descriptive text on SES indices (e.g., if based on a specific type of poverty index etc.)
6. Statistical methodology
  a. Design of study (cross sectional, longitudinal, case-control, cohort studies)
  b. Estimation technique (Cross tables, correlation analysis, OLS, Poisson regression, Logistic regression, Cox regressions, GEE regressions, GLMM models etc.)
  c. Control variables included (e.g. age, gender, marital status, pre-existing disease, health behavior etc.) in light of sample restrictions (e.g. for pregnant women, sex is not among the controls)
  d. Reference categories with which all point estimates are compared
7. Results reported (separate spreadsheet)

### Data Synthesis

Our narrative review includes a table of the study characteristics of the included studies, such as study authors and year, pandemic years, study region (region/country/hospital), sample inclusion criteria, sample size, unit of outcomes, data aggregation level, data sources and type, outcomes, baseline outcomes, SES measure, design, statistical techniques, controls and whether the study estimates are used in the meta analysis and whether SES is an independent predictor. The quantitative part of the study pools results across individual studies using meta-analytic methods. Such methods pool the evidence reported from different studies, weighting each study by its precision.

The simplest meta-analytic model (“fixed effect”) is appropriate when several studies estimate the exact same parameter, making random sampling variation the only source of variation in estimates. This is unlikely to be appropriate in our context, where studies assess the associations between SES indicators and medical outcomes using different indicators of SES and flu outcomes in data from different countries and time periods that allow for different levels of confounder control, etc. The differences in the underlying associations studied can be viewed as a form of *effect heterogeneity*, implying that the studies would report different estimates even if sampling variation could be removed. Since the estimated associations are related, however, we can estimate the *distribution* of these underlying associations using a “random effects” model. And finally, we may take this a step further by exploring whether study-level covariates (e.g., country, period, SES-indicator) are associated with particularly high or low estimates.

We searched the identified studies in our meta-review for quantitative estimates of associations between SES indicators and influenza related outcomes. The resulting estimates were assessed for inclusion in the meta-analysis, and included if they could be expressed as an odds-ratio or relative risk for low versus high socioeconomic status. This implied that estimates had to include an indicator of socioeconomic status at the individual or ecological level, and had to allow for an estimate of how the incidence or prevalence of some flu related outcome varied by levels of this indicator. Where studies included estimates for distinct data subsamples (different age groups, periods), single estimates pooling all data were preferred if available. If not, the separate estimates were all included. For some studies, multiple estimates were also extracted if they performed different comparisons (e.g., risk of infection, and risk of hospitalization given infection). We also collected study level factors indicating the pandemic period (1918 vs 2009), country/region, and whether the study estimate involved an odds ratio or a relative risk or rate. The specific studies included and all judgments and adjustments concerning inclusion and adjustments of reported numbers are detailed in the supporting materials.

Relative to the pre-analysis plan, the ambitions of the quantitative analysis and quality assessments (using e.g. NOS(22)) were scaled back given the large heterogeneity across the studies included (see Table 1). The pre-analysis plan specified three types of analysis (1). The first, a standard random effect analysis with subsample analyses, was conducted as planned using the «REML» algorithm in the Metafor meta-analytic package for R (23). The second, a PET-PEESE analysis testing and adjusting for publication bias, was found unsuitable given the large effect heterogeneity (24). The third, a Bayesian model to assess “dose-response” effects and assess how estimates vary with study-level indicators and the type of comparisons made, is included in a simplified version without the “dose-response” element.

**Table 1.**
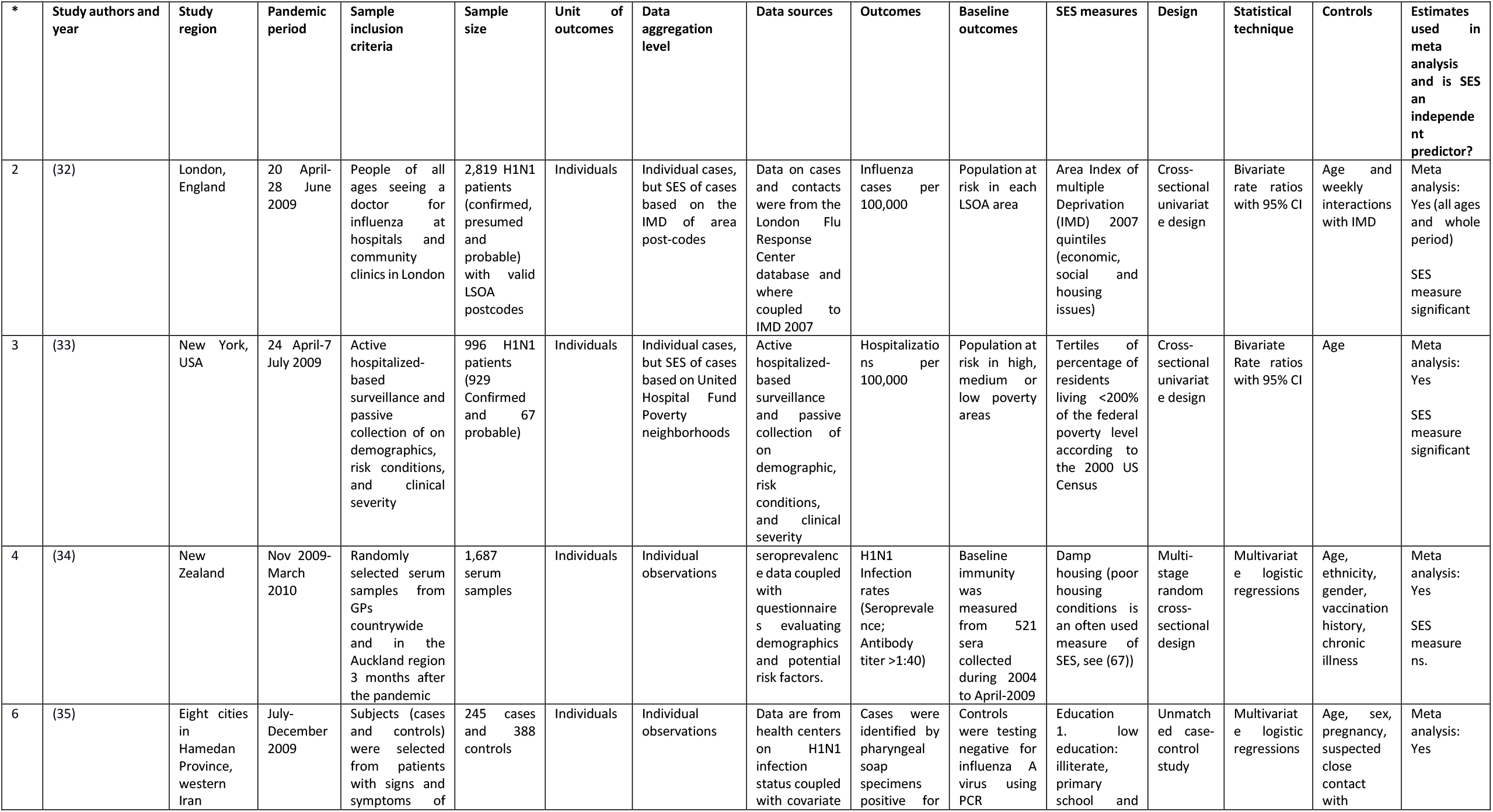
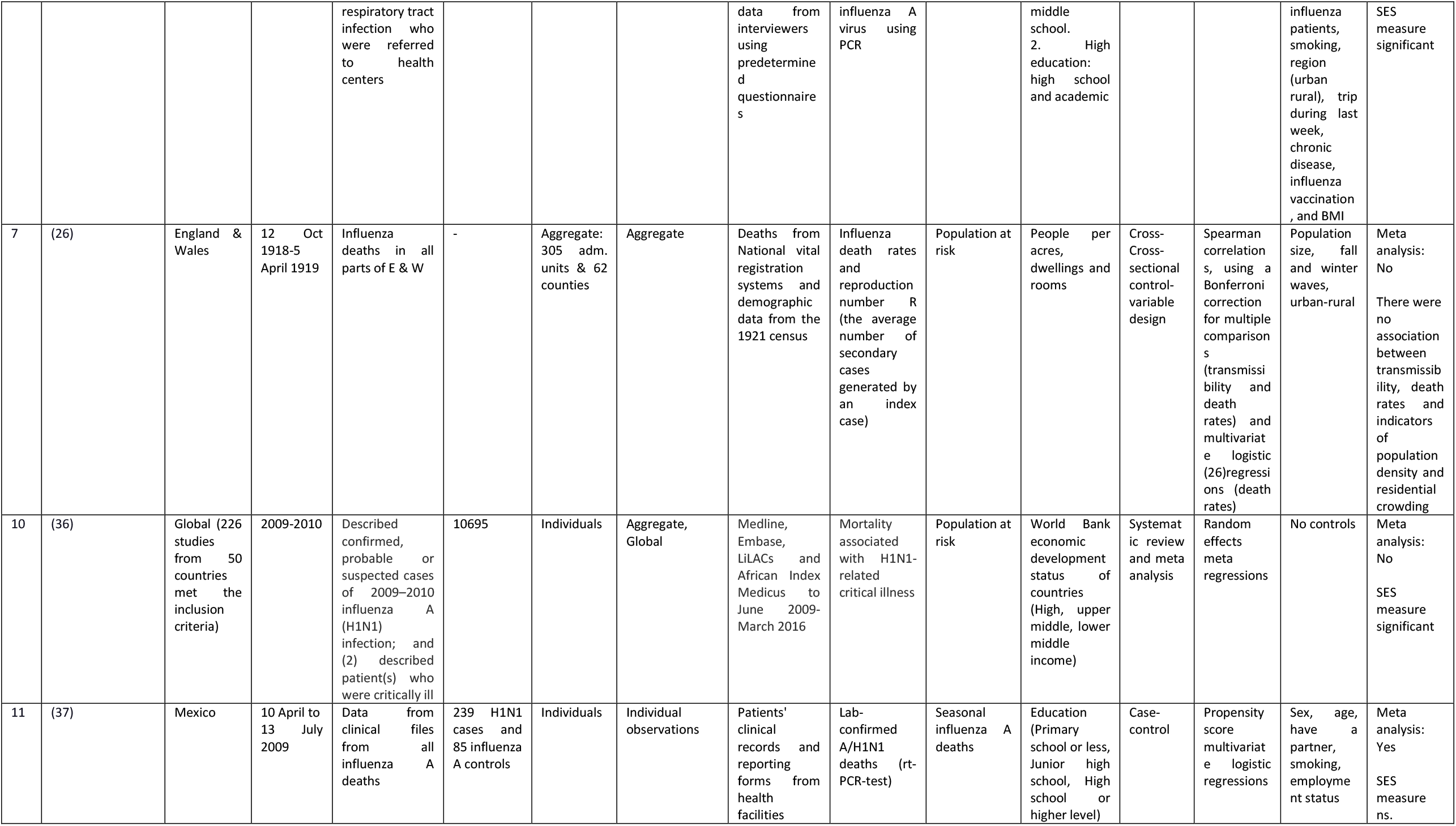
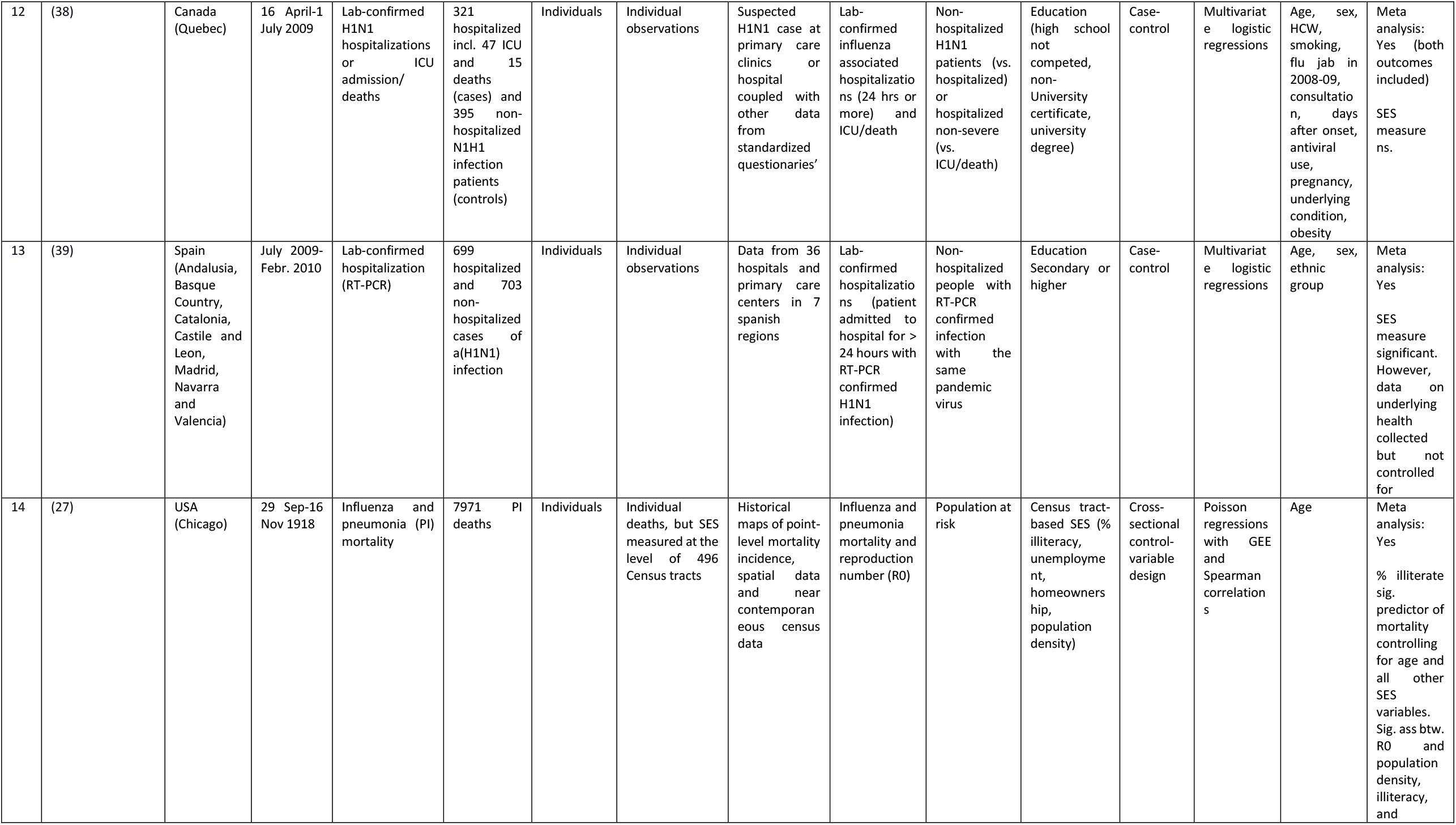
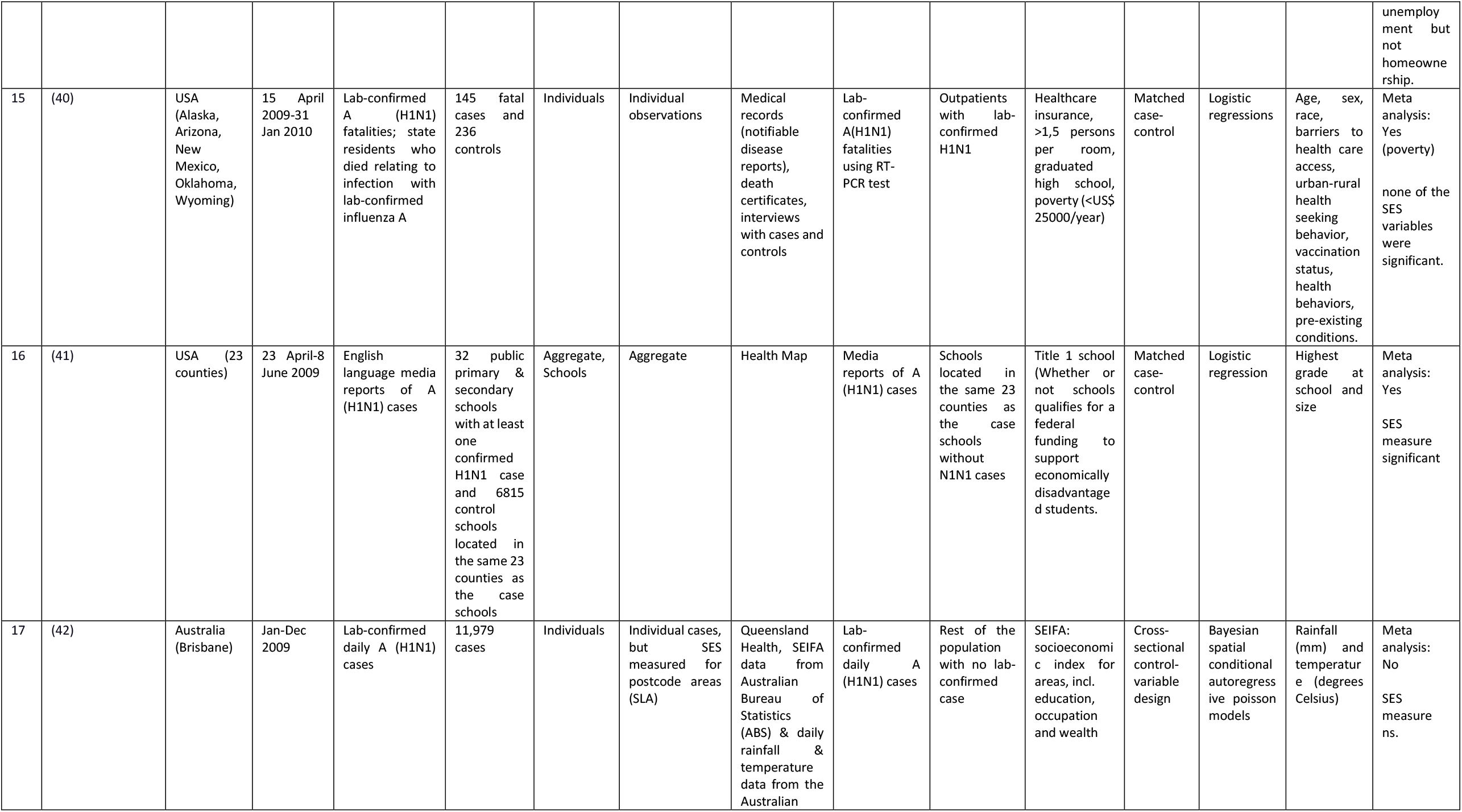
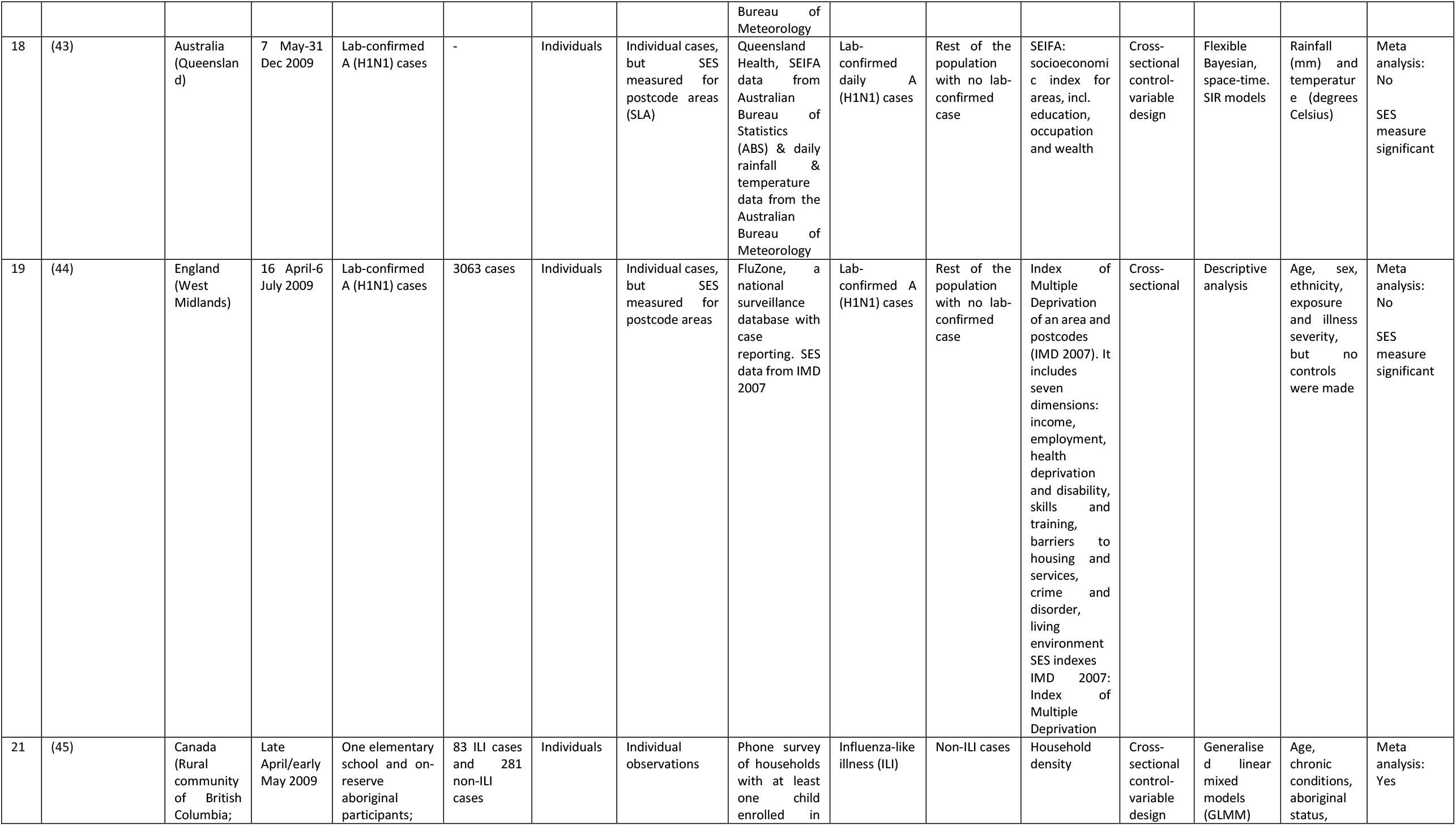
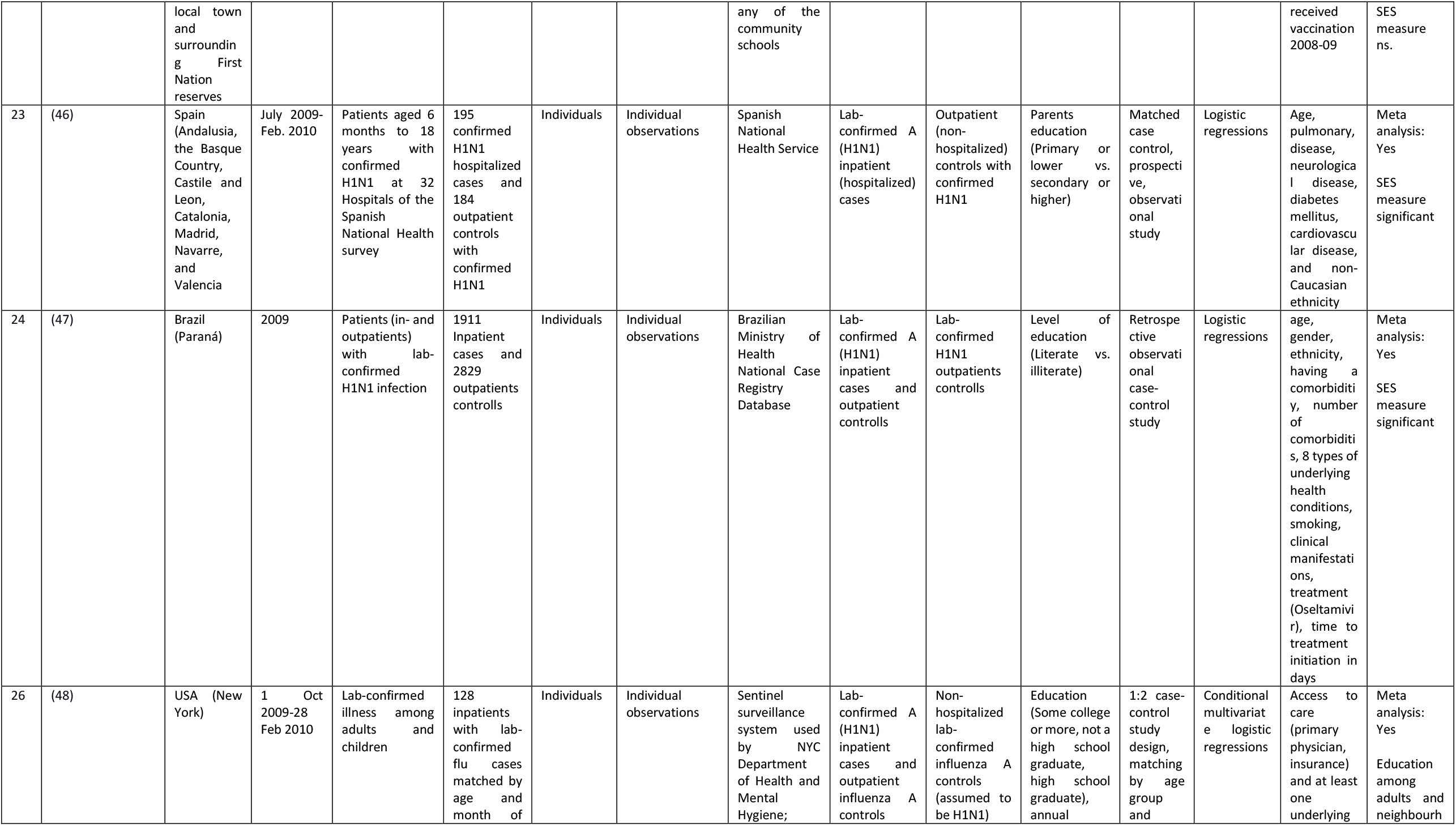
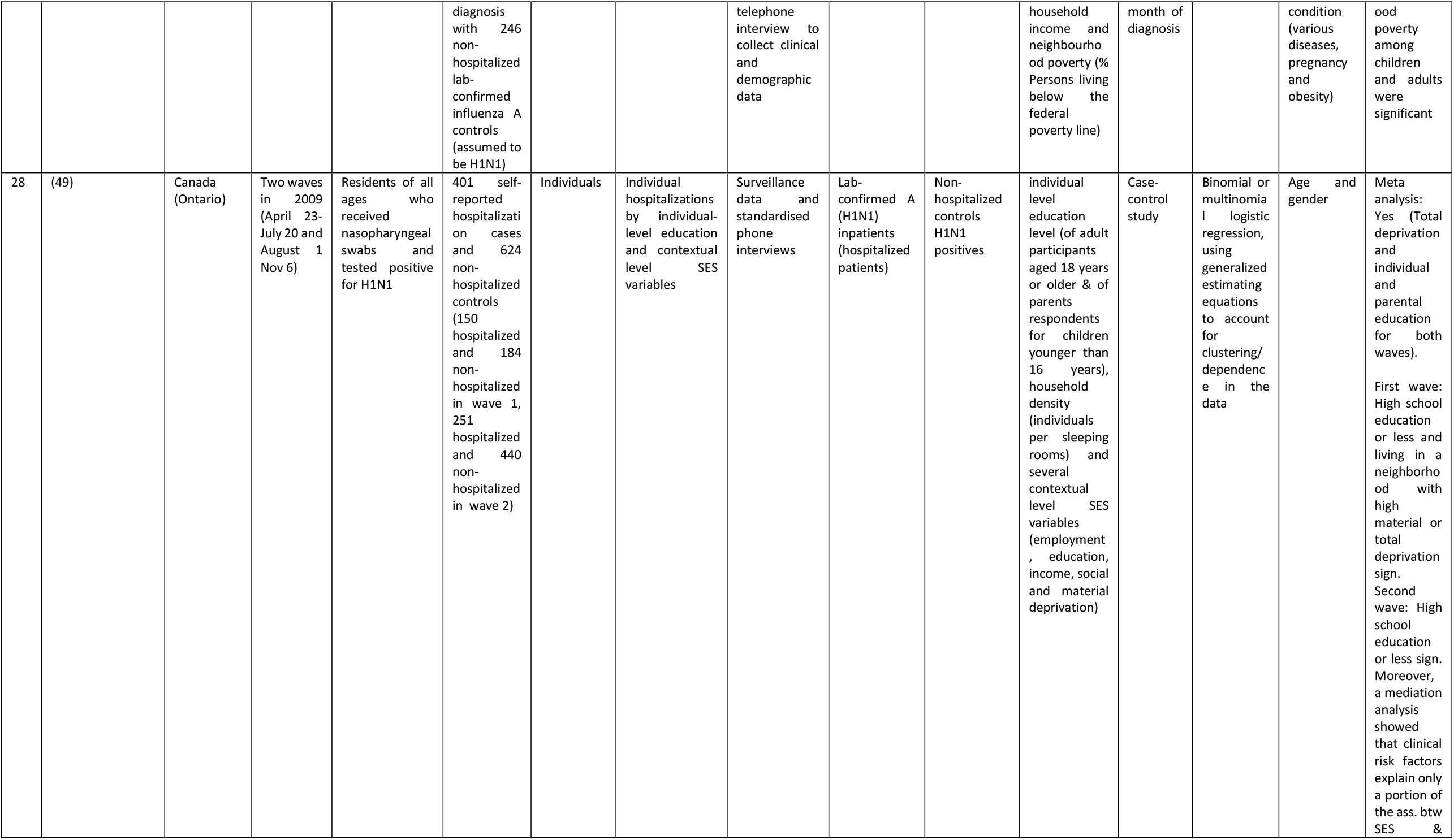
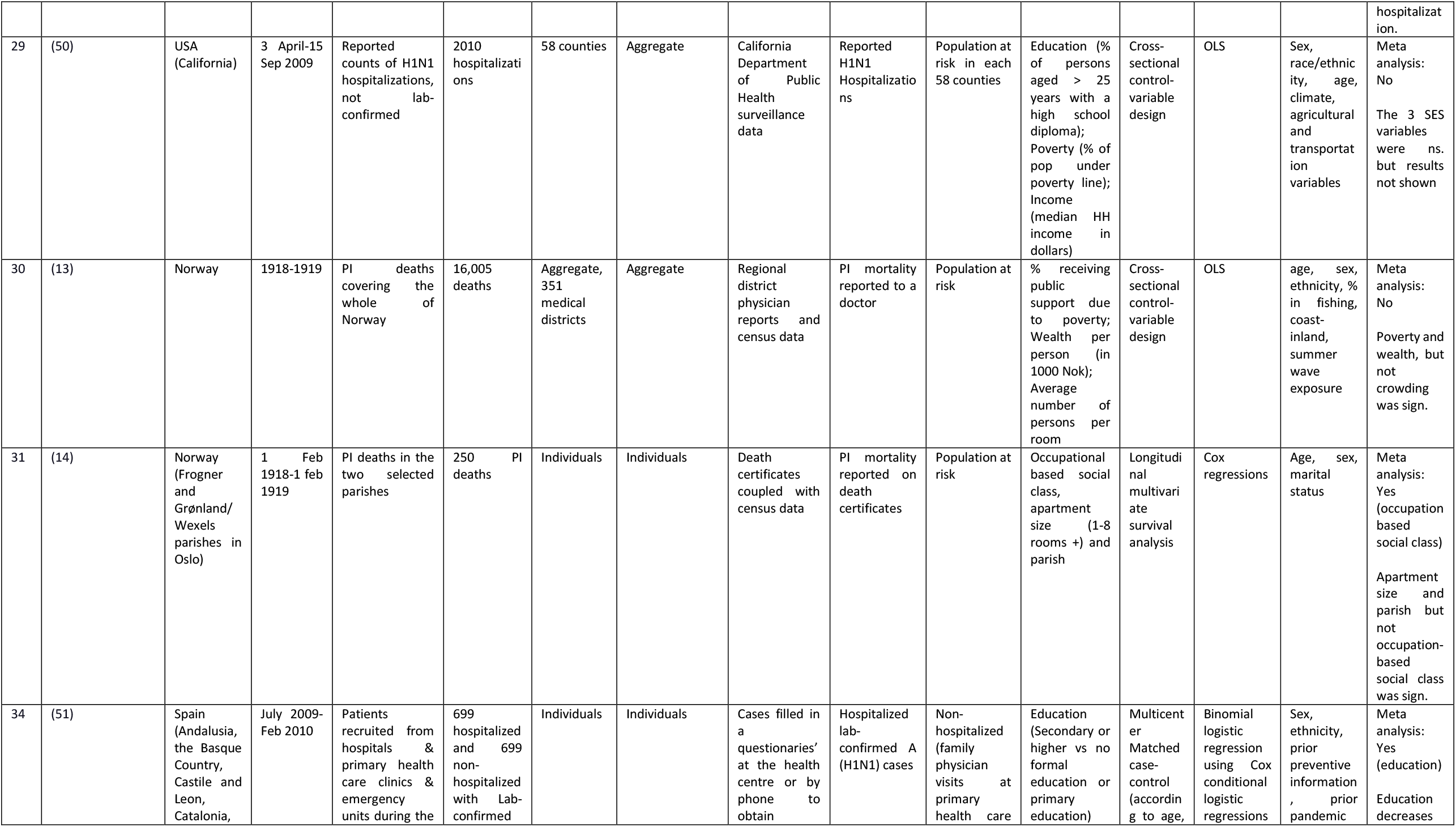
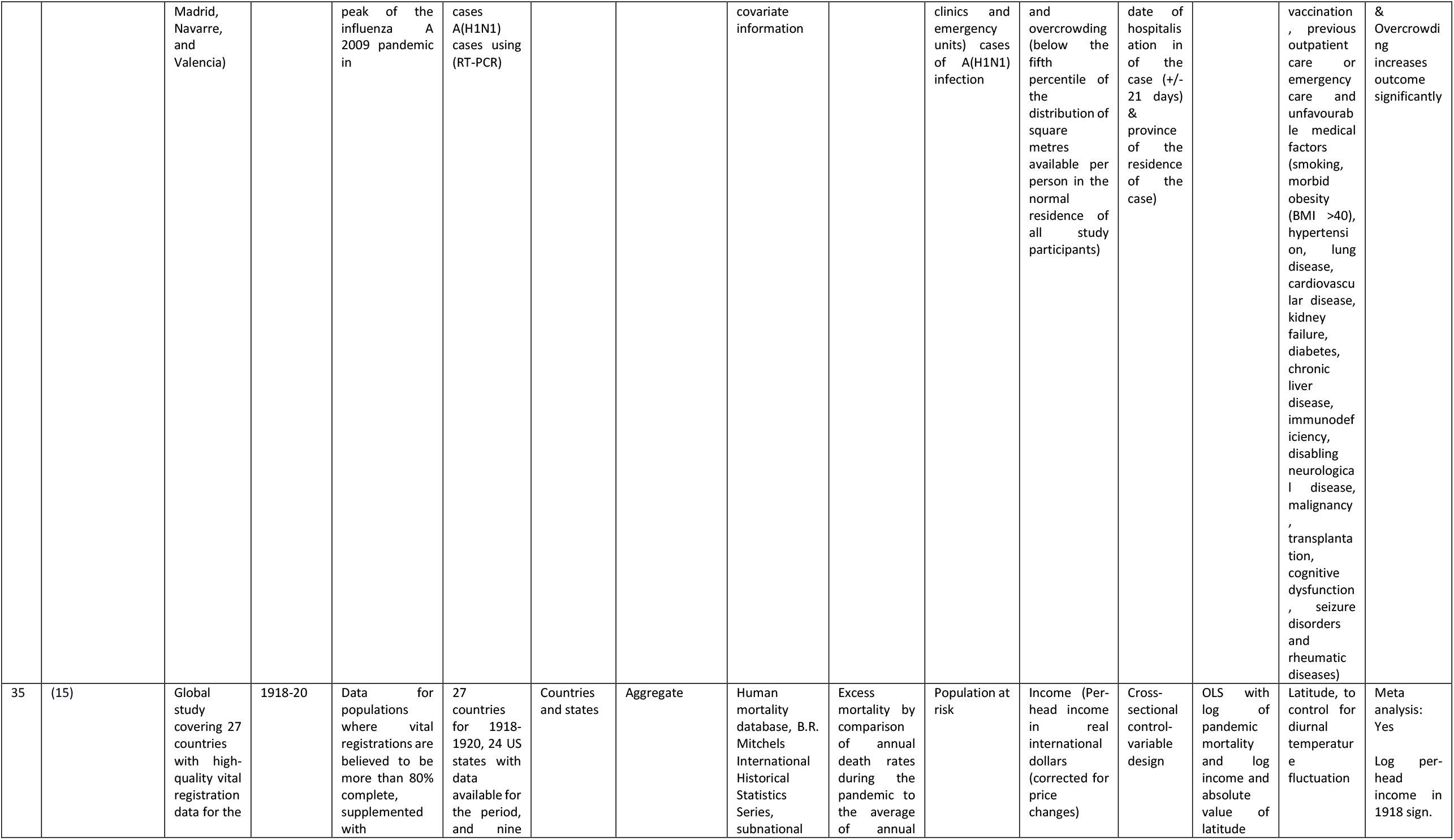
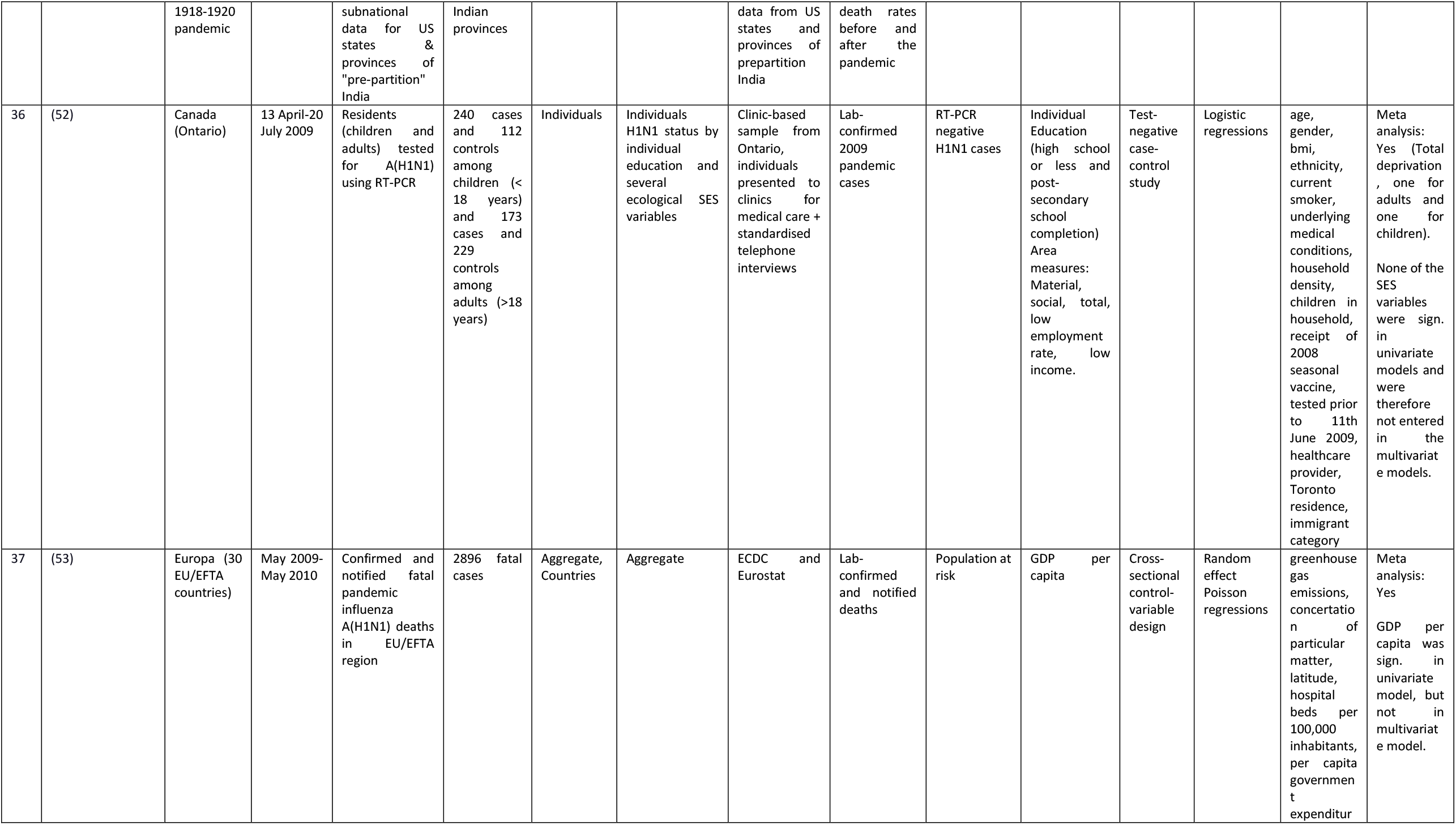
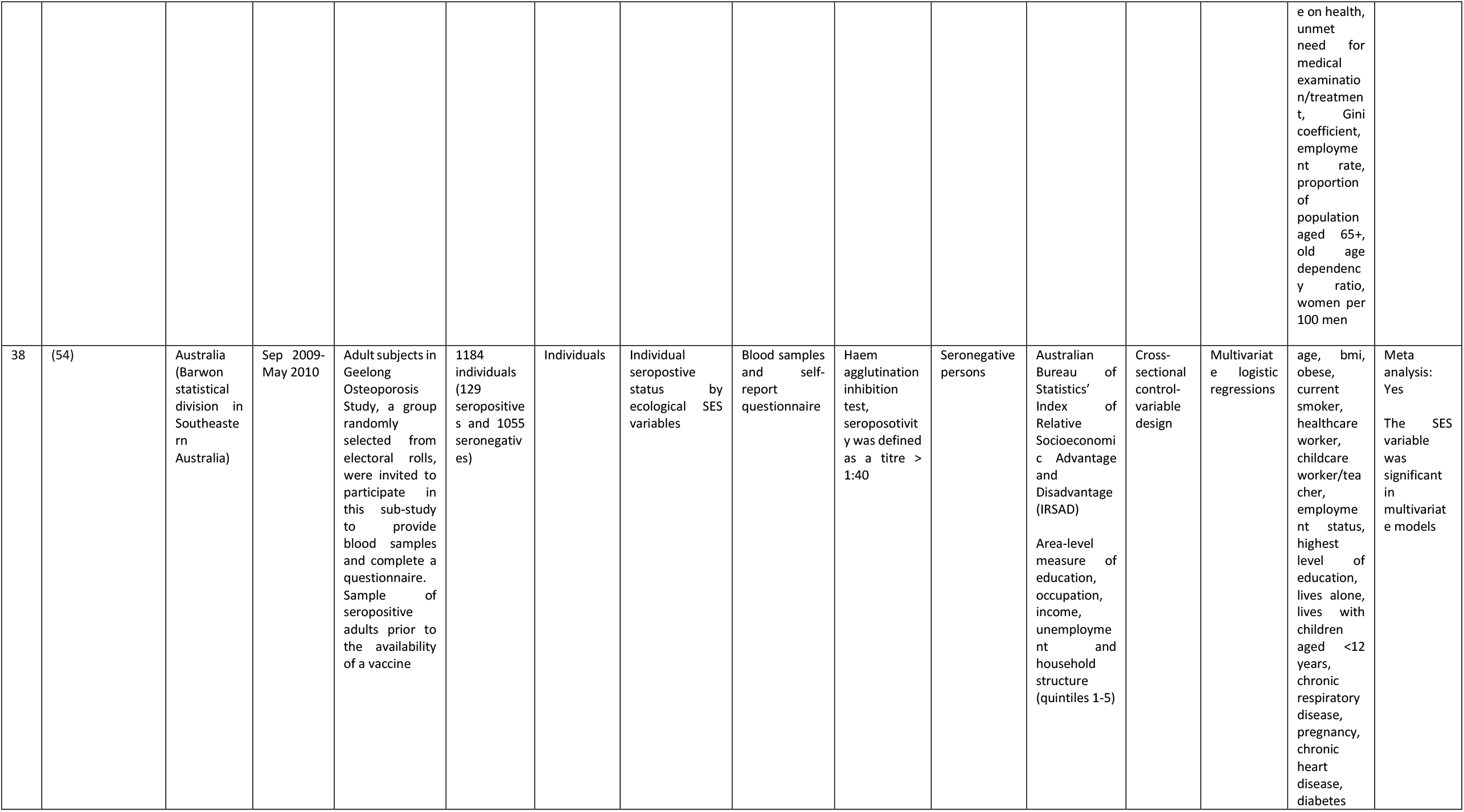
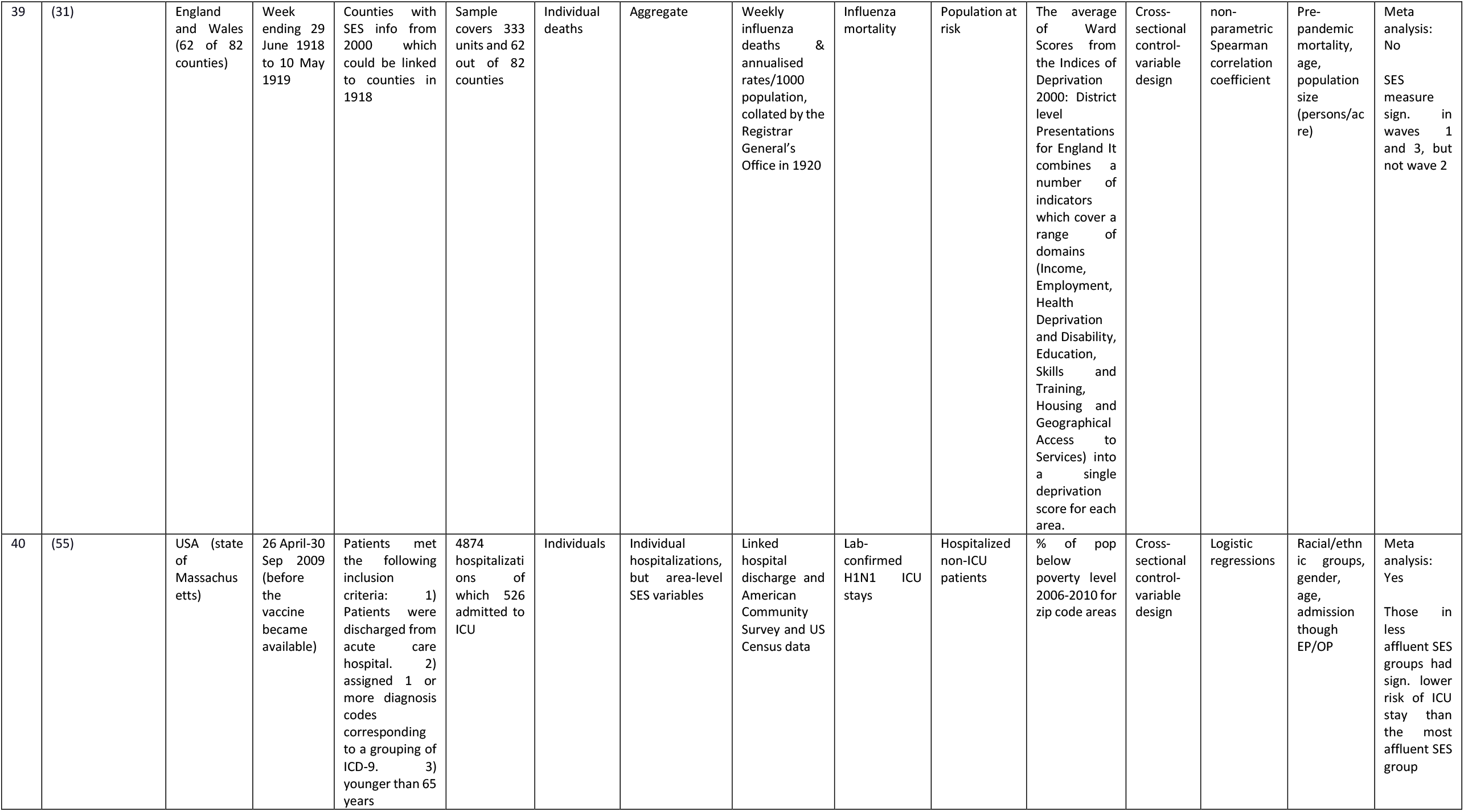
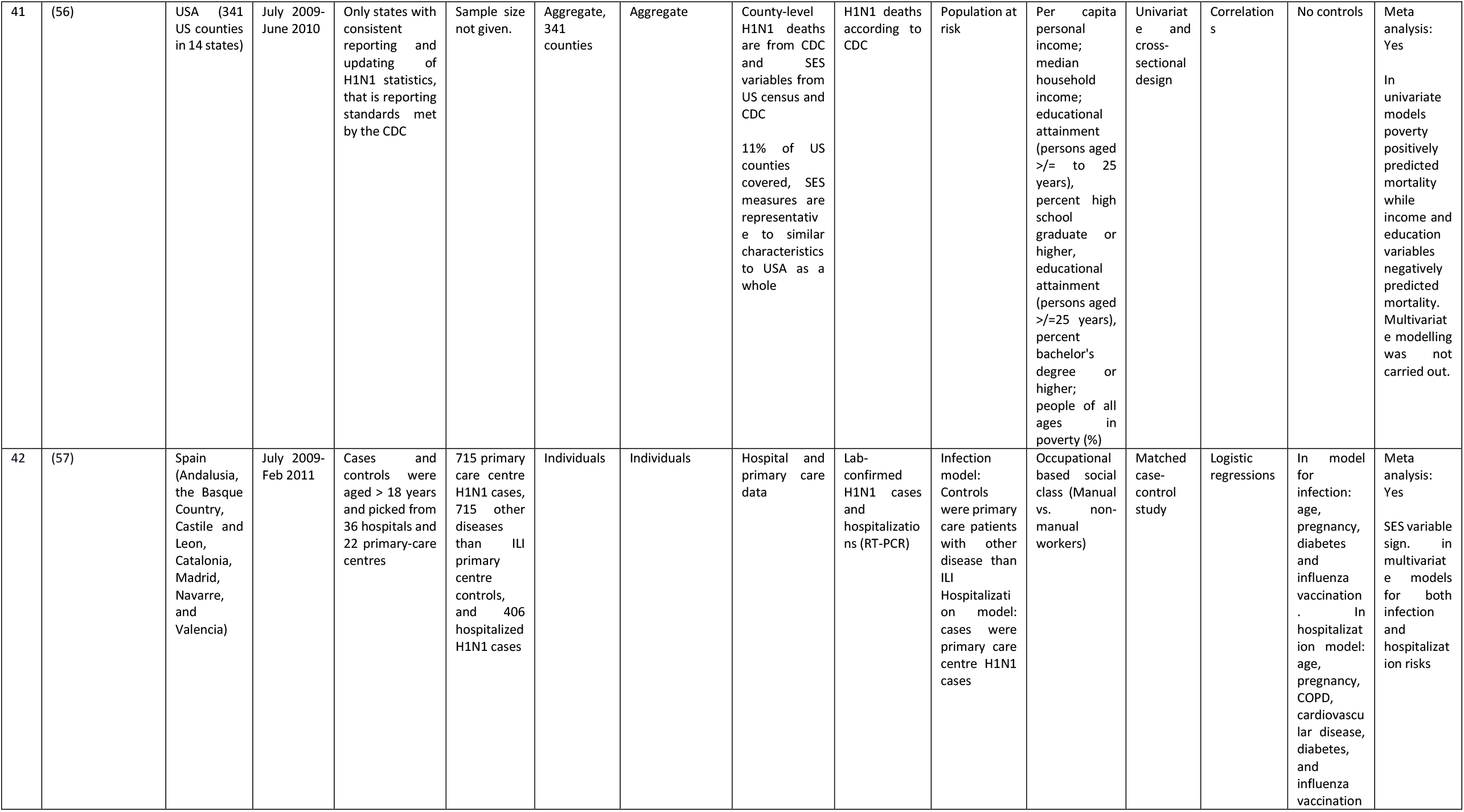
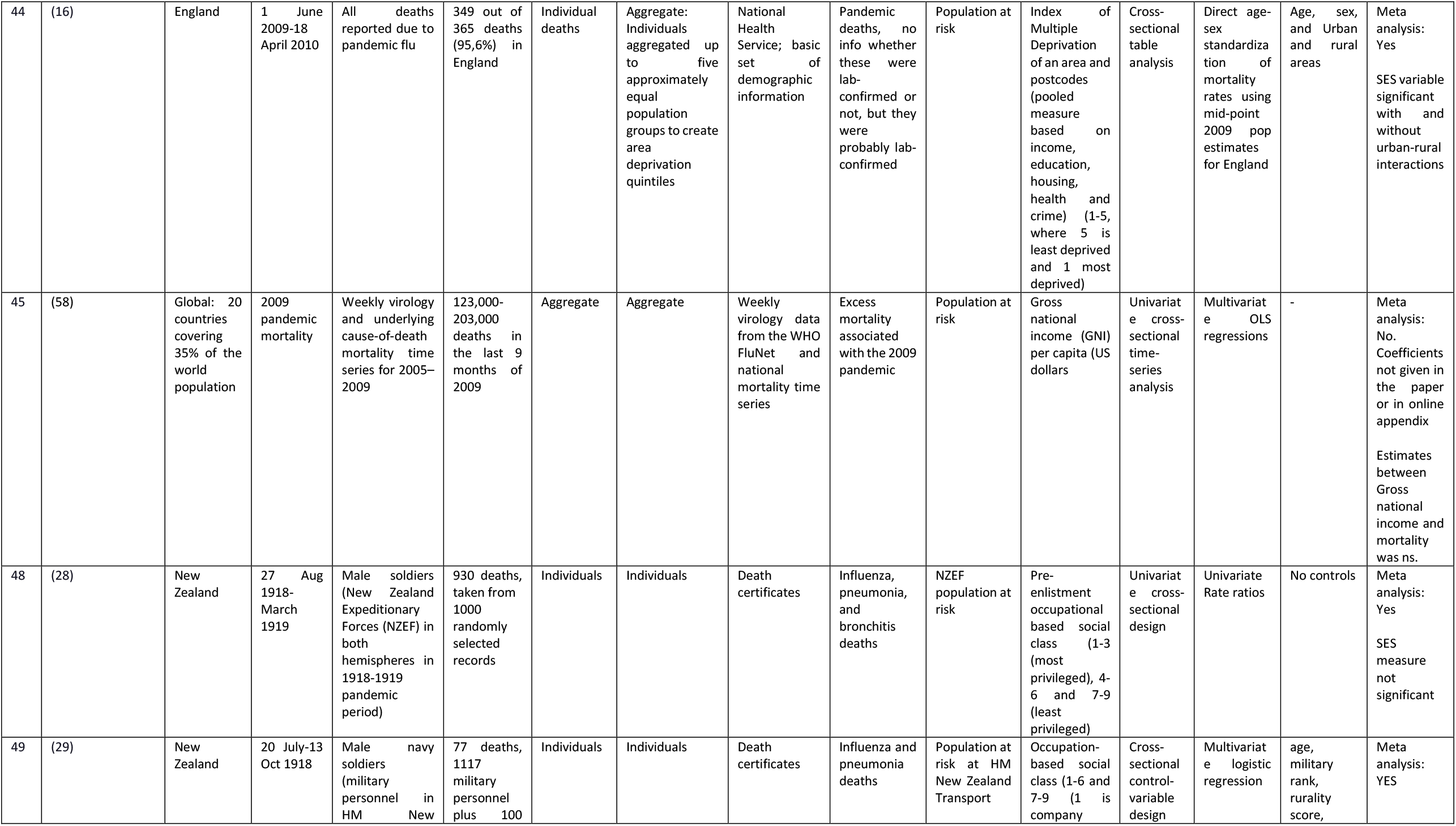
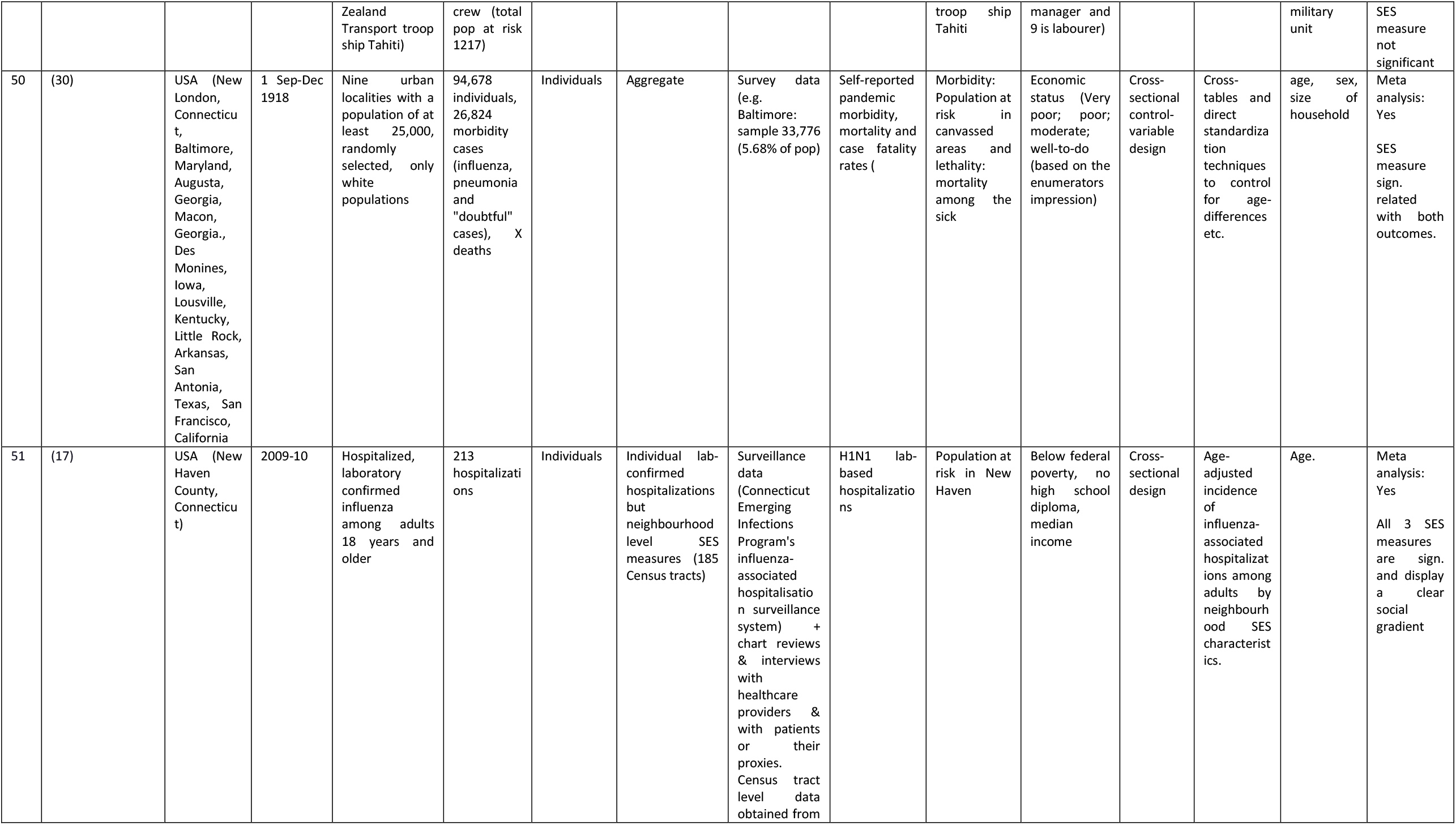
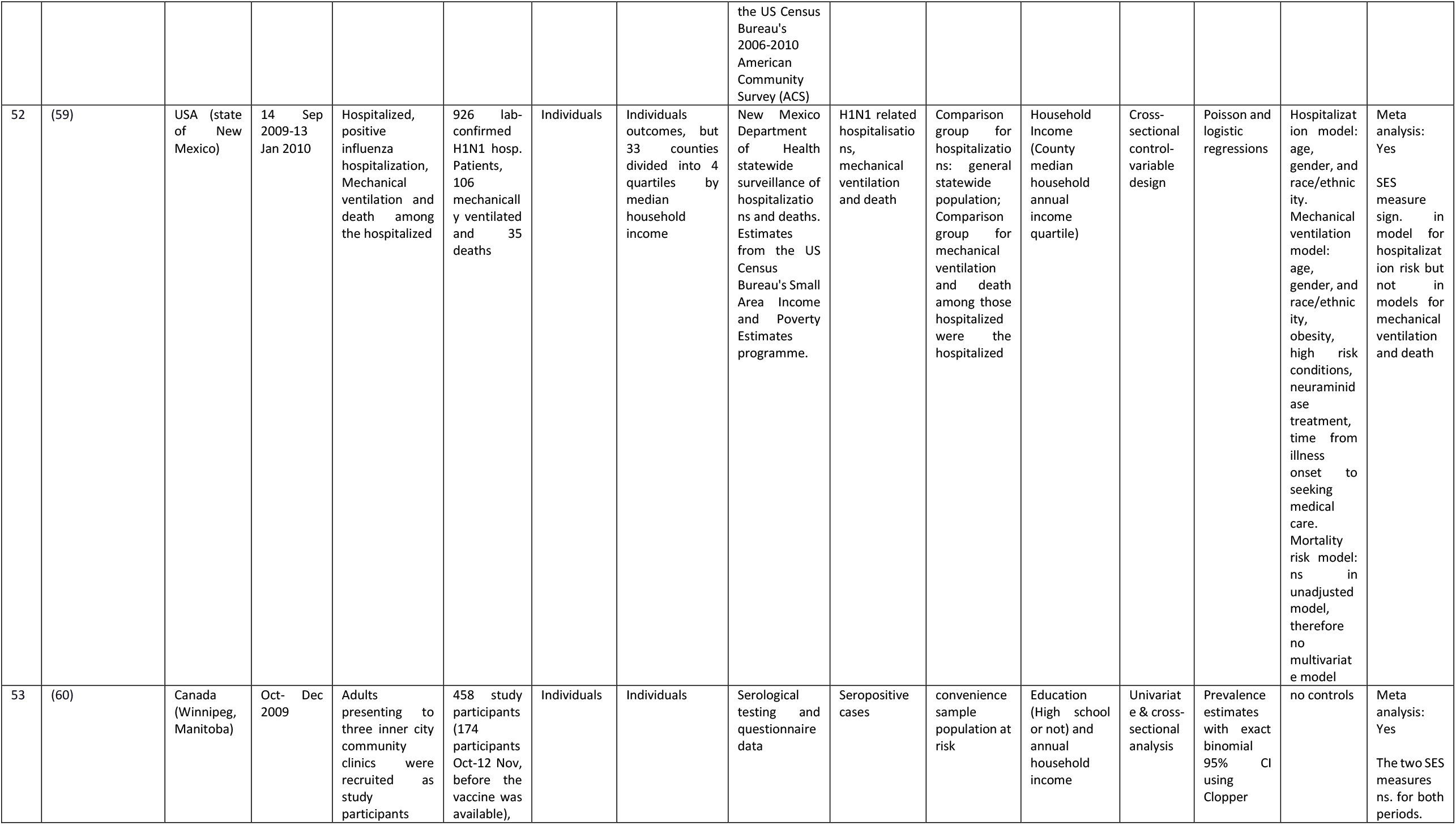
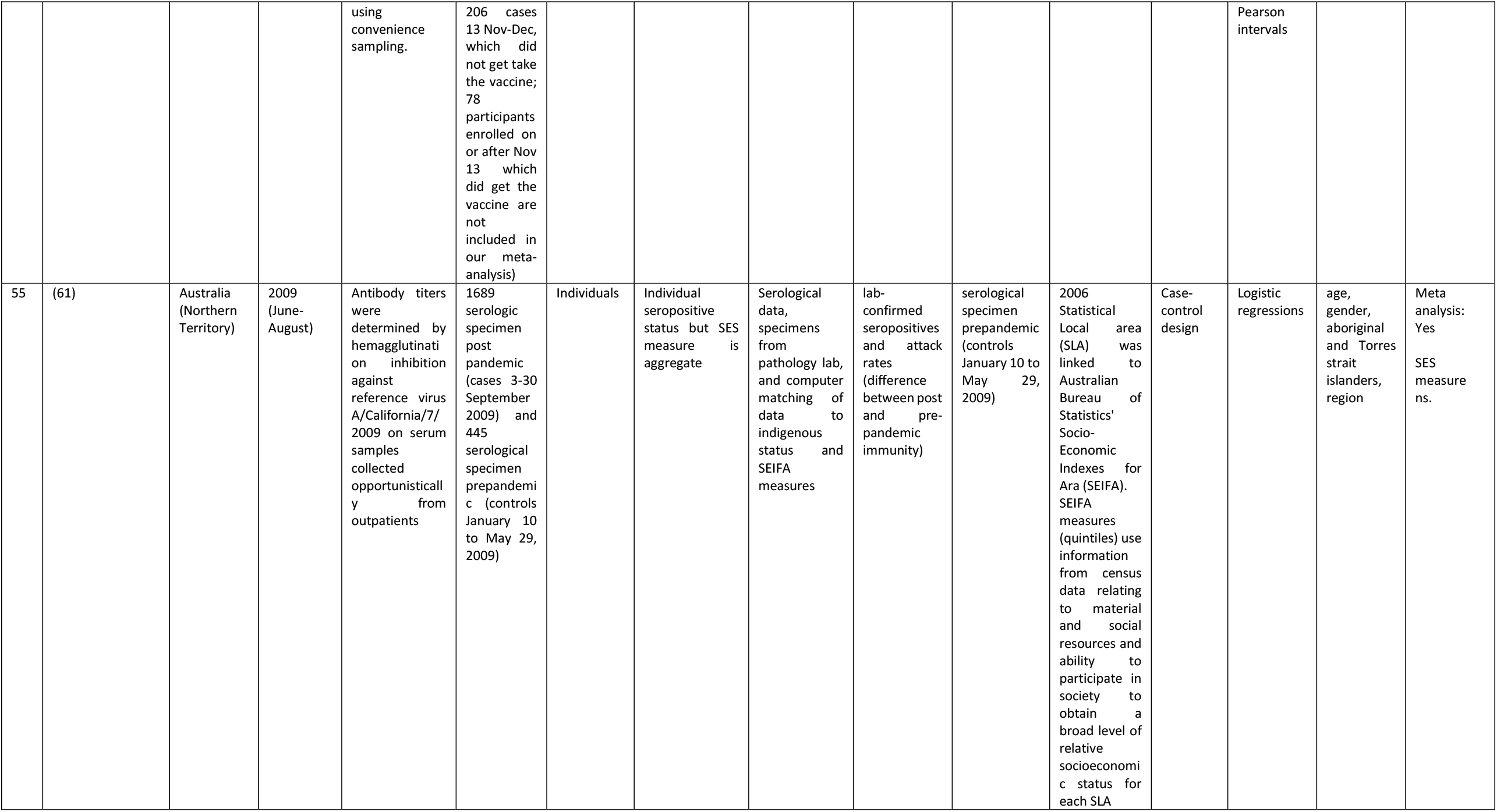
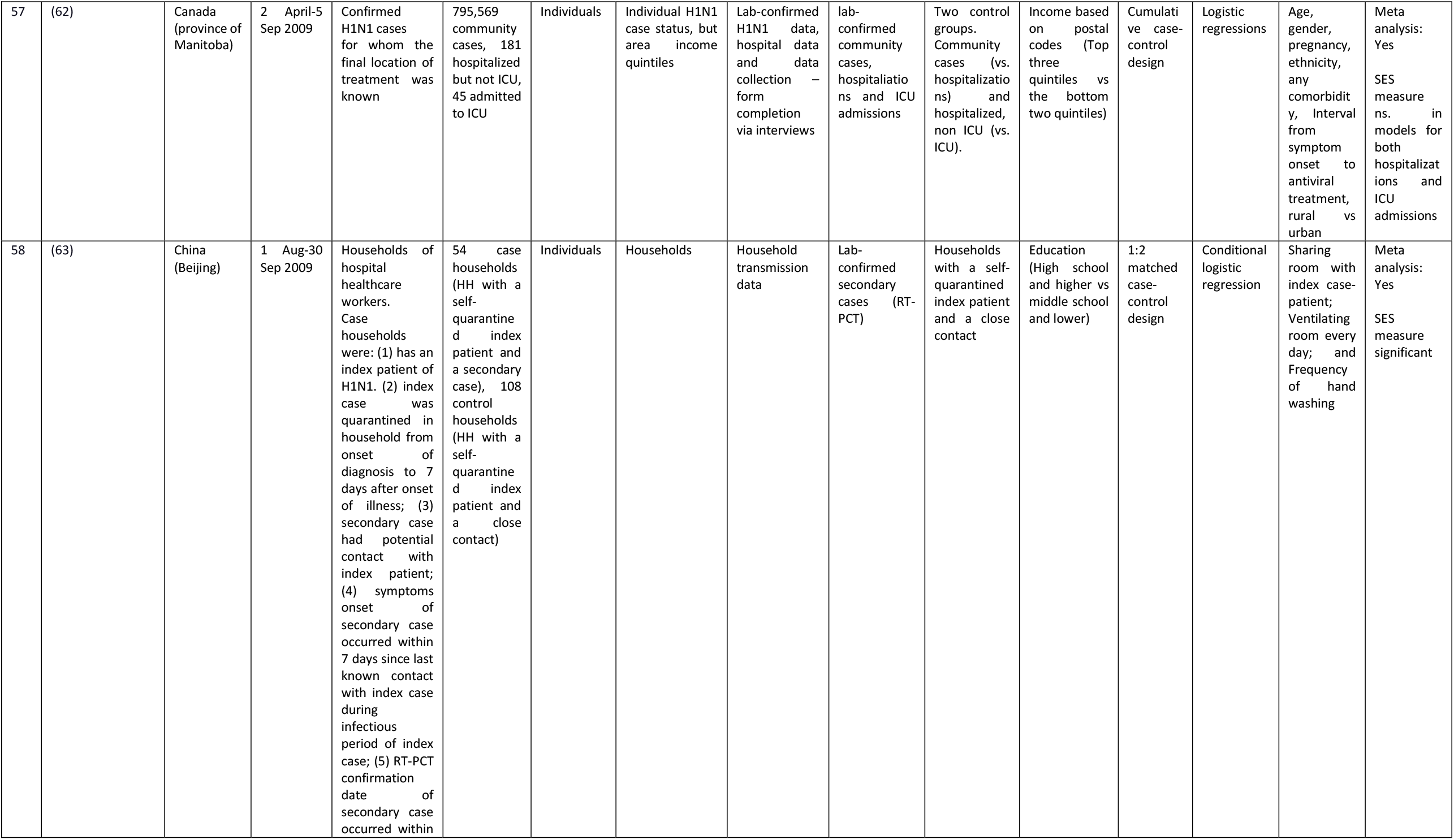
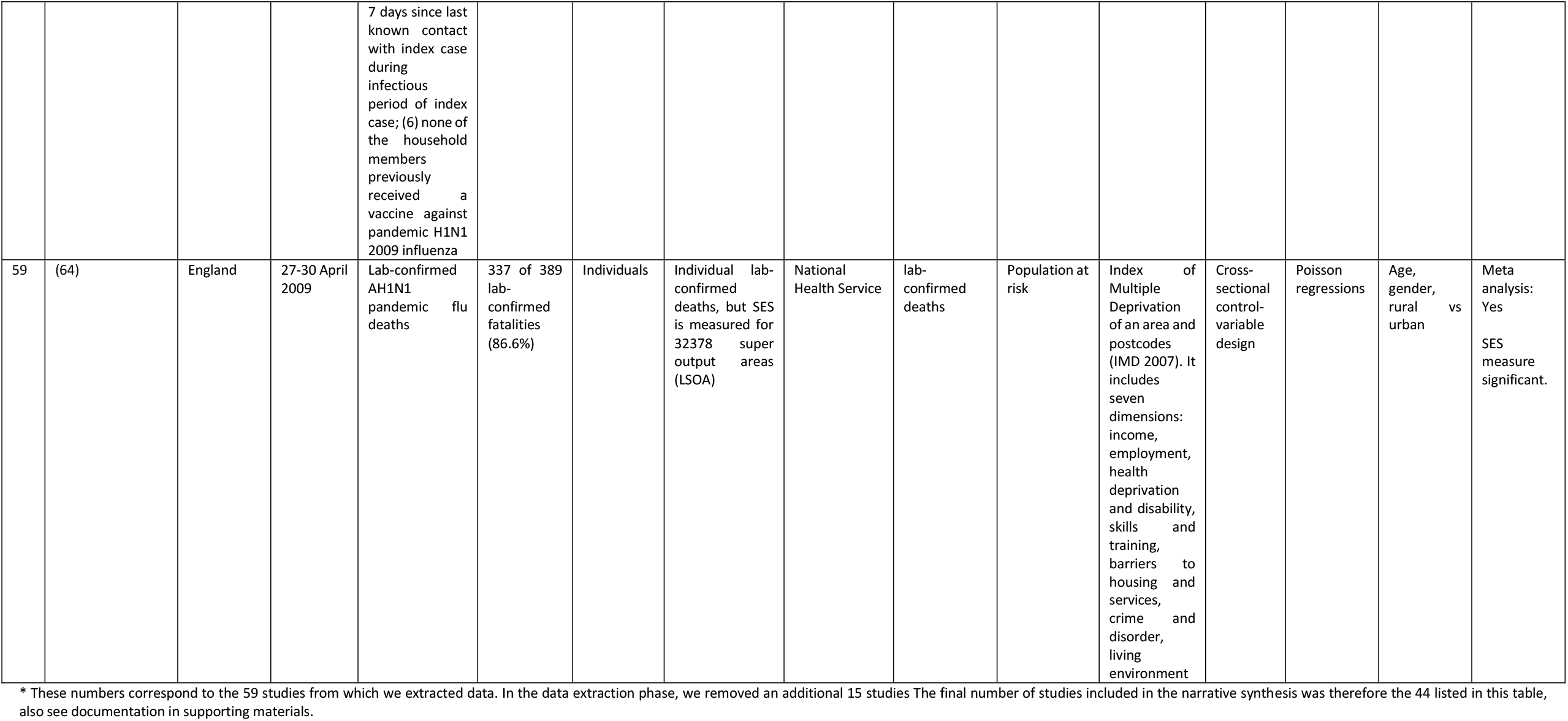
Overview of 44 studies included in the systematic review by study characteristics.

A Bayesian model differs from the ones described above in that it includes a prior distribution for the parameters. A prior distribution expresses reasonable beliefs regarding the parameter values before running the analysis. The analysis calculates how likely the observed data is for different parameter values in the prior distribution, and updates the prior distribution in light of the data, resulting in a posterior distribution that blends the pre-existing knowledge encoded in the prior with the evidence from the observed data. In our context, the benefit of such an approach is that it allows us to assess the joint impact of multiple study level indicators simultaneously despite having few observations, by viewing the parameters for each study level indicator as a draw from a distribution (i.e., a hierarchical specification). Such a hierarchical specification reduces the danger of reporting large but spurious associations that are statistically significant by chance, since the hierarchical specification imposes a partial pooling across the parameters (25). If the evidence as a whole indicates that estimates vary no more across study level indicators than we would expect due to sampling variation, then this will pull the individual indicator coefficients towards zero.

## Results

### Narrative review

#### Flow of included studies

Our database search identified 8,411 records. After leaving out duplicates, 4,203 studies were imported for screening. After removing another 75 duplicates, we screened the titles/abstracts of 4,128 records. Of these, 3952 studies were irrelevant, and 176 full text studies were then assessed for eligibility. In this phase, 117 studies^1^ were excluded leaving us with 59 studies from which to extract data. In the data extraction phase, we removed an additional 15 studies^2^. The final number of studies included in the narrative synthesis was therefore 44 (see also PRISMA Flow Chart in appendix).

### Study characteristics

The review identified a total of 44 studies, 9 studies of “Spanish flu of 1918-20” (13-15, 26-31) and 35 of the “Swine flu of 2009-2010” (16, 17, 32-64) (Table 1). We found no studies of the Russian flu of 1889-90, the Asian flu of 1957-58 or Hong-Kong flu of 1968-70. Most of the studies used data from North America, including 11 for USA (17, 27, 30, 33, 40, 41, 48, 50, 55, 56, 59) and 6 for Canada (38, 45, 49, 52, 60, 62); Europe, including 6 for England (16, 26, 31, 32, 44, 64), 4 for Spain (39, 46, 51, 57), 2 for Norway (13, 14), and 1 for 30 EU/EFTA countries (53); 4 for Australia (42, 43, 54, 61) and 3 for New Zealand (28, 29, 34). While a few studies used data from Central America/South America including 1 for Mexico (37) and 1 for Brazil (47), and Asia, including 1 for Iran (35) and 1 for China (63), we identified no studies using data from Africa. Finally, 3 studies had a global approach studying several countries (15, 36, 58).

The sample inclusion criteria varied greatly from study to study. Two of the 44 studies studied military populations, one of these studied mortality in randomly selected records (28), the other studied mortality on one transport troop ship (29). Of the 42 studies using civilian study populations, some studied particular patient populations/cohorts (46, 54, 61, 63), general patients at various hospitals and health centres (17, 32, 33, 35, 39, 40, 47-49, 51, 52, 55, 57, 59, 60, 62), students at schools or students including their families (41, 45), or general populations living in various cities, states, counties or (several) countries (13-16, 26, 30, 31, 34, 36-38, 42-44, 50, 53, 56, 58, 64).

The sample size in each study varies substantially and is reported in Table 1 whenever information was available for the pandemic events (for cases and controls) and the population at risk.

The unit of the outcome variables is either individual in 36 studies (14, 16, 17, 27-40, 42-49, 51, 52, 54, 55, 57, 59-64) or aggregate in 8 studies (13, 15, 26, 41, 50, 53, 56, 58). However, although a study may have had individual-level outcome data, the data aggregation level is sometimes aggregate. In total, 12 studies included studies at an aggregate data level (13, 15, 16, 26, 30, 31, 36, 41, 50, 53, 56, 58). 15 studies had individual-level outcome variables and control variables, but used area-level (and individual-level) SES variables (17, 27, 32, 33, 42-44, 49, 52, 54, 55, 59, 61, 62, 64). Studies using only ecological SES variables thus picked up a combination of individual-level and area-level SES effects on the outcome variables. Finally, in 17 of the studies, outcomes, explanatory variables and controls are all measured for individuals and the data aggregation level was thus the individual level (14, 28, 29, 34, 35, 37-40, 45-48, 51, 57, 60, 63).

There were generally three types of data source used in the 44 studies included in the narrative synthesis: 1) 28 studies used active surveillance of events coupled with SES and covariate data via questionnaires, face-to-face or telephone interviews or censuses (17, 32-44, 46-52, 54, 57, 59-63); 2) 14 studies used national vital registration systems on events coupled with SES and covariate data via censuses (13-16, 26-29, 31, 53, 55, 56, 58, 64); 3) 2 studies used telephone survey or data collected via door-to-door survey to collect both event and population at risk data (30, 45).

The 3 broad categories of outcomes were studied (see details in Table 1): 1) people seeing doctors due to symptoms of influenza like illness (ILI)/influenza transmission(R0)/lab-confirmed influenza infection (using PCR tests)/immunity towards influenza (using blood serum samples to look for antibodies) (26, 27, 30, 32, 34, 35, 41-45, 52, 54, 57, 60, 61, 63); 2) lab-confirmed influenza hospitalizations/ICU treatment/mechanical ventilation (17, 33, 38, 39, 46-51, 55, 57, 59, 62); 3) lab-confirmed pandemic deaths/Influenza-Pneumonia (PI) deaths/excess deaths associated with pandemic influenza (13-16, 26-31, 36, 37, 40, 53, 56, 58, 59, 64).

The choice of baseline outcomes (or controls in case-control studies) partly depends on the outcomes studied, and includes: 1) General population at risk (13-17, 26-33, 36, 50, 53, 56, 58-60, 64); 2) General population at risk without H1N1 Infection or ILI (41-45); 3) Patients with ILI, persons in quarantine for a suspected case and a close H1N1 contact or patients with ILI testing negative for influenza A H1N1 infection (30, 35, 52, 63); 4) pre-pandemic immunity (34, 61); 5) seasonal influenza A deaths (37); 6) Non-hospitalized H1N1 positive patients or hospitalized H1N1 positive non-severe (not ICU or death) (38, 39, 55, 59, 62); 7) Outpatients with H1N1 infection (40, 46-49, 51, 57); 8) Seronegative for H1N1 (54); 9) Patients with other diseases than ILI (57).

The studies that used individual-level SES measures used one or several of the following; (household) income (40, 48, 60), economic status (30), education (35, 37-40, 46-49, 51, 52, 60, 63), occupation-based social class (14, 28, 29, 57), size of apartments, poor housing or crowding measures (14, 26, 34, 40, 45, 49, 51), and having health insurance (40). Some used both individual-level and area-level measures of SES. The SES measures used at the area-level are often (but not always) indexes of economic, social and housing deprivation/development (13, 15-17, 27, 31-33, 36, 41-44, 48-50, 52-56, 58, 59, 61, 62, 64).

The 44 studies included in the review used study designs that falls into four categories: 1) Systematic review and meta-analysis (36); 2) Cross sectional univariate or control-variable design (13, 15-17, 26-34, 42-45, 50, 53-56, 59, 60, 64); 2) Case-control design (35, 37-41, 46-49, 51, 52, 57, 61-63); 3) Longitudinal survival analysis (14); 4) Time-series analysis (58).

The identified studies were descriptive or explanatory. The descriptive studies used statistical techniques to calculate pandemic disease burden estimates and univariate correlations between the outcomes and various variables as well as demographic standardization techniques to control for age and sex (16, 17, 28, 30-33, 44, 56, 60). The explanatory multivariate studies used modelling techniques such as OLS (13, 15, 50, 58), generalized linear mixed models (45), logistic regressions (26, 29, 34, 35, 38-41, 46-49, 52, 54, 55, 57, 59, 61-63), propensity score logistic regressions (37), Poisson regressions (27, 53, 59, 64), Cox regressions (14, 51), random effect meta-regressions (36), and various types of Bayesian models (42, 43).

### Study results

The results in the 9 identified studies on the 1918 influenza and SES were mixed (13-15, 26-31). After various controls were made, 6 studies found a significant and expected *higher* risk for *lower* SES in mortality (15) or mortality/transmission rates, but not for all SES measures (13, 14, 27); a significant *higher* mortality risks for *lower* SES, but only for 2 out of 3 pandemic waves (31); or a significant *higher* risks for *lower* SES in both for morbidity and mortality (30), while 3 studies found no association between SES and mortality (28, 29) or mortality and transmission rates (26). However, none of the 6 studies documenting significant expected associations with a higher pandemic risk for lower SES had data to control for medical risk factors. Hence, some or all of the identified associations between SES and the pandemic outcomes in the 6 above mentioned studies could potentially have been “explained away” by controlling for having latent tuberculosis (65) or other known comorbidities (66).

To get an idea as to whether SES *may* have played an independent role in the variation in pandemic outcomes in 1918, we now describe the results for the identified studies of the 2009 pandemic. Fourteen of the 35 identified studies on the 2009 pandemic had data to adjust for both medical and social risk factors (34, 35, 38, 40, 45-48, 51, 52, 54, 57, 59, 61); after adjusting for medical risk factors, 7 of these studies documented independent and *expected* impact of SES (*higher* risks for *lower* SES) on either infection/immunity (34, 54), hospitalization (46-48, 51) or both of these outcomes (57); 1 study found both expected significant associations with SES (higher risk of hospitalization) and non-significant (ICU and death) impact of SES after medical risk factor were controlled for (59); 5 studies found non-significant effects of SES on ILI/infection/immunity (45, 52, 61), hospitalization/ICU (38) and mortality (40); and finally, 1 study found a significant but *unexpected* impact of SES on infection, that is *higher* infection rates for those with *high*er vs. lower education (35). Although the findings in these 14 studies investigating both social and medical vulnerabilities are somewhat mixed, they show that medical risk factors are not simply 100% correlated with socioeconomic factors, and in 8 of these 14 studies social factors explain other parts of the variation in the pandemic outcomes than medical factors.

21 of the 35 identified studies on the role of SES in the 2009 pandemic outcomes *did not* control for medical risk factors but found the following: First, 12 studies found significantly *higher* risks for the *lowest* socioeconomic status group, of which 5 studied ILI/infection/immunity (32, 43, 44, 63); 4 investigated hospitalizations (17, 33, 39, 49); and 4 studied mortality (16, 36, 56, 64). Second, 7 studies found non-significant associations with SES, of which 2 studied ILI/infection/immunity (42, 60); 2 studied hospitalizations (50, 62) or ICU treatment (62), and 3 studied mortality (37, 53, 58). Finally, 2 studies found respectively a higher risk of a lab-confirmed case (41) or ICU treatment (55) in the *highest* SES groups. It is clear though, that most of the studies on SES and 2009 pandemic not controlling for medical at risk factors (13 of 21), show that lower SES groups have the highest risks of the 3 considered pandemic outcomes.

### Quantitative meta-analysis

The quantitative analysis includes 46 estimates drawn from 35 of the 44 studies included in the narrative synthesis (14-17, 27-30, 32-35, 37-41, 45-49, 51-57, 59-64), and a standard random effects analysis of all estimates pooled finds a pooled effect mean odds ratio of 1.4 (95% CI: 1.2 – 1.7), comparing the low to the high SES groups. The pooled estimate is statistically significant at the 0.1 percent level, which means that we would have been highly unlikely to see an estimate of this or larger absolute magnitude if the true mean of the effect distribution was zero. As seen in the forest plot, the individual study estimates differ in both precision and location, with more variation in less precise estimates as we would expect (Figure 1).

**Figure 1.**
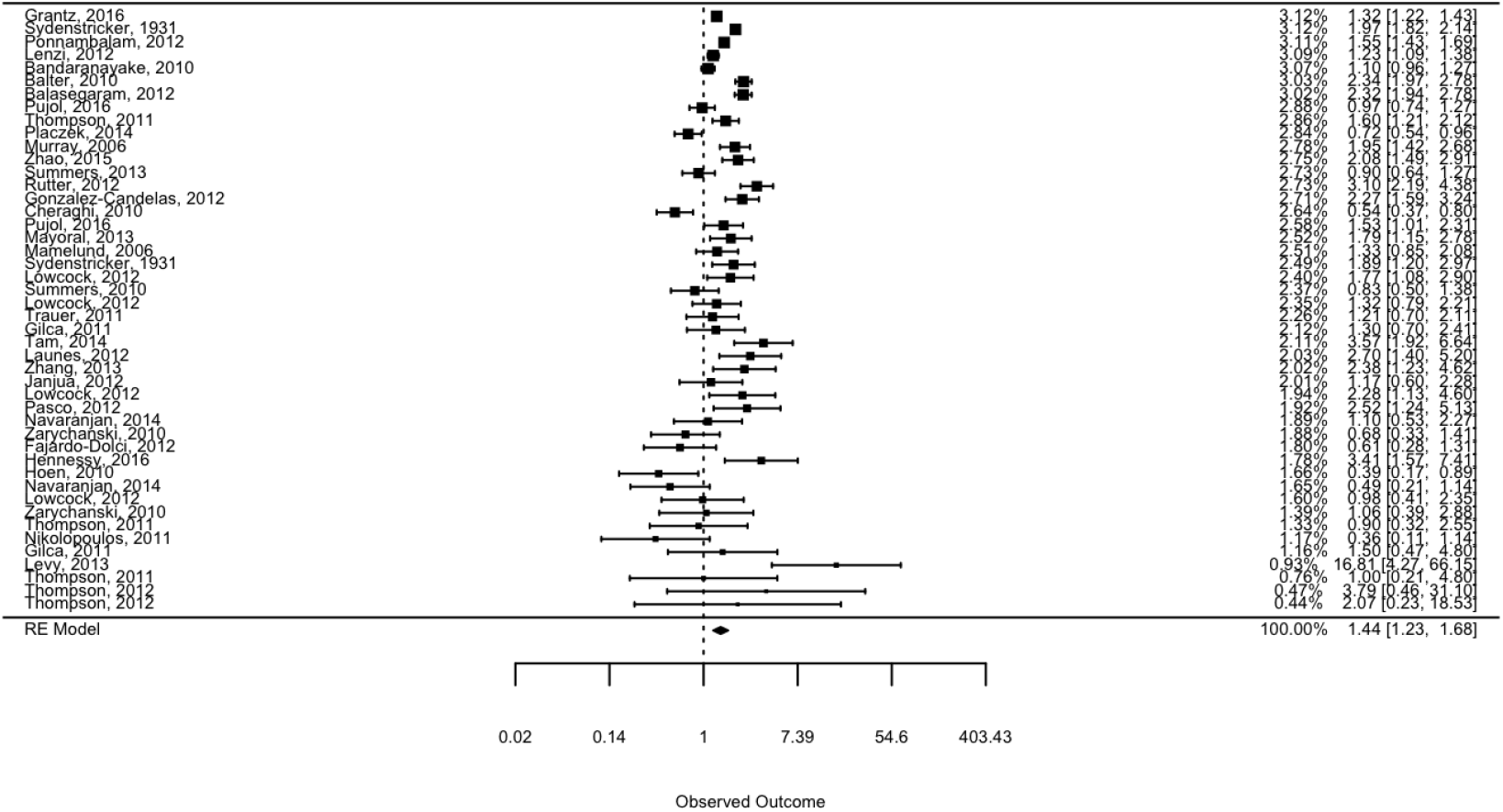
Forest plot. The plot shows the included estimates sorted by precision, along with their weights in the pooled effect estimate.

The random effect analysis finds strong evidence of effect heterogeneity across studies, with an estimated 92% of the total variation across studies reflecting effect differences rather than sampling variation. The estimated standard deviation of the effect distribution is labelled tau and has a point estimate of 0.45 on the log scale. If the underlying effects at the study level are normally distributed around their expectation, this tau is the standard deviation of study effects. Roughly fifty percent of studies would then be estimating “true” ORs in the range of 1.1-1.9. The Cochran’s Q test strongly rejects a test of zero heterogeneity (p < 0.0001), confirming the choice of a random effects over a fixed effect model. Subsample analyses indicated similar results in studies using individual level and aggregate SES indicators, case control and relative risk outcome measures, and studying the 1918 and 2009 pandemic period (Figure 2 and Table 2).

**Table 2.** Subsample analyses. The plot shows point estimates and 95% confidence intervals for different subsamples of studies, with a grey circle indicating the number of studies in each subsample.

| Distinction | Type | Number of estimates | Pooled RE effect | 95% CI lower bound | 95% CI upper bound | Tau |
| --- | --- | --- | --- | --- | --- | --- |
| Measure | Ecological | 20 | 1.38 | 1.05 | 1.80 | 0.53 |
|  | Individual | 26 | 1.45 | 1.20 | 1.76 | 0.41 |
| Period | 1918 | 7 | 1.42 | 1.10 | 1.83 | 0.30 |
|  | 2009 | 39 | 1.44 | 1.20 | 1.756 | 0.50 |
| Method | Relative Risk | 10 | 1.61 | 1.26 | 2.06 | 0.35 |
|  | Odds Ratio | 36 | 1.39 | 1.14 | 1.69 | 0.49 |

**Figure 2.**
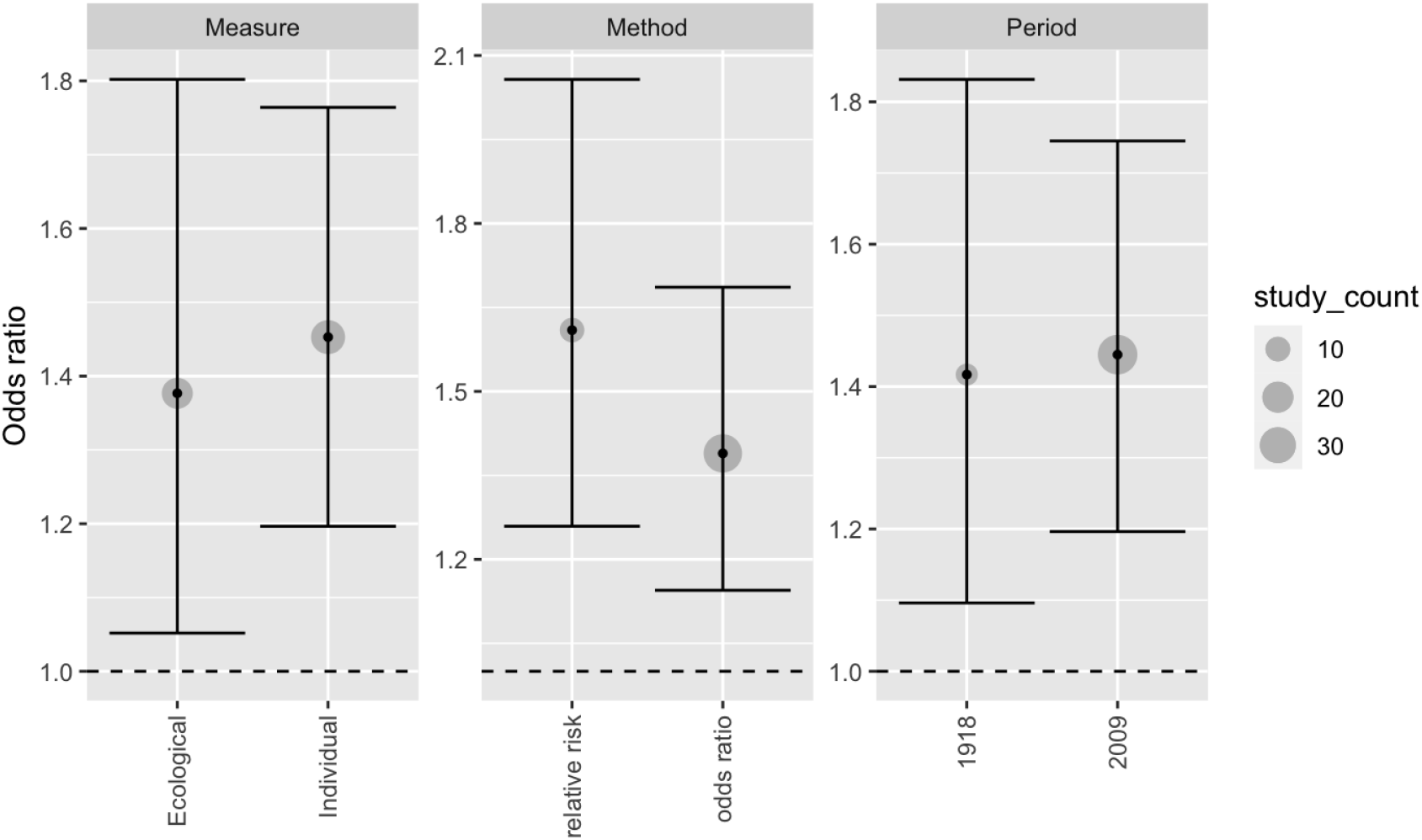
Subsample analyses. The plot shows point estimates and 95% confidence intervals for different subsamples of studies, with a grey circle indicating the number of studies in each subsample.

Subsamples were also defined by specific *combinations* of case and control outcomes (Figure 3). These suggest that studies examining the risk of flu outcomes relative to a general population (here defined as a control sample not selected on indicators of illness) tend to indicate a clear and substantial increased risk for lower SES groups. Studies comparing hospitalized to those infected also point to SES associations. Studies assessing the risk of severe cases (e.g., treatment in ICU or death) *conditional* on hospitalization are fewer, but seem to report no clear SES associations in any direction. Finally, studies using “other” control samples (e.g., patients with flu symptoms who did not have flu, people with non-pandemic flu during a pandemic period, patients accessing or being treated by health care systems for other reasons) tend to find no (or reversed) associations with SES indicators.

**Figure 3.**
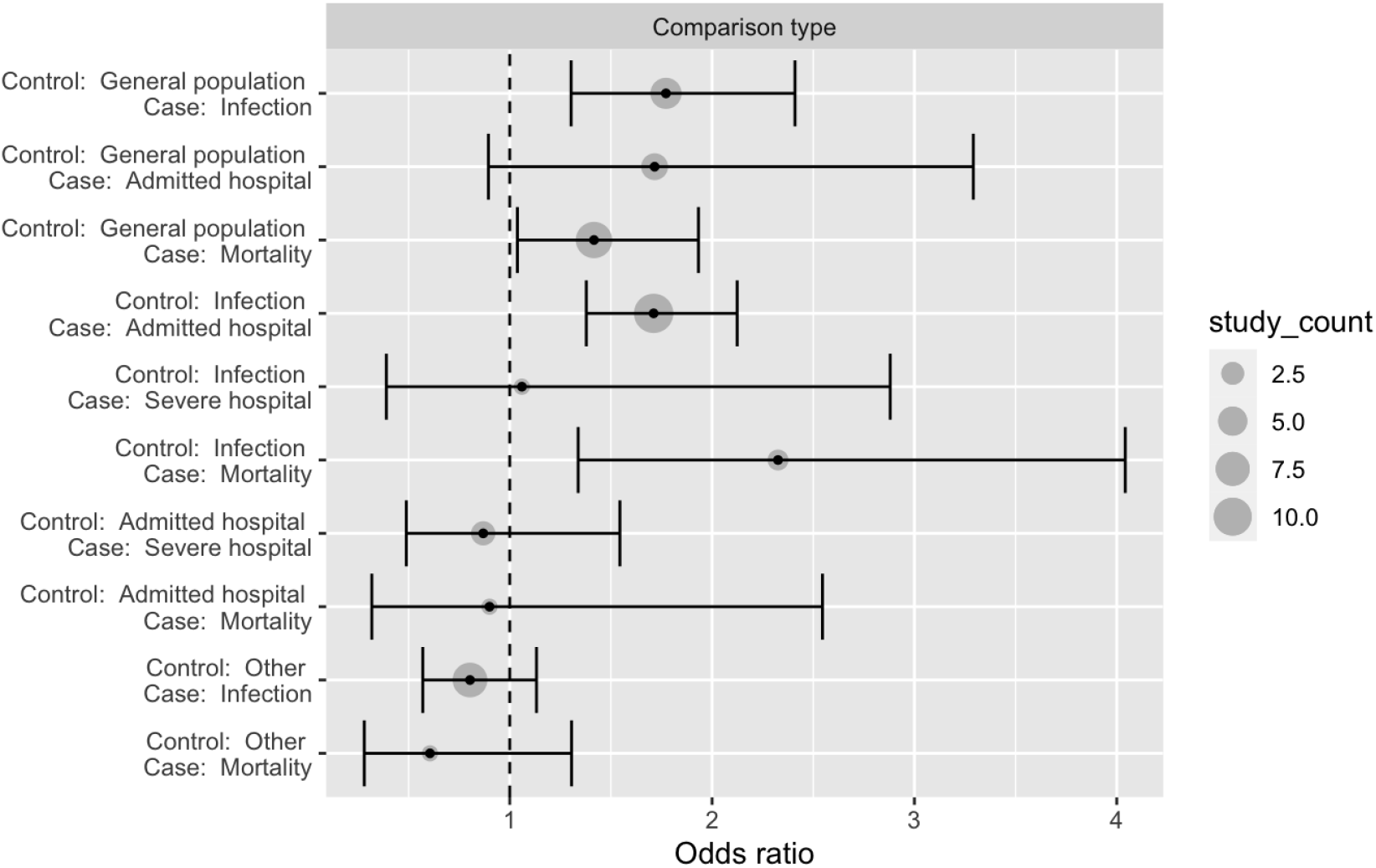
Subsample analyses. The plot shows point estimates and 95% confidence intervals for different subsamples of studies, with a grey circle indicating the number of studies in each subsample.

As all of these comparisons are based on different splits of the same study sample, they can be viewed as a series of univariate analyses. To assess the joint contribution of these study level features, and to include country/region indicators, we estimated two Bayesian models: One, without study level covariates, is closely analogous to the above meta-analysis, and was included to ensure that results from the two approaches are similar and comparable. This Bayesian model finds a pooled effect mean of 1.4 with a 95% credibility interval from 1.2-1.7, which is essentially identical to the above estimate of 1.4 (95% CI: 1.2 – 1.7). The estimated standard deviation of the underlying study parameters, analogous to the parameter tau in the earlier analysis, is estimated at 0.46 (0.3-0.6), the same as the above estimated tau of 0.45^3^. The second Bayesian model included all study level indicators (level of SES indicator, RR/OR indicator, period, case and control outcomes, and country/region), as well as an indicator for each unique *combination* of case and control outcome (as in Figure 3). Jointly, this reduces the estimated unexplained heterogeneity (tau) substantially, with the average value estimated dropping from 0.46 to 0.34)^4^.

As shown in Figure 4 and Figure 5, the Bayesian analysis finds similar results as the earlier subsample analyses, indicating that the patterns for the control and treatment outcome combinations are not “explained away” in an analysis when simultaneously accounting for other study level characteristics.

**Figure 4.**
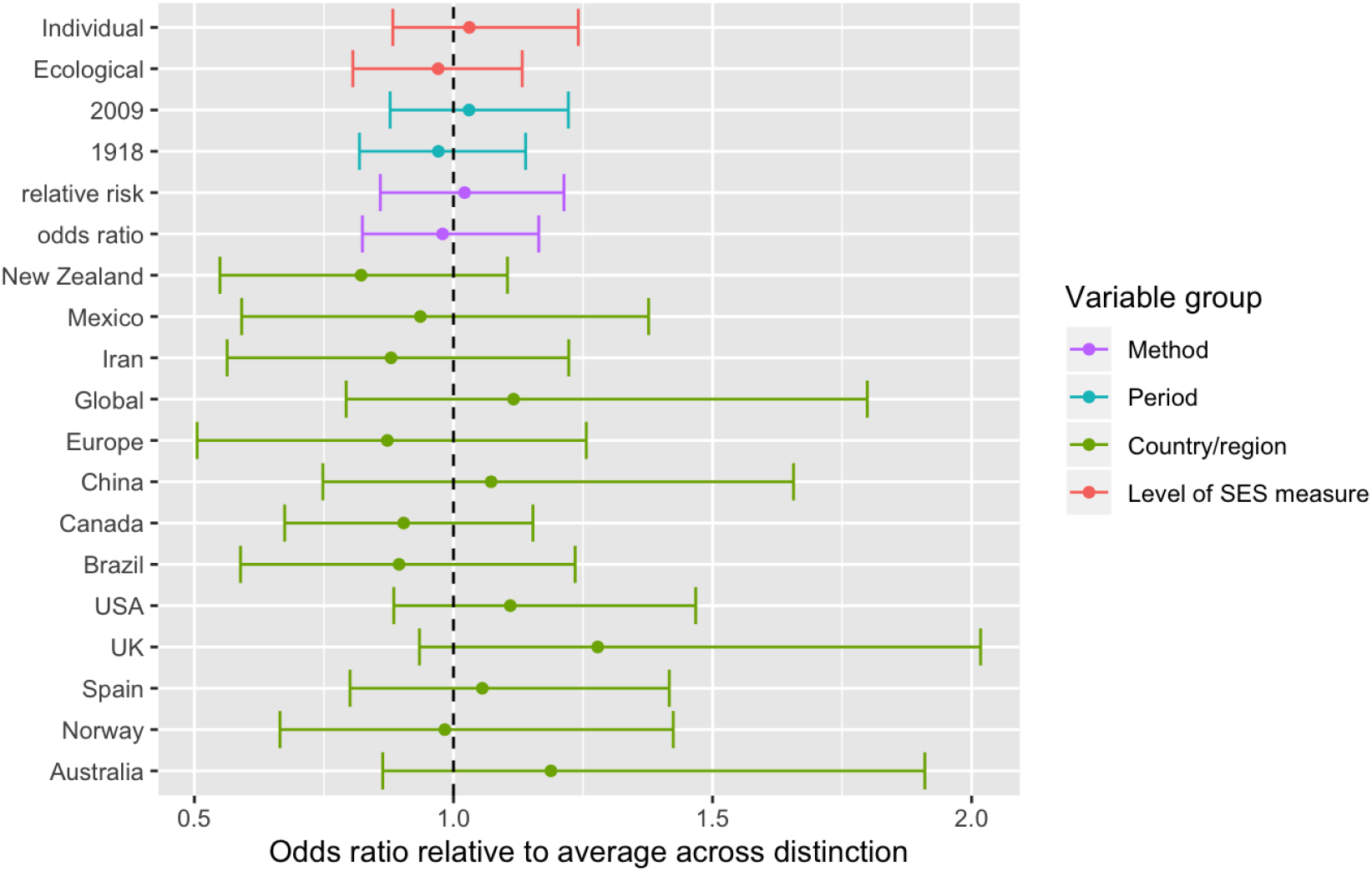
Differences across study level covariates. The plot shows average estimates and 95% credibility intervals for different study level covariates. The parameters are constrained to sum to zero within each category (e.g., for each draw from the posterior distribution, the sum of country parameters will sum to zero, as will the sum of the period parameters, etc.) See supporting materials for model details.

**Figure 5.**
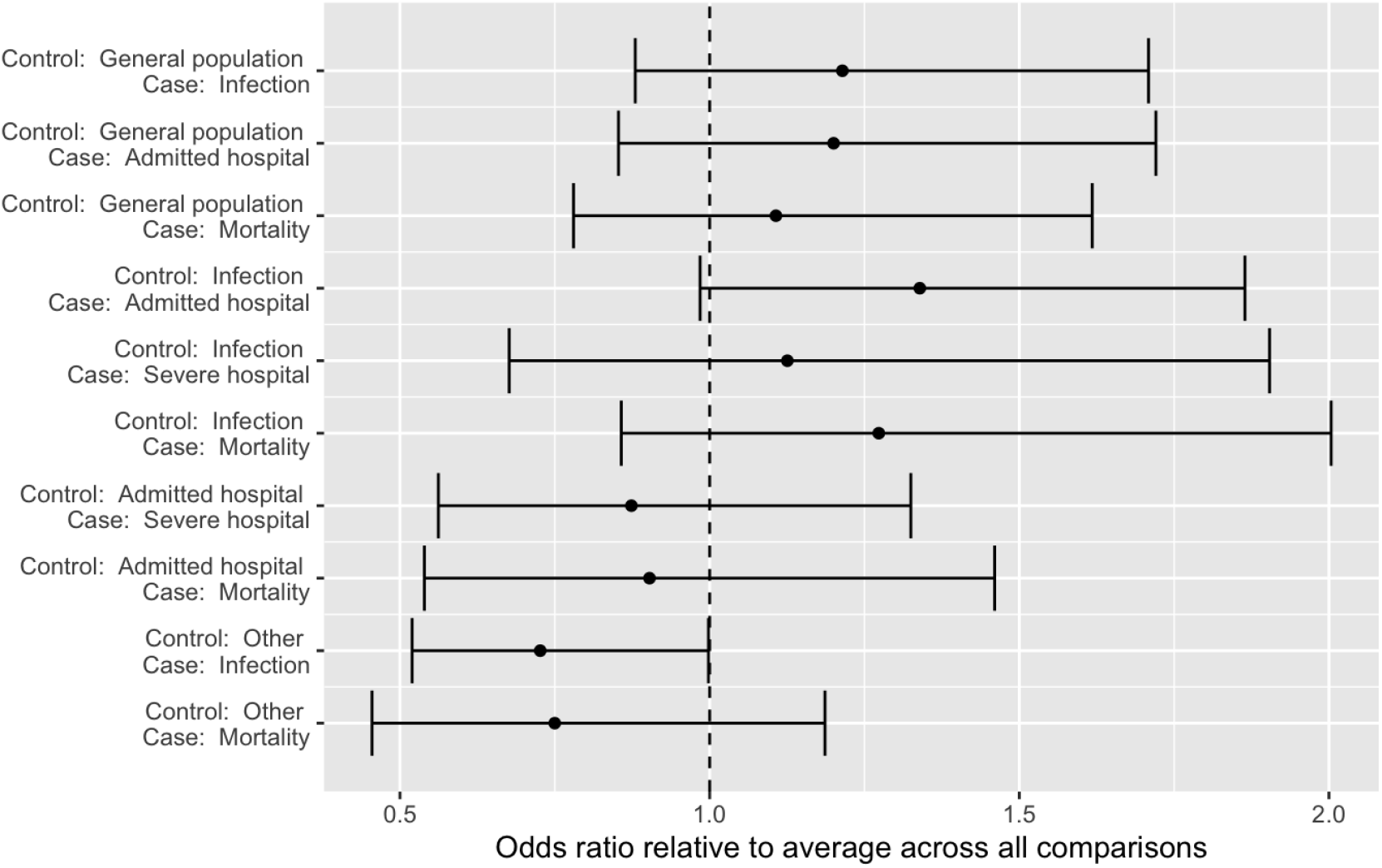
Differences across case and control outcome combinations. The plot shows average estimates and 95% credibility intervals for all combinations of case and control outcomes observed in the data. See supporting materials for model details.

## Discussion

Early research on Covid-19 has shown that the disease burden differs by SES, race and ethnicity (3-6). This is consistent with the results we report from the first systematic literature review on the associations between SES and disease outcomes in the last 5 influenza pandemics. We identified nine studies of the “Spanish flu of 1918-20” and 35 of the “Swine flu of 2009-2010”, but no studies of the “Russian flu” pandemic of 1889-90, the “Asian flu” of 1957-58 or the “Hong-Kong flu” of 1968-70. Most of the studies included for the 1918 and 2009 influenza pandemics used data from western high-income countries. Out of 51 estimates from 35 studies, the overall pooled mean pandemic outcome odds ratio was 1.44 (95% CI: 1.23 – 1.68) comparing the lowest to the highest SES groups. There was no evidence suggesting differences by pandemic period (1918 or 2009), the level of SES measure (individual or ecological), or type of method (odds ratio or relative risk). Finally, studies using healthy controls tended to find low SES associated with worse influenza outcome, and studies using infected controls find low SES associated with more severe influenza outcomes. Studies comparing severe outcomes (ICU or death) to hospital admissions were few but indicated no clear association. Studies with more unusual comparisons (e.g., pandemic vs seasonal influenza, seasonal influenza vs other patient groups) reported no or negative associations. These patterns were similar in a multivariate Bayesian model accounting for all study level indicators simultaneously. The Bayesian model also included indicators for study region/country. Relative to the “across all country/regions” average, studies from Australia, UK and to a lesser extent the USA tend to report stronger associations in our sample, while New Zealand tends to report weaker associations. These country-level results should be viewed as exploratory: two of the three studies from New Zealand (28, 29), for instance, are studies of how pandemic flu outcomes vary across pre-service occupational status amongst military personnel during the 1918 pandemic, which are unlikely to speak broadly to such associations in New Zealand more generally.

Our results provide strong evidence that social risk factors matter for pandemic flu outcomes in addition to medical risk factors. We also documented that in the 2009 pandemic, social risk factors independently explained variation in disease outcomes even when medical risk factors were controlled for (34, 46-48, 51, 54, 57, 59). This resembles the finding of a study of COVID-19 hospital deaths demonstrating that medical risk factors did little to explain the higher risks of the deprived and of immigrants in the UK (4). Although we did not find support for our hypothesis that social disparities would be larger for more severe (e.g. ICU and death) than less severe outcomes (e.g. infection or hospitalization not requiring ICU), the similarity of results for the 1918 and 2009 pandemics show the persistence of individual- and ecological-level social risk factors, although the specific mechanisms and types of social vulnerabilities leading to social disparities in pandemic outcomes may differ between 1918 and 2009, or in 2020 during the COVID-19 pandemic. Results from this review on pandemic influenza and results from studies on the role of social and ethnic vulnerability in COVID-19 disease outcomes (3-6), support recent calls for inclusion of social and ethnic vulnerabilities in addition to medical at risk factors in pandemic preparedness plans (19): Examples given are prioritizing of vaccines for medically vulnerable people living in socially vulnerable areas (urban slums or hard-to-reach groups in rural and remote areas), or SES groups with undiscovered medical vulnerability, and others who are at significantly higher risk of severe disease or death (various indigenous, ethnic, or racial groups, people living in extreme poverty, homeless and those living in informal settlements or; low-income migrant workers; refugees, internally displaced persons, asylum seekers, populations in conflict settings or those affected by humanitarian emergencies, vulnerable migrants in irregular situations and nomadic populations).

The studies reporting on social inequalities in influenza outcomes in 1918 and in 2009, identified in this review, and also early research on social disparities in COVID-19 outcomes, often lacked a discussion of the possible mechanisms for the estimated social disparities, a framework to discuss those mechanisms and/or the data to separate the distal (social and policy) and proximal (behavioral and biological factors) factors for unequal exposure, susceptibility and access to health care leading to socially unequal pandemic outcomes (68). Socially unequal exposure may relate to hand washing behavior or mask use, cleaning of surfaces, cramped living conditions, multigenerational living, occupational exposure, ability to work from home or stay away from work in order to care for family members and use of public transportation. Social disparities in susceptibility may relate to poor nutritional status or, concurrent illnesses (e.g. NCDs). Finally, socioeconomic inequalities in understanding of or access to health advice (e.g. hand hygiene, social distancing, travel advisories) and vaccination or other public recommendations due to poor reading and writing skills may also explain part of variation in outcomes by SES (14, 19).

Two of the studies on the 2009 pandemic included in our review, on Iran (35) and USA (55), reported increased risks for those with high socioeconomic status – contrary to the author’s hypothesis. For the US study, the authors suggest that this may reflect social gradients in testing and demand for treatment and health care resources.

An important strength of the study is the use of a pre-registered study protocol for data gathering and analysis, which was peer-reviewed and published prior to the gathering of study data (1). This helped ensure that the process was specified in a reproducible way and followed a rigorous and systematic workflow to identify studies and describe and analyze results. The engagement of professional information specialists to design, test and improve the literature search strategies that were applied to a broad range of literature databases is particularly important, given the lack of any previous systematic reviews on this topic with which our list of included studies could be compared.

Our study also has some potential limitations. First, we carried out our library search 17 November 2017, and potential studies published 2018-2020 are not included. Given the strength and consistency of the results, however, we do not expect that newer studies would alter our general conclusions, at least not for the 2009 pandemic that were the topic of 35 of the 44 included studies. Systematic reviews and meta-analysis of the associations between socioeconomic status/race/ethnicity and COVID-19 are also needed. Second, we would note that the generalizability of our results is necessarily limited by the geographic focus of the research we synthesize: no studies using data from Africa were found, and few from Asia and South America. It is therefore reasonable to ask whether our results are representative outside high-income countries in North America, Europe and Oceania.

## Conclusion

We have shown that influenza pandemic outcomes in 2009 were not always socially neutral «great equalizers» once you adjust for medical risk factors (34, 46-48, 51, 54, 57, 59). This resembles the finding of a study of COVID-19 hospital deaths demonstrating that medical risk factors did little to explain the higher risks of the deprived and of immigrants in the UK (69). The social lessons from historical influenza pandemics such as those in 1918 or 2009 have not yet been taken into account in influenza pandemic preparedness (19), and this blind spot has also been evident in the response to the COVID-19 pandemic. Such social and ethnic vulnerability factors should be explicitly included and addressed in current and future plans and responses in order to more effectively reduce pandemic burdens, reduce social disparities and ameliorate the social consequences of future pandemics (70). The global health and economic crisis created by the COVID-19 pandemic has made us only too aware of the need for a more holistic and comprehensive approach towards pandemic preparedness.

## Supporting information

1.Medline search strategy

2.PRISMA Checklist

3.PRISMA Flow Diagram

4.Specific studies included and all judgments and adjustments concerning inclusion and adjustments of reported numbers

## Data Availability

Data comes from a systematic review of the literature.

## Acknowledgements

This research is part of the project *PANRISK: Socioeconomic risk groups, vaccination and pandemic influenza*, funded by a research grant from the Research Council of Norway (grant agreement No. 302336). We are indebted to our librarians Bettina Grødem Knutsen, Ingjerd Legreid Ødemark and Elisabeth Karlsen at the Learning Center and Library, Oslo Metropolitan University. Without their expertise and assistance in doing library searches this research would never have been accomplished.

## Supporting information files

1. Medline search strategy
2. PRISMA Checklist
3. PRISMA Flow Diagram
4. Specific studies included and all judgments and adjustments concerning inclusion and adjustments of reported numbers.

## Footnotes

1 Reasons for these **117** exclusions were the following: **50** No control for socioeconomic factors in addition to biological risk factors; **18** No control for SES in a study of ethnic groups and biological risk factors; **15** No control for SES in addition to ethnic groups; **15** No control for socioeconomic confounders; **4** Wrong patient population; **4** Wrong time period; **2** No quantitative data; **2** wrong language; **1** Duplicate; **1** Spanish language; **1** Studies on both seasonal and pandemic influenza that do not distinguish between non-pandemic and pandemic years; **1** Reason not given; **1** Wrong intervention; **1** Wrong outcomes; **1** Wrong study design.

2 Reasons for these **15** exclusions were the following: **5** wrong intervention; **1** Studies on both seasonal and pandemic influenza that do not distinguish between non-pandemic and pandemic years; **4** Duplicates; **1** No control for socioeconomic confounders; **2** Wrong outcomes; **1** Wrong time period; **1** Wrong study design.

3 See supporting materials for model code and discussion of prior choices.

4 See supporting materials for model code and discussion of prior choices.

