## Supplementary material for "The association between socioeconomic status and pandemic influenza: systematic review and meta-analysis": 1.Medline search strategy

| # | Searches |
|---|---|
| 1 | Orthomyxoviridae/ |
| 2 | Influenzavirus A/ |
| 3 | Influenza A virus/ |
| 4 | Influenza A Virus, H1N1 Subtype/ |
| 5 | Influenza A Virus, H1N2 Subtype/ |
| 6 | Influenza A Virus, H2N2 Subtype/ |
| 7 | Influenza A Virus, H3N2 Subtype/ |
| 8 | Influenza A Virus, H3N8 Subtype/ |
| 9 | Influenza Pandemic, 1918-1919/ |
| 10 | Pandemics/ |
| 11 | Disease Outbreaks/ |
| 12 | 1 or 2 or 3 or 4 or 5 or 6 or 7 or 8 |
| 13 | 10 or 11 |
| 14 | 12 and 13 |
| 15 | ((Influenza or flu) adj3 (pandemic* or outbreak* or epidemic*)).tw,kw,kf. |
| 16 | ((Russian or Spanish or Asian or Hong Kong or Mexican) adj3 (flu or influenza or pandemic* or epidemic*)).tw,kw,kf. |
| 17 | ((H1N1 or H1N2 or H2N2 or H3N2 or H3N8) adj3 (pandemic* or outbreak* or epidemic*)).tw,kw,kf. |
| 18 | (PH1N1 or H1N1pdm09 or H1N1p).tw,kw,kf. |
| 19 | (("1889" or 1889-90 or 1889-1890 or 1889-91 or 1889-1891 or 1889-92 or 1889-1892 or 1889-93 or 1889-1893 or 1889-94 or 1889-1894 or 1918-19 or 1918-1919 or 1918-20 or 1918-1920 or "1957" or "1958" or 1957-58 or 1957-1958 or "1968" or 1968-69 or 1968-1969 or 1968-70 or 1968-1970 or "1969" or 1969-70 or 1969-1970 or "1970" or "2009" or 2009-10 or 2009-2010) adj7 (flu or influenza or pandemic* or epidemic* or outbreak*)).tw,kw,kf. |
| 20 | 9 or 14 or 15 or 16 or 17 or 18 or 19 |
| 21 | Socioeconomic Factors/ |
| 22 | Poverty/ |
| 23 | Poverty Areas/ |
| 24 | Working Poor/ |
| 25 | Social Class/ |
| 26 | Social Conditions/ |
| 27 | Social Marginalization/ |
| 28 | Social Isolation/ |
| 29 | Educational Status/ |
| 30 | Employment/ |

| | |
|---|---|
| 31 | Medical Indigency/ |
| 32 | Medically Uninsured/ |
| 33 | "Social Determinants of Health"/ |
| 34 | Vulnerable Populations/ |
| 35 | Minority Groups/ |
| 36 | "Transients and Migrants"/ |
| 37 | Housing/ |
| 38 | Crowding/ |
| 39 | Indians, North American/ |
| 40 | Indians, Central American/ |
| 41 | Indians, South American/ |
| 42 | Inuits/ |
| 43 | Alaska Natives/ |
| 44 | Oceanic Ancestry Group/ |
| 45 | Refugees/ |
| 46 | "Emigrants and Immigrants"/ |
| 47 | "Emigration and Immigration"/ |
| 48 | Ethnic Groups/ |
| 49 | Demography/ |
| 50 | (((social* or socioeconomic* or socio-economic* or sociodemographic* or socio-demographic* or socioecologic* or socio-ecologic* or economic*) adj3 (factor* or condition* or aspect* or impact* or indicator* or indice* or index* or disparit* or difference* or depriv* or inequalit* or justice or injustice or determin* or class* or status)) or SES).tw,kw,kf. |
| 51 | (Gini adj1 (coefficient or ratio or index)).tw,kw,kf. |
| 52 | (health adj3 (inequalit* or difference* or disparit*)).tw,kw,kf. |
| 53 | (living condition* or poverty or material deprivation* or crowding or housing or employment or unemployment or income* or wealth or social vulnerability or ((public or social) adj3 (welfare or security)) or working class or slum* or homelessness or homeownership or apartment size).tw,kw,kf. |
| 54 | ((poor or vulnerable or underprivileged or disadvantaged or homeless or remote) adj3 (people or person* or group* or communit* or parish* or neighbourhood* or countr* or population* or area*)).tw,kw,kf. |
| 55 | (literacy or illiteracy or educat*).tw,kw,kf. |
| 56 | (Nativ* or immigrant* or refugee* or migrant* or minorit*).tw,kw,kf. |
| 57 | (indigenous or ethnic or American Indian* or Alaska native* or aborigin* or maori or inuit* or Yupik or Cupik or Inupiaq or Saint Lawrence Island Yupik or Unangax or Alutiiq or Eyak or Tlingit or Haida or Tsimshian or sami or eskimo* or Torres strait islander* or pacific islander* or pacific people or athabascan).tw,kw,kf. |
| 58 | or/21-57 |
| 59 | 20 and 58 |

| | |
|---|---|
| 60 | limit 59 to (danish or english or norwegian or swedish) |
