## Supplementary material for "The association between socioeconomic status and pandemic influenza: systematic review and meta-analysis": 3.PRISMA Flow Diagram

### PRISMA 2009 Flow Diagram

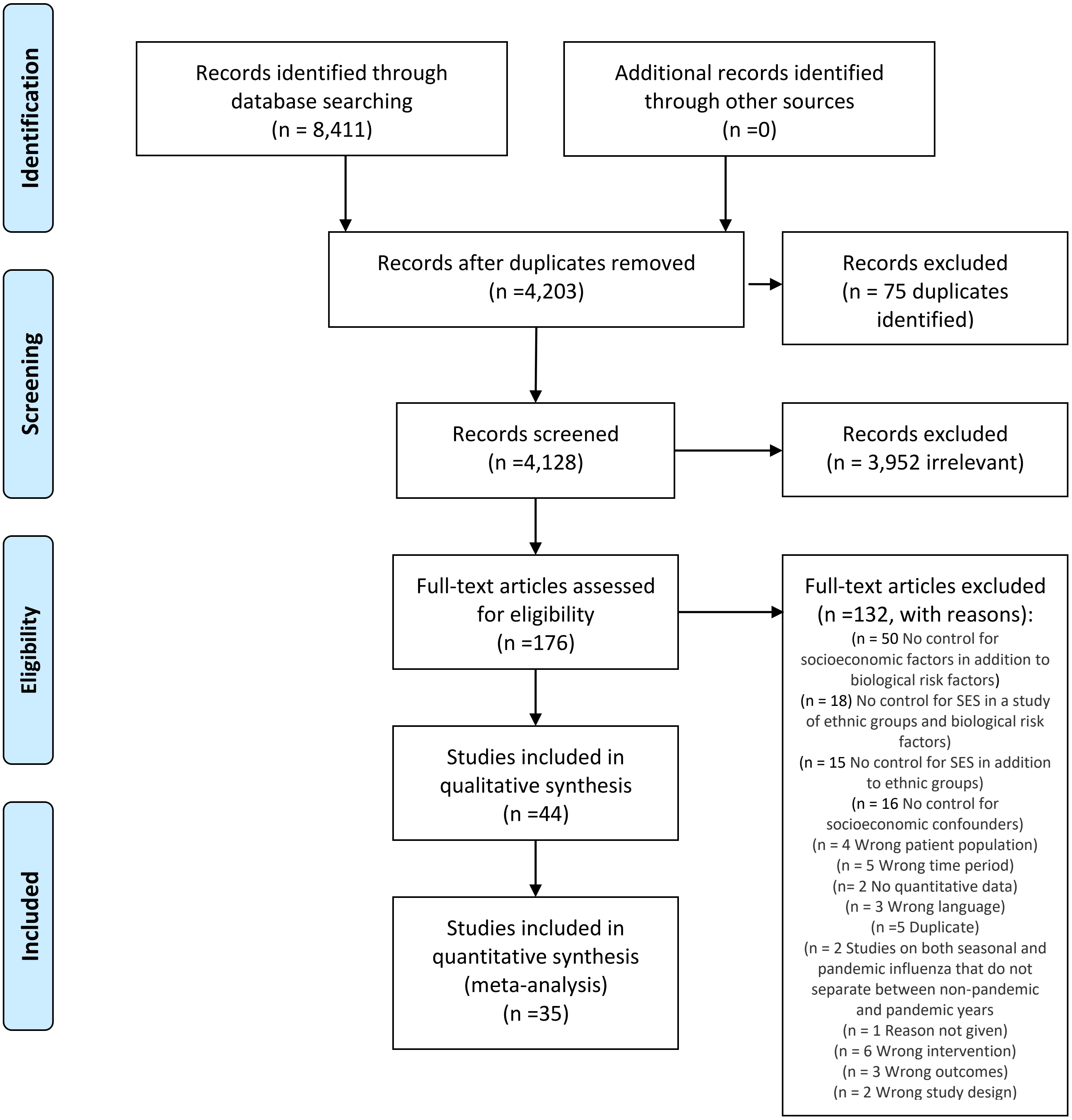


*From:* Moher D, Liberati A, Tetzlaff J, Altman DG, The PRISMA Group (2009). *P*referred *R*eporting *I*tems for *S*ystematic Reviews and *M*eta-*A*nalyses: The PRISMA Statement. PLoS Med 6(7): e1000097. doi:10.1371/journal.pmed1000097

**For more information, visit www.prisma-statement.org.**
