## Supplementary material for "The association between socioeconomic status and pandemic influenza: systematic review and meta-analysis": 4.Specific studies included and all judgments and adjustments concerning inclusion and adjustments of reported numbers

### Meta-analysis - quantitative part

#### Introduction

This note documents the judgments and calculations made during the quantitative analyses performed as part of the meta-analysis of studies assessing the association between indicators of socio-economic status and different pandemic related outcomes (e.g., infections, hospitalizations, deaths).

As a result of the Covid-19 outbreak and the increased interest for our study, we decided to focus on the established meta-analytic techniques and finish the paper based on these.

#### Analysis plan

##### Initial

The original plan was to analyze the data in three ways:

1. A random effects meta-analysis
2. A PET-PEESE meta-regression, using study level variables as explanatory covariates
3. Bayesian model estimating "dose-response" gradients

This is based on the published pre-analysis plan, which included the following section on how we planned to perform the quantitative analysis of the gathered data:

> The quantitative part of the study will pool results across studies. Such pooling can be done using various methods that impose different constraints on the type of studies that can be pooled. We will pursue three strategies. The first two are within the frequentist statistical tradition. We will here note whether coefficients are statistically significant at the 1%, 5%, or 10% level, i.e., whether the evidence base indicates pooled effects that would be unlikely if the true effect was zero. We will also discuss the strength of this test by assessing the magnitude of the pooled coefficient and its standard error (precision) in relation to plausible effect sizes. In the third, Bayesian methods will be used and we will assess how the evidence updates weakly informative priors for the coefficients.
>
> *Pooled effect meta-analysis* Where several studies are available with similar outcome and exposure measures, we will show forest plots and estimate pooled effects using fixed and random effects analyses with the metafor meta-analytic package in R [20], transforming the outcome variable when this is required to make the sampling distribution approximate the normal distribution, e.g., taking the log of odds ratios or using the Freeman-Tukey double arcsine transformation for proportions. In these analyses, the pooled estimates will reflect comparisons of the highest to the lowest reported socioeconomic group. We expect random effects to be more appropriate, since the socioeconomic gradient in outcomes may differ across time and region (e.g., we would expect a lower gradient in countries and periods with lower inequality). Cochran's Q test will be used to assess whether data indicate statistically significant heterogeneity in effects at the 5% level. Effect estimates are also expected to differ systematically across studies according to the socioeconomic "distance" between compared groups. For instance, we would expect a larger outcome difference between the top and bottom 10% of a distribution than we would between those above and below

the mean. Depending on the total number of studies that can be pooled in a given analysis, it may also be appropriate to conduct subsample analyses that assess whether pooled effects differ within subgroups of studies characterized by region, pandemic, age-group, gender, and estimation technique or quality assessment score.

*Meta-regression* A recent innovation in meta-analyses is a meta-regression technique with precision effect test and precision effect estimate with standard error ("PET-PEESE") [21, 22]. This technique will be used to pool estimates with similar outcome measures and will allow us to include study-level information as covariates and explore how these correlate with the coefficient estimates. This allows us to assess whether coefficients from comparisons of educational groups tend to differ from those comparing income groups, whether coefficients vary systematically by study-level variables such as pandemic, country-level inequality measures, statistical methodology used, or quality assessment score. The method additionally allows for the examination of how estimates differ systematically with e.g., age-groups. Finally, the technique tests whether coefficients vary systematically with reported standard errors, which may indicate the presence of small sample or publication bias.

*Bayesian meta-analysis* The above strategies require a similar outcome measure and will pool coefficients for the highest relative to the lowest socioeconomic group from each study. This ignores the "dose-response" information available from studies that report coefficients comparing multiple socioeconomic levels to a reference level (e.g., coefficients for different income quantiles). Under the assumption that an underlying socioeconomic gradient will be linear on the logit scale, all such reported estimates can contribute to estimating the underlying gradient [23, 24]. The resulting statistical model will be coded and estimated using the Stan language for probabilistic modeling [25] with a multilevel/hierarchical specification to account for heterogeneity across exposure measures (e.g, income, education), pandemic, and study-level covariates. We will also explore whether such an approach makes it feasible to pool studies across outcome measures to assess the hypothesis that gradients vary systematically by the severity of outcome.

#### Amended

Due to the Covid-19 outbreak and the strong interest in this study, as well as the large heterogeneity in outcomes and indicators used across studies, it was decided to simplify the quantitative analysis and prioritize standard meta-analytic analyses and a set of comparisons across studies of different types (e.g., those dealing with the 1918 vs those dealing with the 2009 pandemic).

Two approaches were used:

1. Using the R Metafor meta-analytic package, we estimated random effect models on the total sample of studies and on splits across different subsample dimensions
2. Using the Stan programming language for probabilistic modelling, we estimated a hierarchical model that included parameters for different subsample characteristics in a joint analysis.

#### Study level characteristics defining meta-analytic subgroups

1. *Deprivation measure: ecological, individual level.* Does the study use information on the level of individuals (e.g., self-reported income) or does it proxy individual characteristics (e.g., neighborhood poverty levels)?
2. *Case-criterion: Infected, admitted hospital, severe hospital, mortality*
3. *Control-criterion: General population, infected, admitted hospital, sever hospital, other* (This would refer to the control sample in a case control or the at-risk population in other studies). General population should not be taken to mean "population representative" - this category is also used to cover other studies where the controls are an appropriate non-infected sample from the population the cases are selected from (e.g., military personnel).

4. *Period: 1918, 2009*
5. *Country/region* Which country or multi-national region was studied
6. *Type of estimate reported: odds ratio, relative risk*

The case and control defining criteria are included to help inform an answer two questions central in this project: 1. Are flu related outcomes (infections, hospitalizations, death) more or less common in groups and individuals with lower socioeconomic status. 2. Is there a progressively increasing gradient with severity, such that the over-representation of low-SES individuals is stronger for hospitalizations than for infections, stronger for severe hospital treatments than for hospital admissions, and stronger for death than for severe outcomes.

### Study level data - documentation

We begin by constructing a data-set, labelled meta_df, appropriate to a random-effects analysis. We include studies that allow us to express the relative risk or relative rate of some flu-related outcome (infection, hospitalization, death) in a group with low relative to a high socio-economic indicator.

For each study, we include either a) the estimate and its upper and lower bound, or b) the estimate and its standard error. The data frame has the following columns:

1. Study number
2. odds ratio
3. lower bound
4. upper bound
5. standard error (log)

In a separate data frame, we also add information needed for the subsample analyses.

The data was extracted from the articles in question by Ole Rogeberg, and compared to estimated extracted and compiled into an Excel data sheet by Svenn Erik Mamelund and Clare Shelley-Egan. The resulting documentation (this note) was consequently read and controlled again by SEM, and discrepancies resolved.

In several cases, multiple estimates were available from the same study using different methods or different indicators of socio-economic status. To avoid giving undue weight to single studies, we did not include estimates from multiple univariate analyses using different SES-indicators from the same sample. Individual level indicators were preferred over ecological if both were available, and all else equal income > education > other indicators.

If multiple estimates from distinct sub-groups were estimated and a combined estimate could be used instead, the full sample estimate was used. In other cases, if a study included estimates for different periods, both were included. Likewise, if e.g., two age groups could not be combined, they would be entered separately.

When multivariate estimates are available, these were preferred and the most direct estimate of economic deprivation was used. Note that the use of adjusted estimates could potentially be an issue if some studies "overcontrol" in the sense of adding controls on the "causal path" from SES or deprivation to outcomes. If a researcher were to estimate the "effect" of poverty after controlling for diet, dwellings, health related behaviors, etc., at some point the baby would be thrown out with the bath-water. Note that the comparison of odds ratios across samples and models is inherently problematic in some ways.

For each study we include the study's identifier (from the Excel sheet with extracted data), the odds-ratio with lower and upper bounds, the standard error, and the other information.

```
meta_df <- data.frame(study_index = numeric(),
                      or = numeric(),
                      lb = numeric(),
                      ub = numeric(),
                      log_se = numeric())
```

For each study, we also add information required for the subsample analyses:

```
study_df <- data.frame(study_index = numeric(),
                       deprivation_type = character(),
                       case_crit = character(),
                       control_crit = character(),
                       period = integer(),
                       country = character(),
                       method = character(),
                       stringsAsFactors = FALSE)
```

##### Study 1

Aligne 2016

Not relevant: Crowding in this military setting is not an indicator of individual SES

##### Study 2

Balasegaram 2012

*Deprivation measure:* Ecological (postcode of residence mapped to Index of Multiple Deprivation)

*Case-criterion:* Infection ("cases of pandemic flu [. . . ] reported to the London Flu Response Centre")

*Control-criterion:* General population

*Period:* 2009

*Country/region:* London, UK

*Type of estimate reported:* relative risk

The estimates are found on page e38 - we use the estimates that pools all age groups and all weeks. They are based on the 81% of cases with valid postcode.

Contains estimates of all ages, comparing areas grouped by levels of deprivation. Also contains sub-group analyses for different age groups and outcomes at different time. We use the "all ages" estimate, using the most affluent group as the reference.

```
meta_df[1,] <- c(2,
                 2.32,
                 1.94,
                 2.78,
                 NA_real_)

study_df[1, ] <- c(2,
                 "Ecological",
                 "Infection",
                 "General population",
                 2009,
                 "UK",
                 "relative risk")
```

##### Study 3

Balter 2010

*Deprivation measure:* Ecological ("We defined neighborhoods using the United Hospital Fund (UHF) designation, which aggregates adjoining ZIP codes to create 42 NYC neighbor- hoods (12). We then created a neighborhood poverty variable by categorizing UHF neighborhoods into tertiles (low-, medium-, and high-poverty neighborhoods) based on the percentage of residents living <200% of the federal poverty level, according to the US Census 2000")

*Case-criterion:* "Admitted hospital ("We analyzed surveillance data to describe NYC residents who were hospitalized with pandemic (H1N1) 2009 in NYC from the start of the first ICS activation to the end of the second activation (April 24–July 7).) Note that this also includes people hospitalized for other reasons found to have H1N1.

*Control-criterion:* General population

*Period:* 2009

*Country/region:* New York, USA

*Type of estimate reported:* odds ratio

This study has two estimates listed - one adjusting for age and one not. Neither has standard errors or confidence intervals. We consequently use the study information to calculate an odds-ratio.

The study lists 498 high-poverty events and 172 low poverty events. These are used as case-counts.

To get control counts: The study notes that 0.327 and 0.264 were the respective shares of high and low poverty individuals in NYC based on the 2000 NYC census. An online search found the total NYC population in 2000 to be at 8 008 278 individuals. This is used to get counts for the high and low poverty groups in the full NYC population, which we use as control counts.

Using this information, we can calculate an odds-ratio with a standard error:

```
cases <- c(498, 172)
controls <- c(0.327, 0.264) * 8008278 - cases

or <- (cases[1]/controls[1])/(cases[2]/controls[2])
cat("Odds ratio: ", or, "\n")
```

```
## Odds ratio:  2.337784
```

```
log_or_se <- sqrt(sum(1/c(cases, controls)))
cat("Log_or_se: ", log_or_se)
```

```
## Log_or_se:  0.08844682
```

```
meta_df <- rbind(meta_df,
                 c(3,
                   or,
                   exp(log(or) - 1.96 * log_or_se),
                   exp(log(or) + 1.96 * log_or_se),
                   log_or_se))
study_df[2, ] <- c(3,
                   "Ecological",
                   "Admitted hospital",
                   "General population",
                   2009,
                   "USA",
                   "odds ratio")
```

##### Study 4

Bandaranayak 2010

*Deprivation measure:* individual - ("reported damp housing")

*Case-criterion:* Infected (assessed after pandemic by measuring antibodies)

*Control-criterion:* General population (stratified random sample with stratified and weighted analysis)

*Period:* 2009

*Country/region:* New Zealand

*Type of estimate reported:* odds ratio

The estimates are found in table 3

```
meta_df <- rbind(meta_df,
                 c(4,
                   1.1,
                   0.83,
                   1.1,
                   NA_real_))
study_df[3, ] <- c(4,
                   "Individual",
                   "Infection",
                   "General population",
                   2009,
                   "New Zealand",
                   "odds ratio")
```

##### Study 5

Chandrasekhar 2017

This study includes data from 2009-10 to 2013-2014. Exclusion reason: It combines seasonal and pandemic influenza across non-pandemic and pandemic years.

##### Study 6

Cheraghi 2010

*Deprivation measure:* Individual - Education

*Case-criterion:* Infected

*Control-criterion:* Other ("Subjects (cases and controls) were selected from those patients with signs and symptoms of respiratory tract infection who referred to health centers of eight cities throughout Hamedan Province, western Iran form July to December 2009.")

*Period:* 2009

*Country/region:* Iran

*Type of estimate reported:* odds ratio

This means that the controls are not representative of the general population. The interpretation of the estimates would have to be how the odds that someone with signs and symptions of respiratory tract infection actually had H1N1, as a function of deprivation.

Study of education. The reference category is low education so I take the reciprocal to reverse. The estimates are given in the abstract.

```
meta_df <- rbind(meta_df,
                 c(6,
                   1/1.84,
                   1/2.86,
                   1/1.32,
                   NA_real_))

study_df <- rbind(study_df,
                  c(6,
                    "Individual",
                    "Infection",
                    "Other",
                    2009,
                    "Iran",
                    "odds ratio"))
```

##### Study 7

Chowell 2008

This study is part of the narrative analysis, but data cannot be used in the meta-analysis, as it only reports correlations with p-values.

##### Study 8

Chowell 2014

Four estimates for different waves, with population density (persons/km^2) as the contrast. None of the socieconomic measures are credible indicators of poverty.

##### Study 9

Dawood 2012 Region not very specific as SES measure.

##### Study 10

Duggal 2016 Comparisons of high, upper middle and lower middle income economy (country level).

This is a meta-analysis - which makes it problematic to include, as it will reflect evidence already included from individual studies.

##### Study 11

Fajardo-Dolci 2012

*Deprivation measure:* Individual (education)

*Case-criterion:* Mortality

*Control-criterion:* Other (mortality from non-pandemic influenza)

*Period:* 2009

*Country/region:* Mexico

*Type of estimate reported:* odds ratio

These estimates are found in table 3

```
meta_df <- rbind(meta_df,
                 c(11,
                   1/1.651,
                   1/3.559,
                   1/1/0.766,
                   NA_real_))

study_df <- rbind(study_df,
                  c(11,
                    "Individual",
                    "Mortality",
                    "Other",
                    2009,
                    "Mexico",
                    "odds ratio"))
```

#### Study 12

Gilca 2011

This one provided two separate estimates: One comparing hospitalized to non-hospitalized cases, and one comparing severe (ICU or death) vs non-severe hospital cases.

##### Estimate 1

Compares people with verified H1N1 tracked in the confirmed case registry to those in hospital - using phone interviews to get information on the community cases.

*Deprivation measure:* Individual - Education (collected at the individual level in a survey)

*Case-criterion:* Hospitalized

*Control-criterion:* Verified H1N1 outside of hospital system

*Period:* 2009

*Country/region:* Canada

*Type of estimate reported:* odds ratio

These estimates are found in table 2 - column 2

```
meta_df <- rbind(meta_df,
                 c(12,
                   1.3,
                   0.7,
                   2.4,
                   NA_real_))

study_df <- rbind(study_df,
                  c(12,
```

```
                     "Individual",
                     "Admitted hospital",
                     "Infection",
                     2009,
                     "Canada",
                     "odds ratio"))
```

###### Estimate 2

Compares people with ICU or death (severe cases) to non-severe cases in hospital.

*Deprivation measure:* Individual - Education (collected at the individual level in a survey)

*Case-criterion:* Severe hospital

*Control-criterion:* Admitted hospital (non-severe)

*Period:* 2009

*Country/region:* Canada

*Type of estimate reported:* odds ratio

These estimates are found in table 2 - column 4

```
meta_df <- rbind(meta_df,
                c(12.5,
                  1.5,
                  0.4,
                  4.1,
                  NA_real_))




study_df <- rbind(study_df,
                 c(12.5,
                   "Individual",
                   "Severe hospital",
                   "Admitted hospital",
                   2009,
                   "Canada",
                   "odds ratio"))
```

#### Study 13

Gonzalex-Candelas

*Deprivation measure:* Individual - Own education - gathered using phone interviews

*Case-criterion:* Admitted to hospital with >24 hours stay due to confirmed H1N1

*Control-criterion:* Non-hospitalized persons with confirmed infection attending primary care centers

*Period:* 2009

*Country/region:* Spain

*Type of estimate reported:* odds ratio

The estimates are found in table 2 - I take the reciprocal to make the highest education the reference.

```
meta_df <- rbind(meta_df,
                 c(13,
                   1/0.44,
                   1/0.63,
                   1/0.31,
                   NA_real_))

study_df <- rbind(study_df,
                  c(13,
                    "Individual",
                    "Admitted hospital",
                    "Infection",
                    2009,
                    "Spain",
                    "odds ratio"))
```

#### Study 14

Grantz 2016

*Deprivation measure:* Ecological (census tract-level data on demographic characteristics in 1920)

*Case-criterion:* Death from influenza

*Control-criterion:* General population

*Period:* 1918

*Country/region:* Chicago, US

*Type of estimate reported:* relative risk

Table 1 of this paper includes the results of a Poisson model estimating pandemic influenza mortality and its associations with various neighborhood characteristics. Using percentage illiterate as the indicator of poverty and the multivariate regression adjusted RR, we have a relative risk of 1.028 (1.020, 1.036). Figure 2 indicates that the illiteracy rate across districts vary from 0-7 (lowest) to 21-28 (highest). This gives an approximate extreme-to-extreme range of 20 percentage points. To avoid extremes where the linear relationship may break down we use the numbers in the text: "for every 10% increase in illiteracy rate within a given census tract, mortality increased by 32.2% (95% CI: 22.2, 43)."

We interpret the 10% increase as a 10 percentage point increase, as this fits with the numbers: If 1 percentage point increases with 1.028, then 10 would increase it by $(1.028)^\wedge 10 = 1.318$ which seems close enough that their number is calculated the same way but with more decimals.

```
meta_df <- rbind(meta_df,
                 c(14,
                   1.322,
                   1.222,
                   1.43,
                   NA_real_))

study_df <- rbind(study_df,
                  c(14,
                    "Ecological",
                    "Mortality",
                    "General population",
```

```
                    1918,
                    "USA",
                    "relative risk"))
```

#### Study 15

Hennessy 2016

*Deprivation measure:* Individual - gathered using interview (25% with missing on income - imputed)

*Case-criterion:* Mortality

*Control-criterion:* Infected who were not hospitalized for 30 days after specimen collection

*Period:* 2009

*Country/region:* USA (selected states)

*Type of estimate reported:* odds ratio

The estimates are found in table 2

This study has several indicators of poverty (education, insurance, crowded dwellings and poverty directly). I use the direct poverty resukt only - including several coefficients would give this study excessive influence on the pooled estimate, as the variables are correlated and results given for univariate analyses.

In addition, the study has a second set of results for a subsample (Alaskan Native/American Indian), but these data are also included in the full analysis used here.

NOTE: The sheet states that none of the variable were significant in a multivariate analysis - if the multivariate analysis simply included these proxies at the same time this is likely due to multicollinearity, i.e., a situation where the variables are collectively significant but correlated to such an extent that it is hard to quantify their individual contribution.

```
meta_df <- rbind(meta_df,
                 c(15,
                   3.41,
                   1.57,
                   7.41,
                   NA_real_))

study_df <- rbind(study_df,
                  c(15,
                    "Individual",
                    "Mortality",
                    "Infection",
                    2009,
                    "USA",
                    "odds ratio"))
```

#### Study 16

Hoen 2010 This study compares schools with media-reports of H1N1 influenza to nearby schools not mentioned in the media, i.e. estimating the probability that a school has "confirmed cases of novel H1N1 influenza that are picked up by the media and detected by HealthMap." This means that the associations are a mix of differential prevalence and varying media interest concerning infections for rich and poor schools.

*Deprivation measure:* Ecological - Title-1 funding status of attended school (indicates more economically disadvantaged)

*Case-criterion:* US Schools with confirmed cases of H1N1 influenza mentioned in english-language media reports (32 schools)

*Control-criterion:* Other schools in the community of the cases (6815 schools)

*Period:* 2009

*Country/region:* US

*Type of estimate reported:* odds ratio

The multivariate model estimates are used, found in table 1

```
meta_df <- rbind(meta_df,
                 c(16,
                   0.385,
                   0.166,
                   0.894,
                   NA_real_))

study_df <- rbind(study_df,
                  c(16,
                    "Ecological",
                    "Infection",
                    "Other",
                    2009,
                    "USA",
                    "odds ratio"))
```

#### Study 17

Hu 2012 This is a study of how incidence rates varied with lagged rainfall and weather, as well as the socioeconomic status of different areas.

The socioeconomic area index (SEIFA) ranges from 800 to 1200 with a mean (sd) of 1064 (70), and the coefficient on SEIFA (on the logarithmic scale) is estimated at -0.06 (-0.19 to 0.06). We did not see how the results from this paper could be translated to a metric comparable to those used in other studies.

#### Study 18

Huang 2016 This is an estimated SIR infection model. We did not see how the estimates using socioeconomic index could be converted to something that could be included in the meta-analysis.

#### Study 19

Inglis 2014 This study reports the number of cases from regions grouped into different quintiles on the basis of deprivation scores. Out of 2978 total confirmed cases, 1837 cases came from the lowest and 170 from the highest, with 971 from the remaining quintiles.

This study was not included in the quantitative analysis. We would need to know the extent to which cases were found in the "high deprivation" areas relative to what we would expect in the "no effect" counterfactual where cases would be proportional to the population in each quintile. However, we do not know the size of the different populations residing in each "area quintile".

As far as we could tell from the paper, however, the quintiles are quintiles of *areas*, not the population. To put it simply: consider a case where there are two neighborhoods - one crowded urban neighborhood where everyone poor is crammed into small apartments, and one sparsely populated rural area with manors and castles. We would expect more people from the first region, which would be "high deprivation", even if there were no association with SES.

#### Study 20

Inglis 2013 Marked as duplicate of 19

#### Study 21

Janjua 2012

This study compares influenza incidence across parents with children community schools of a rural BC community

*Deprivation measure:* Individual - Household density collected by phone

*Case-criterion:* Infected (antibodies tested on people self-reporting symptons in phone survey)

*Control-criterion:* General population

*Period:* 2009

*Country/region:* Canada

*Type of estimate reported:* odds ratio

The estimates are found in table 1, multivariable model. We assume that 1st to 3rd quantiles (reference) of household density have the lowest density, i.e., highest SES, and that the fourth quantile is the one with the highest density (lowest SES).

```
meta_df <- rbind(meta_df,
                 c(21,
                   1.17,
                   0.6,
                   2.28,
                   NA_real_))

study_df <- rbind(study_df,
                  c(21,
                    "Individual",
                    "Infection",
                    "General population",
                    2009,
                    "Canada",
                    "odds ratio"))
```

#### Study 22

Kumar 2012 Marked as not relevant

##### Study 23

Launes 2012

*Deprivation measure:* Individual (Parental education level)

*Case-criterion:* Hospitalized ("patients aged 6 months to 18 years hospitalized for influenza syndrome")

*Control-criterion:* Infected ("patients aged 6 months to 18 years with confirmed influenza A (H1N1) 2009 infection using real-time RT-PCR and man- aged on an outpatient basis.")

*Period:* 2009

*Country/region:* Spain

*Type of estimate reported:* odds ratio

The estimates are found in Table 1

```
meta_df <- rbind(meta_df,
                 c(23,
                   2.7,
                   1.4,
                   5.2,
                   NA_real_))

study_df <- rbind(study_df,
                  c(23,
                    "Individual",
                    "Admitted hospital",
                    "Infection",
                    2009,
                    "Spain",
                    "odds ratio"))
```

##### Study 24

Lenzi 2012

*Deprivation measure:* Individual - level of education (literate vs illiterate)

*Case-criterion:* Admitted hospital (marked as hospitalized in National Case Registry Database)

*Control-criterion:* Infected (marked as non-hospitalized in National Case Registry Database)

*Period:* 2009

*Country/region:* Brazil

*Type of estimate reported:* odds ratio

The estimates are found in table 3 (the illiterate are set as reference, so we take the reciprocal)

```
meta_df <- rbind(meta_df,
                 c(24,
                   1/0.815,
                   1/0.917,
                   1/0.724,
                   NA_real_))

study_df <- rbind(study_df,
```

```
                  c(24,
                    "Individual",
                    "Admitted hospital",
                    "Infection",
                    2009,
                    "Brazil",
                    "odds ratio"))
```

##### Study 25

Lenzi 2011 No quantitative estimates

##### Study 26

Levy 2013

*Deprivation measure:* Individual (telephone interview)

*Case-criterion:* Admitted hospital

*Control-criterion:* Infected (Non-hospitalized lab-confirmed influenza A patients)

*Period:* 2009

*Country/region:* USA

*Type of estimate reported:* odds ratio

The estimates are found in Model 3 in table 3, which adjusts for underlying conditions, insurance and access to care and for Bronx (from where cases were likely oversampled). We use the educational level and risk of hospitalization results.

Has estimates for education, % below the poverty line and household income, each of these for adults and children respectively.

Income data were only reported by 60% of participants, so neighborhood SES was added using percentage below poverty in neighborhood (using zip-codes and census poverty data)

```
meta_df <- rbind(meta_df,
                 c(26,
                   16.81,
                   4.27,
                   66.13,
                   NA_real_))

study_df <- rbind(study_df,
                  c(26,
                    "Individual",
                    "Admitted hospital",
                    "Infection",
                    2009,
                    "USA",
                    "odds ratio"))
```

#### Study 27

Lowcock 2011 Just abstract - published as study 28

#### Study 28

Lowcock 2012

Has estimates from two phases of the pandemic - "phase 1" (April 23-July 20 2009) and phase 2 (August 1 Nov 6 2009).

*Deprivation measure:* Individual (educational level collected through interview)

*Case-criterion:* Hospitalized (self-reported)

*Control-criterion:* Infected

*Period:* 2009

*Country/region:* Canada

*Type of estimate reported:* odds ratio

The estimates are found in table 2.

We include four estimates from different subpopulations (phase x age_group) as children are assigned parental education

```
# Adults phase 1
meta_df <- rbind(meta_df,
                c(28,
                  2.28,
                  1.13,
                  4.59,
                  NA_real_))
# Adults phase 2

meta_df <- rbind(meta_df,
                c(28,
                  1.77,
                  1.08,
                  2.89,
                  NA_real_))

# Children phase 1
meta_df <- rbind(meta_df,
                c(28,
                  0.98,
                  0.41,
                  2.35,
                  NA_real_))
# Children phase 2
meta_df <- rbind(meta_df,
                c(28,
                  1.32,
                  0.79,
                  2.21,
                  NA_real_))
```

```r
study_df <- rbind(study_df,
                  c(28,
                    "Individual", # Own education (those above 18) - parental (those below)
                    "Admitted hospital", # Hospitalization
                    "Infection",
                    2009,
                    "Canada",
                    "odds ratio"))
```

##### Study 29

Maliszewski 2011 Not shown in underlying study.

##### Study 30

Mamelund 2003 Mutlivariate coefficients given with no standard error or confidence interval. Left out.

##### Study 31

Mamelund 2006

*Deprivation measure:* Individual (Census data - Using the working class vs bourgeois distinction)

*Case-criterion:* Mortality

*Control-criterion:* General population

*Period:* 1918

*Country/region:* Norway

*Type of estimate reported:* relative risk

The point-estimate is found in table 4 model 3. The confidence interval is taken from SEM's spreadsheet.

```r
meta_df <- rbind(meta_df,
                 c(31,
                   1/0.75,
                   1/1.17,
                   1/0.48,
                   NA_real_))

study_df <- rbind(study_df,
                  c(31,
                    "Individual",
                    "Mortality",
                    "General population",
                    1918,
                    "Norway",
                    "relative risk"))
```

##### Study 32

Manabe Not a relevant study

##### Study 33

Mansieaux 2015 Not a relevant study: Wrong study period - Covers a post-pandemic period (20 Dec 2010 to 20 February 2011).

##### Study 34

Mayoral 2013

*Deprivation measure:* Individual (educational level - questionnaire)

*Case-criterion:* Hospitalized

*Control-criterion:* Infected (outpatients with confirmed H1N1 infection)

*Period:* 2009

*Country/region:* Spain

*Type of estimate reported:* odds ratio

The estimates are found in table 2, adjusted OR

Multivariate coefficients with education as contrast.

```
meta_df <- rbind(meta_df,
                 c(34,
                   1/0.56,
                   1/0.87,
                   1/0.36,
                   NA_real_))

study_df <- rbind(study_df,
                  c(34,
                    "Individual",
                    "Admitted hospital",
                    "Infection",
                    2009,
                    "Spain",
                    "odds ratio"))
```

##### Study 35

Murray 2006

*Deprivation measure:* Ecological (country level income per-head)

*Case-criterion:* Mortality (estimated excess mortality in 1918-1920)

*Control-criterion:* General population

*Period:* 1918

*Country/region:* Global

*Type of estimate reported:* relative risk

The estimates are found in table 2, which compares mortality at the country level by per-head income controlling for latitude. The model estimates a gradient of -0.967 with a standard error of 0.229 where the outcome is log(pandemic mortality) and income is logged. This gives a percent to percent interpretation. As they write: "This means that a 10% increase in per-head income was associated with a 9–10% decrease in mortality."

Statistically, per head income explained almost half the variation in excess mortality seen for the 1918 pandemic (R^2 = 0.482). Unfortunately, the paper does not say what the range or standard deviation of logged per-head incomes was in the data.

In the absence of this information, I consider a contrast between two countries - one of which has double the income of the other. We call the excess mortality of these two countries Y_H and Y_L, with income X_H = 2*X_L.

What we are interested in is Y_L/Y_H. Taking the log gives us log(Y_L) - log(Y_H). We know that log(Y_H) = alpha + beta * log(X_H), while log(Y_L) = alpha + beta * log(0.5 X_H) = alpha - beta log(2) + beta log(X_H). This means that log(Y_L/Y_H) = - beta log(2). Taking the exponent gives us that Y_L/Y_H = 1.95 - or a near doubling.

Doing the same for the endpoints in the CI we get an expected difference of 1.95, with CI (1.42, 2.69)

```
meta_df <- rbind(meta_df,
                 c(35,
                   1.95,
                   1.42,
                   2.69,
                   NA_real_))

study_df <- rbind(study_df,
                  c(35,
                    "Ecological",
                    "Mortality",
                    "General population",
                    1918,
                    "Global",
                    "relative risk"))
```

#### Study 36

Navaranjan 2015

A test negative case-control study

*Deprivation measure:* Ecological

*Case-criterion:* Infection

*Control-criterion:* Other (Test negative individuals with flu-like symptoms)

*Period:* 2009

*Country/region:* Canada

*Type of estimate reported:* odds ratio

The estimates are found in table 2 - unadjusted ORs

Would have preferred to use individual education - but do not understand what their reference category is and how to get a "low education to high education" comparison. They include coefficients for "high school or less education" and for "post-secondary school completion" - is the reference group people with non-completed post-secondary school?

Using total deprivation. One score for children, one for adults.

```
meta_df <- rbind(meta_df,
                 c(36,
                   1/2.04,
                   1/4.75,
                   1/0.88,
                   NA_real_))

meta_df <- rbind(meta_df,
                 c(36,
                   1/0.91,
                   1/1.87,
                   1/0.44,
                   NA_real_))

study_df <- rbind(study_df,
                  c(36,
                    "Ecological",
                    "Infection",
                    "Other",
                    2009,
                    "Canada",
                    "odds ratio"))
```

#### Study 37

Nikolopoulos 2011

*Deprivation measure:* Ecological (GDP per capita)

*Case-criterion:* Mortality

*Control-criterion:* General population

*Period:* 2009

*Country/region:* Europe

*Type of estimate reported:* relative risk

The estimates are found in table 2 - multivariable analysis including all covariates - coefficient on GDP per capita.

GDP per capita gradient for EU countries. GDP per capita had a mean of 102 and sd of 45. The coefficient on GDP per capita was 0.017 (0.00, 0.039), and the model was a Poisson regression with log link on the mean. Looking at the reported mean, a reasonable contrast that avoids going into the extremes of the data might be between a country with 120 vs 60 on the GDP per capita scale. The relative mortality of the poor relative to rich country will then be given by exp(-60 * beta), which gives us an expected relative mortality of 0.36 (0.1, 1).

```
country_means <- c(123,115,41,98,80,117,62,110,107,116,95,63,120,131,102,49,53,268,78,130,177,56,78,42,7

hist(country_means, breaks = 10)
```

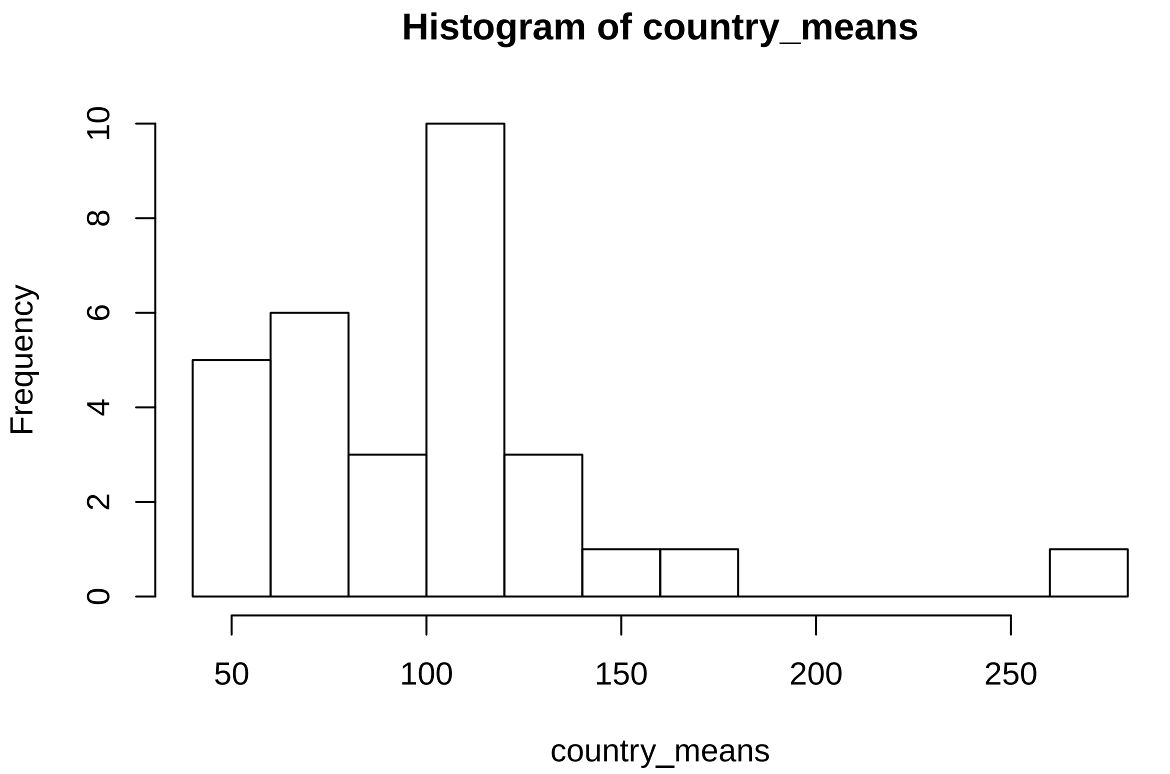


```
plot(1:30/30, sort(country_means))
abline(h = c(60, 120), lty = "dashed")
```

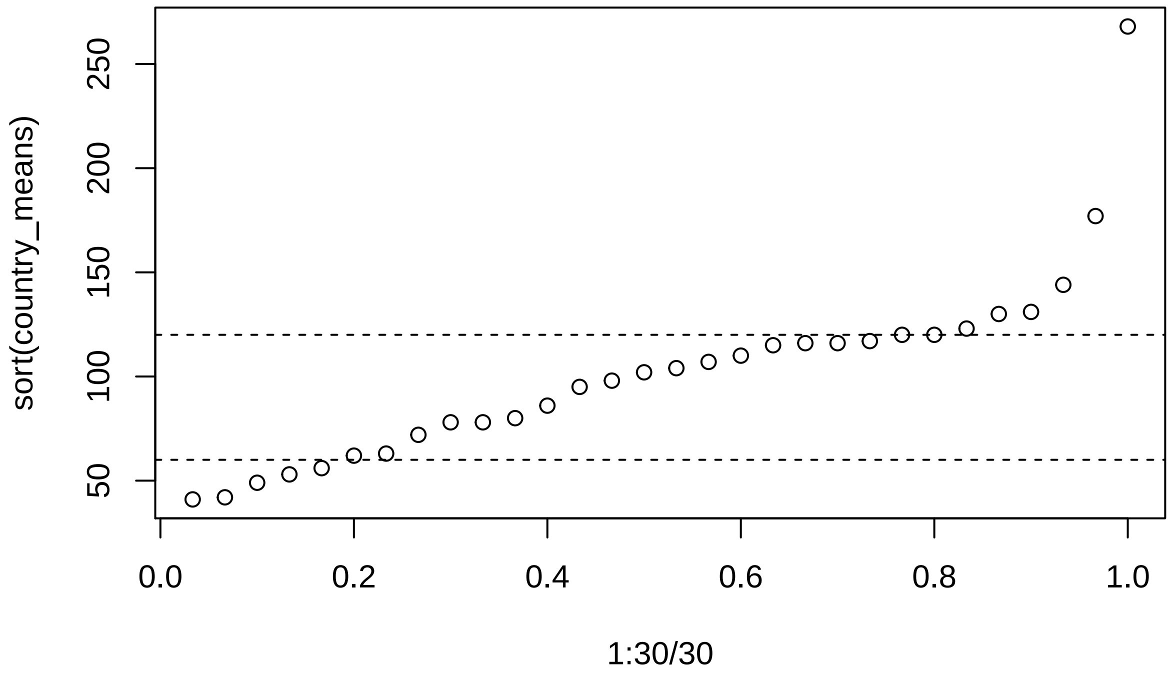

```r
meta_df <- rbind(meta_df,
                 c(37,
                   0.36,
                   0.1,
                   1,
                   NA_real_))

study_df <- rbind(study_df,
                  c(37,
                    "Ecological",
                    "Mortality",
                    "General population",
                    2009,
                    "Europe",
                    "relative risk"))
```

##### Study 38

Pasco 2012

*Deprivation measure:* Ecological (area level index of relative socioeconomic advantage and disadvantage)

*Case-criterion:* Infection

*Control-criterion:* General population (random sample from electoral rolls - 57% response)

*Period:* 2009

*Country/region:* Australia

*Type of estimate reported:* odds ratio

The estimates are found in last paragraph on page 81

```r
meta_df <- rbind(meta_df,
                 c(38,
                   2.52,
                   1.24,
                   5.13,
                   NA_real_))

study_df <- rbind(study_df,
                  c(38,
                    "Ecological",
                    "Infection",
                    "General population",
                    2009,
                    "Australia",
                    "odds ratio"))
```

##### Study 39

Pearce 2011 This study uses data on mortality during the 1918 pandemic (different waves) using an indice of deprivation based on 2000 data. I could not find estimates allowing for comparison of high to low SES groups. The closest I could find were correlations with p-values.

**Notes:**

From SEM:

> The average of Ward Scores from the Indices of Deprivation 2000: District level Presentations for England It combines a number of indicators which cover a range of domains (Income, Employment, Health Deprivation and Disability, Education, Skills and Training, Housing and Geographical Access to Services) into a single deprivation score for each area.

While they write that this indice correlated with pre-pandemic mortality, "showing that geographical predictors of social disadvantage and all-cause mortality have been stable over many decades.

The study results were initially excluded from the quantitative meta-analysis over concerns regarding the use of a control variable measured 80 years after an event. SEM correctly noted that this was not an exclusion criteria discussed in the pre-analysis plan, but examining the results we were unable to find results that allowed for a comparison of high to low SES with confidence intervals.

#### Study 40

Placzek 2014

*Deprivation measure:* Ecological (area-based socioeconomic measure at the 5 digit zip code level)

*Case-criterion:* Admitted hospital (using patient discharge data)

*Control-criterion:* General population (the estimates are adjusted for population size in case areas)

*Period:* 2009

*Country/region:* USA

*Type of estimate reported:* odds ratio

The estimates are found in Table 4 (Model d -full)

```
meta_df <- rbind(meta_df,
                 c(40,
                   0.72,
                   0.54,
                   0.96,
                   NA_real_))

study_df <- rbind(study_df,
                  c(40,
                    "Ecological",
                    "Admitted hospital",
                    "General population",
                    2009,
                    "USA",
                    "odds ratio"))
```

#### Study 41

Ponnambalam

*Deprivation measure:* Ecological (county level analysis)

*Case-criterion:* Mortality

*Control-criterion:* General population

*Period:* 2009

*Country/region:* USA

*Type of estimate reported:* relative risk

The estimates are found in table 3

This is a machine learning paper that has a set of correlations early on. They report correlations between log mortality rate and different indicators of SES: personal and household income, educational attainment, poverty rate. Although not entirely clear - it seems that the information is all at the ecological level.

We use the poverty rate (although they're all pretty similar) which is reported as 0,44 (0,35 0,52). Comparing two regions that have a standard deviation difference in poverty would thus find the poorer one having an expected 0.44 higher log(mortality), implying a relative mortality of 1.55 (1.42, 1.68).

```
meta_df <- rbind(meta_df,
                 c(41,
                   1.55,
                   1.42,
                   1.68,
                   NA_real_))

study_df <- rbind(study_df,
                  c(41,
                    "Ecological",
                    "Mortality",
                    "General population",
                    2009,
                    "USA",
                    "relative risk"))
```

#### Study 42

Pujol 2016

Two separate analyses:

##### Estimate 1:

*Deprivation measure:* Individual (occupation social class)

*Case-criterion:* Infection

*Control-criterion:* Other ("Ambulatory controls were people attending a primary-care centre for any reason other than influenza-like illness.")

*Period:* 2009

*Country/region:* Spain

*Type of estimate reported:* odds ratio

The estimates are found in abstract

```
meta_df <- rbind(meta_df,
                 c(42,
                   0.97,
```

```
                   0.74,
                   1.27,
                   NA_real_))

study_df <- rbind(study_df,
                  c(42,
                    "Individual",
                    "Infection",
                    "Other",
                    2009,
                    "Spain",
                    "odds ratio"))
```

**Estimate 2:**

*Deprivation measure:* Individual (occupation social class)

*Case-criterion:* Hospitalization

*Control-criterion:* Infection (non-hospitalized - attending primary-care centre)

*Period:* 2009

*Country/region:* Spain

*Type of estimate reported:* odds ratio

The estimates are found in abstract

```
meta_df <- rbind(meta_df,
                 c(42.5,
                   1.53,
                   1.01,
                   2.31,
                   NA_real_))


study_df <- rbind(study_df,
                  c(42.5,
                    "Individual",
                    "Admitted hospital",
                    "Infection",
                    2009,
                    "Spain",
                    "odds ratio"))
```

#### Study 43

Quinn 2011 Not included.

#### Study 44

Rutter 2012

*Deprivation measure:* Ecological (area based socioeconomic deprivation)

*Case-criterion:* Mortality

*Control-criterion:* General population

*Period:* 2009

*Country/region:* UK

*Type of estimate reported:* relative risk

The estimates are found in the abstract.

Least to highest deprived.

```
meta_df <- rbind(meta_df,
                 c(44,
                   3.1,
                   2.2,
                   4.4,
                   NA_real_))

study_df <- rbind(study_df,
                  c(44,
                    "Ecological",
                    "Mortality",
                    "General population",
                    2009,
                    "UK",
                    "relative risk"))
```

#### Study 45

Simonsen 2014 Not included.

#### Study 46

Sloan 2017 Abstract - paper included elsewhere

#### Study 47

Suarthana 2010 Not relevant

#### Study 48

Summers 2013

A study of soldiers in first world war - mortality by (pre-enlistment) occupational class

*Deprivation measure:* individual - pre-enlistment occupation class

*Case-criterion:* Mortality

*Control-criterion:* General population (more specifically: at risk population, Military Personnel)

*Period:* 1918

*Country/region:* New Zealand

*Type of estimate reported:* relative risk

The estimates are found in Table 1

```
meta_df <- rbind(meta_df,
                 c(48,
                   0.9,
                   0.6,
                   1.2,
                   NA_real_))
study_df <- rbind(study_df,
                  c(48,
                    "Individual",
                    "Mortality",
                    "General population",
                    1918,
                    "New Zealand",
                    "relative risk"))
```

#### Study 49

Summers 2010

*Deprivation measure:* Individual (pre-enlistment occupation group)

*Case-criterion:* Mortality

*Control-criterion:* General population (that is: at risk population, military personnel on same boat)

*Period:* 1918

*Country/region:* New Zealand

*Type of estimate reported:* odds ratio

The estimates are found in table 2 - model 2

```
meta_df <- rbind(meta_df,
                 c(49,
                   0.83,
                   0.5,
                   1.38,
                   NA_real_))
study_df <- rbind(study_df,
                  c(49,
                    "Individual",
                    "Mortality",
                    "General population",
                    1918,
                    "New Zealand",
                    "odds ratio"))
```

#### Study 50

Sydenstricker 1931

###### Estimate 1: Morbidity

*Deprivation measure:* Individual ("economic condition" of household as judged at first impression by enumerator with no pre-specified criteria)

*Case-criterion:* Infection (self reported influenza, pneumonia or indefinitely diagnosed illness suspected to be influenza)

*Control-criterion:* General population

*Period:* 1918

*Country/region:* US

*Type of estimate reported:* odds ratio

The numbers used are found in table 1 - incidence by general economic condition - all ages, all localities.

```
cases <- c(1486, 2211)
controls <- c(3988, 9550) - cases

or <- (cases[1]/controls[1])/(cases[2]/controls[2])
cat("Odds ratio: ", or, "\n")
```

```
## Odds ratio:  1.971422
```

```
log_or_se <- sqrt(sum(1/c(cases, controls)))
cat("Log_or_se: ", log_or_se)
```

```
## Log_or_se:  0.04075746
```

```
meta_df <- rbind(meta_df,
                 c(50,
                   or,
                   exp(log(or) - 1.96 * log_or_se),
                   exp(log(or) + 1.96 * log_or_se),
                   log_or_se))
study_df <- rbind(study_df,
                  c(50,
                    "Individual",
                    "Infection",
                    "General population",
                    1918,
                    "USA",
                    "odds ratio"))
```

###### Estimate 2: Mortality relative to infection

*Deprivation measure:* Individual ("economid condition" of household as judged at first impression by enumerator with no pre-specified criteria)

*Case-criterion:* Death (household stated influenza, pneumonia or indefinitely diagnosed illness suspected to be influenza)

*Control-criterion:* General population

*Period:* 1918

*Country/region:* US

*Type of estimate reported:* odds ratio

These numbers are more shaky, as the raw numbers are not included in the paper. For mortality, the mortality rate by SES is given as rates after correcting for age. Age-correction seems to have been as follows: A separate (not included) estimate of mortality rate at the SES-age_group level, averaged using age-group weights similar to overall continental US in 1910.

The numbers used to calculate odds ratio and its standard error were derived as follows:

Using table IV, we have case fatality rates of 2.8 per 100 for the very poor and 1.5 for the well off (both adjusted for age-composition)

Using table II, we have infection rates of 364 per 1000 very poor and 252 per well to do.

*Ignoring* the age adjustment, we have 9550 individuals in the study from the well to-do and 3988 from the very poor groups (as given in table 1).

This gives us approximately:

infected poor: 0.364 * 3988 = 1451.6 infected well-off: 0.252 * 9550 = 2406.6

dead poor: 0.028 * 1451.6 = 40.6 dead well-off: 0.015 * 2406.6 = 36.1

This point estimate is close but not identical to what you get if you divide the case fatality rate of the very poor (2.8/1.5 = 1.87).

```
cases <- c(40.6, 36.1)
controls <- c(1451.6, 2406.6) - cases

or <- (cases[1]/controls[1])/(cases[2]/controls[2])
cat("Odds ratio: ", or, "\n")
```

```
## Odds ratio:  1.889434
```

```
log_or_se <- sqrt(sum(1/c(cases, controls)))
cat("Log_or_se: ", log_or_se)
```

```
## Log_or_se:  0.2312184
```

```
meta_df <- rbind(meta_df,
                 c(50.5,
                   or,
                   exp(log(or) - 1.96 * log_or_se),
                   exp(log(or) + 1.96 * log_or_se),
                   log_or_se))
study_df <- rbind(study_df,
                  c(50.5,
                    "Individual",
                    "Mortality",
                    "Infection",
                    1918,
                    "USA",
                    "odds ratio"))
```

#### Study 51

Tam 2014

Studies regular influenza - not pandemic

*Deprivation measure:* Ecological (area based using info from American Community Survey - median income used as SES indicator)

*Case-criterion:* Admitted hospital - laboratory confirmed influenza in the 2007-2008 throughtout 2010-2011 influenza season

*Control-criterion:* General population

*Period:* 2009

*Country/region:* US

*Type of estimate reported:* odds ratio

The estimates are based on approximated numbers calculated from Supplemental table S1 (data for 2009-2010). Note that there is a mistake in the table: the numbers for areas categorized by income in table S1 don't match the aggregate numbers in the article table 2 and the association is reversed. In table 2 of the article the authors report a total of 218 low-income cases and 57 high income cases (the numbers I initially had used). In the supporting table, the sum of the high income cases across all years is 218 while the sum of all the low income cases is 57.

In the text they write: "Influenza-related hospitalization of adults associated with low census tract socioeconomic status and female sex in New Haven County, Connecticut, 2007-2011

The incidence increased as the percent of persons living below poverty in a census tract increased, as the percent of persons in a census tract with no high school diploma increased, as the percent of crowded households in a census tract increased, as the percent of non-English speaking households in a census tract increased, and as median income in the census tract decreased. These trends were present in each influenza season including the 2009-10 H1N1pdm season (Table S1)."

Given this, we have two pieces of information indicating that flu was over-represented in low SES and under-represented in high SES (table 2 and the text), and one piece indicating the opposite. We take the labels in table S1 to be wrong and adjust for this. This gives us

Low income cases: 43 High income cases: 13

Approximate size of low_income population size in person-years, using *age adjusted* incidence rates per 100 000 person-years: 13 / (72.1/100000) = 18 030

Approximate size of high_income population size in person-years, using *age adjusted* incidence rates per 100 000 person-years: 43 / (20.2/100000) = 212 870

```
cases <- c(13, 43)
controls <- c(18030, 212870) - cases

or <- (cases[1]/controls[1])/(cases[2]/controls[2])
cat("Odds ratio: ", or, "\n")
```

```
## Odds ratio:  3.571241
```

```
log_or_se <- sqrt(sum(1/c(cases, controls)))
cat("Log_or_se: ", log_or_se)
```

```
## Log_or_se:  0.3166056
```

```
meta_df <- rbind(meta_df,
                 c(51,
                   or,
                   exp(log(or) - 1.96 * log_or_se),
                   exp(log(or) + 1.96 * log_or_se),
                   log_or_se))
study_df <- rbind(study_df,
                  c(51,
                    "Ecological",
```

```
                     "Admitted hospital",
                     "General population",
                     2009,
                     "USA",
                     "odds ratio"))
```

##### Study 52

Thompson 2011

###### Estimate 1 - hospitalization

*Deprivation measure:* Ecological (County median household income)

*Case-criterion:* Admitted hospital

*Control-criterion:* General population

*Period:* 2009

*Country/region:* US

*Type of estimate reported:* relative risk

The estimates are found in Table 2 (adjusted RR).

```
meta_df <- rbind(meta_df,
                 c(52,
                   1.6,
                   1.2,
                   2.1,
                   NA_real_))

study_df <- rbind(study_df,
                  c(52,
                    "Ecological",
                    "Admitted hospital",
                    "General population",
                    2009,
                    "USA",
                    "relative risk"))
```

###### Estimate 2 - Mechanical ventilation

*Deprivation measure:* Ecological (County median household income)

*Case-criterion:* Severe hospital

*Control-criterion:* Admitted hospital

*Period:* 2009

*Country/region:* US

*Type of estimate reported:* odds ratio

The estimates are found in Table 4 (adjusted RR). I use model 2 estimates (unlike the model 1 estimates extracted to the Excel sheet), as these are based on more cases (they include individuals with missing obesity info, which would otherwise cause substantial attrition).

```
meta_df <- rbind(meta_df,
                 c(52.5,
                   1.0,
                   0.2,
                   4.6,
                   NA_real_))

study_df <- rbind(study_df,
                  c(52.5,
                    "Ecological",
                    "Severe hospital",
                    "Admitted hospital",
                    2009,
                    "USA",
                    "odds ratio"))
```

###### Estimate 3 - Death

*Deprivation measure:* Ecological (County median household income)

*Case-criterion:* Death

*Control-criterion:* Admitted hospital

*Period:* 2009

*Country/region:* US

*Type of estimate reported:* odds ratio

The estimates are found in Table 3 - adjusted estimates for income groups are not available in table 4.

```
meta_df <- rbind(meta_df,
                 c(52.75,
                   0.9,
                   0.3,
                   2.4,
                   NA_real_))

study_df <- rbind(study_df,
                  c(52.75,
                    "Ecological",
                    "Mortality",
                    "Admitted hospital",
                    2009,
                    "USA",
                    "odds ratio"))
```

#### Study 53

Thompson 2012

*Deprivation measure:* Individual (annual household income)

*Case-criterion:* Infected (serological test - for some of the sample due to vaccination)

*Control-criterion:* Other - Adults presenting to community clinic - convenience sample

*Period:* 2009

*Country/region:* Canada

*Type of estimate reported:* odds ratio

The estimates are based on the following numbers for two groups distinguished by period (the third group is from the period after vaccination has begun rolling out and is excluded here):

The counts given for different income groups in table 1 and 2 appear to differ. I assume the correct ones are in table 2 where the positivity rates are given.

##### Estimate 1 - Group A

```
inc_groups_a <- c(81, 32)
pos_rate_a <- c(0.062, 0.031)

cases <- inc_groups_a * pos_rate_a
controls <- inc_groups_a * (1 - pos_rate_a)

or <- (cases[1]/controls[1])/(cases[2]/controls[2])
cat("Odds ratio: ", or, "\n")
```

```
## Odds ratio:  2.066098
```

```
log_or_se <- sqrt(sum(1/c(cases, controls)))
cat("Log_or_se: ", log_or_se)
```

```
## Log_or_se:  1.119196
```

```
meta_df <- rbind(meta_df,
                 c(53,
                   or,
                   exp(log(or) - 1.96 * log_or_se),
                   exp(log(or) + 1.96 * log_or_se),
                   log_or_se))

study_df <- rbind(study_df,
                  c(53,
                    "Individual",
                    "Infection",
                    "Other",
                    2009,
                    "Australia",
                    "odds ratio"))
```

##### Estimate 2 - Group B

Because there are zero observations of infections in the high income group I add 1 to all cells and scale them down to their previous sum.

```
inc_groups_b <- c(113, 15)
pos_rate_b <- c(0.186, 0.0)
```

```r
cases <- inc_groups_b * pos_rate_b
controls <- inc_groups_b * (1 - pos_rate_b)

temp_norm_factor <- sum(cases, controls)/(sum(cases, controls) + 4)

cases <- (cases + 1) * temp_norm_factor
controls <- (controls + 1) * temp_norm_factor


or <- (cases[1]/controls[1])/(cases[2]/controls[2])
cat("Odds ratio: ", or, "\n")
```

```
## Odds ratio:  3.788776
```

```r
log_or_se <- sqrt(sum(1/c(cases, controls)))
cat("Log_or_se: ", log_or_se)
```

```
## Log_or_se:  1.074072
```

```r
meta_df <- rbind(meta_df,
                 c(53.5,
                   or,
                   exp(log(or) - 1.96 * log_or_se),
                   exp(log(or) + 1.96 * log_or_se),
                   log_or_se))

study_df <- rbind(study_df,
                  c(53.5,
                    "Individual",
                    "Infection",
                    "Other",
                    2009,
                    "Australia",
                    "odds ratio"))
```

##### Study 54

Tora-Rocamora 2012 Not relevant.

##### Study 55

Trauer 2011

*Deprivation measure:* Ecological (socio-economic indexes for area)

*Case-criterion:* Infection

*Control-criterion:* Other (outpatient serum specimens)

*Period:* 2009

*Country/region:* Astralia

*Type of estimate reported:* odds ratio

The estimates are found in table 3.

```
meta_df <- rbind(meta_df,
                 c(55,
                   1.21,
                   0.7,
                   2.12,
                   NA_real_))
study_df <- rbind(study_df,
                  c(55,
                    "Ecological",
                    "Infection",
                    "Other",
                    2009,
                    "Australia",
                    "odds ratio"))
```

##### Study 56

Viboud 2012 Not relevant.

##### Study 57

Zarychanski 2010

###### Estimate 1 - ICU vs community cases

*Deprivation measure:* ecological - area (income quintile using postal codes)

*Case-criterion:* Severe hospital (ICU treatment)

*Control-criterion:* Infection (laboratory confirmed cases)

*Period:* 2009

*Country/region:* Canada

*Type of estimate reported:* odds ratio

The estimates are found in table 2

```
meta_df <- rbind(meta_df,
                 c(57,
                   1.06,
                   0.39,
                   2.88,
                   NA_real_))

study_df <- rbind(study_df,
                  c(57,
                    "Ecological",
                    "Severe hospital",
                    "Infection",
                    2009,
                    "Canada",
                    "odds ratio"))
```

##### Estimate 2 - ICU vs hospitalized

*Deprivation measure:* ecological - area (income quintile using postal codes)

*Case-criterion:* Severe hospital (ICU treatment)

*Control-criterion:* Admitted hospital

*Period:* 2009

*Country/region:* Canada

*Type of estimate reported:* odds ratio

The estimates are found in table 2

```
meta_df <- rbind(meta_df,
                c(57.5,
                  0.68,
                  0.33,
                  1.42,
                  NA_real_))

study_df <- rbind(study_df,
                 c(57.5,
                   "Ecological",
                   "Severe hospital",
                   "Admitted hospital",
                   2009,
                   "Canada",
                   "odds ratio"))
```

#### Study 58

Zhang 2013

*Deprivation measure:* Individual (interviews)

*Case-criterion:* Infected (households with self-quarantined index patient and a secondary case)

*Control-criterion:* General population (matched households with self-quarantined index patient and a close contact)

*Period:* 2009

*Country/region:* China

*Type of estimate reported:* odds ratio

The estimates are found in table 2. This is one of those cases where one could discuss whether some of the controlled for variables are "on the causal path" from poverty to infection (e.g., controlling for "sharing room with index case patient"). However, a simple case/control counts ratio of lowest to highest income (based on counts in table 1) gives almost identical point estimate: $(11/9)/(8/16) = 2.44$, while the point estimate from the multivariate on education is $1/0.42 = 2.38$

```
meta_df <- rbind(meta_df,
                c(58,
                  1/0.42,
                  1/0.83,
                  1/0.22,
```

```
                   NA_real_))

study_df <- rbind(study_df,
                  c(58,
                    "Individual",
                    "Infection",
                    "General population",
                    2009,
                    "China",
                    "odds ratio"))
```

##### Study 59

Zhao 2015

*Deprivation measure:* Ecological (area index)

*Case-criterion:* Mortality

*Control-criterion:* General poulation

*Period:* 2009

*Country/region:* UK

*Type of estimate reported:* relative risk

The estimates are found in table 3.

Deprivation index quantiles - here a higher deprivation index is more deprived.

Only one period fits study selection criteria.

```
meta_df <- rbind(meta_df,
                 c(59,
                   2.08,
                   1.49,
                   2.91,
                   NA_real_))

study_df <- rbind(study_df,
                  c(59,
                    "Ecological",
                    "Mortality",
                    "General population",
                    2009,
                    "UK",
                    "relative risk"))
```

### Preparing analysis data

#### Adding study level information from spreadsheet

We add in author, journal and year of each study using a csv exported from the data extraction excel sheet.

```
study_info <- fread("study_information.csv")

setnames(study_info, "Study _index", "study_index_orig")

meta_dt <- data.table(meta_df)
study_dt <- data.table(study_df)

study_dt[, study_index := as.numeric(study_index)]

meta_dt <- merge(meta_dt,
                 study_dt,
                 by = "study_index",
                 all.x = T)

meta_dt[, study_index_orig := as.integer(floor(study_index))]

meta_dt <- merge(meta_dt,
                 study_info,
                 by = "study_index_orig",
                 all.x = T)
```

##### Calculating necessary magnitudes

```
meta_dt[, ':='(log_or = log(or))]
meta_dt[, log_se_orig := log_se]
meta_dt[is.na(log_se) == T,
        log_se := (log(ub) - log(lb))/(1.96 * 2)]
meta_dt[is.na(lb) == T,
        lb := exp(log_or - 1.96 * log_se)]
meta_dt[is.na(ub) == T,
        ub := exp(log_or + 1.96 * log_se)]
```

#### Analysis

There are 35 different studies contributing a total of 46 estimates.

##### Standard meta-analysis

###### All studies

```
# Fix year of Sydenstricker

meta_dt[Author == "Sydenstricker", Year := 1931]

random_effects <- rma(yi = log_or,
                      sei = log_se,
                      data = meta_dt)
```

```r
forest(random_effects,
       atransf = exp,
       order = "prec",
       showweights = T,
       slab = meta_dt[, list(v = paste0(Author, ", ", Year))]$v)
```

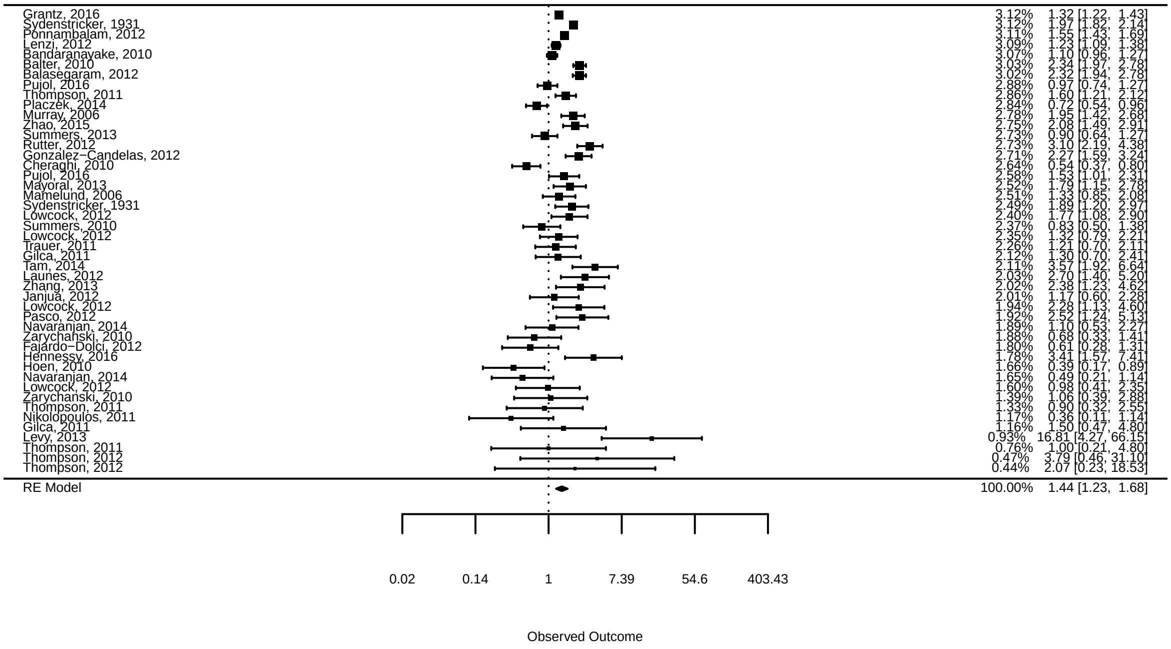


```r
random_effects
```

```
##
## Random-Effects Model (k = 46; tau^2 estimator: REML)
##
## tau^2 (estimated amount of total heterogeneity): 0.2053 (SE = 0.0595)
## tau (square root of estimated tau^2 value):      0.4531
## I^2 (total heterogeneity / total variability):   92.48%
## H^2 (total variability / sampling variability):  13.29
##
## Test for Heterogeneity:
## Q(df = 45) = 308.5241, p-val < .0001
##
## Model Results:
##
## estimate      se    zval    pval   ci.lb   ci.ub
##   0.3631  0.0803  4.5203  <.0001  0.2057  0.5205  ***
##
## ---
## Signif. codes:  0 '***' 0.001 '**' 0.01 '*' 0.05 '.' 0.1 ' ' 1
```

##### Subsample analyses by comparison type

```r
meta_dt[, control_fact := factor(control_crit, levels = c("General population",
                                                          "Infection",
                                                          "Admitted hospital",
```

```
                                                                 "Other"),
                                  ordered = T)]
meta_dt[, case_fact := factor(case_crit, levels = c("Infection",
                                                    "Admitted hospital",
                                                    "Severe hospital",
                                                    "Mortality"),
                                  ordered = T)]
meta_dt[, comparison_type := .GRP, keyby = list(control_fact, case_fact)]
meta_dt[, comparison_label := paste("Control: ", control_crit, "\n", "Case: ", case_crit)]

temp_list <- list()

for (i in 1:max(meta_dt$comparison_type)){
  temp <- rma(yi = log_or,
                        sei = log_se,
                        data = meta_dt[comparison_type == i])

  temp_list[[i]] <- data.table(distinction = "Comparison type",
                               type = meta_dt[comparison_type == i]$comparison_label[1],
                               order = max(meta_dt$comparison_type) + 1 -i,
                               pooled_effect = temp$b[[1]],
                               lb = temp$ci.lb,
                               ub = temp$ci.ub,
                               tau = sqrt(temp$tau2),
                               study_count = temp$k.all)

}

comp_results <- rbindlist(temp_list)

setkey(comp_results, order)
comp_results[, type_fact := factor(type, levels = unique(comp_results$type), ordered = T)]

ggplot(comp_results,
       aes(type_fact, exp(pooled_effect))) +
  geom_point(aes(size = study_count), col = "grey") +
  geom_point(aes(size = 0.5)) +
  geom_errorbar(aes(ymin = exp(lb),
                    ymax = exp(ub))) +
  facet_wrap(~ distinction,
             scales = "free_x") +
  geom_hline(yintercept = 1, linetype = "dashed") +
  coord_flip()+
  labs(x = "", y = "Odds ratio") +
  guides(size = guide_legend(override.aes = list(col = "grey")))
```

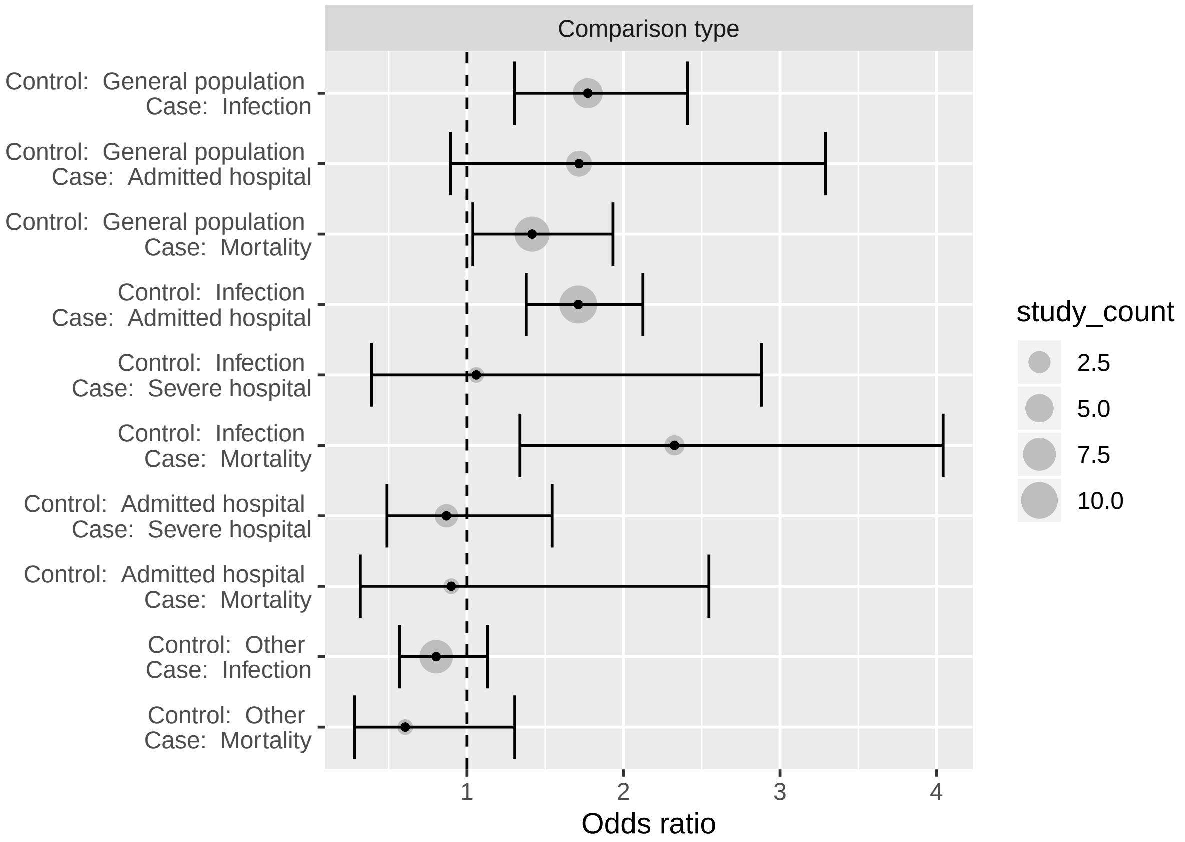


```r
knitr::kable(comp_results[, list(type,
                                  study_count,
                                  pooled_effect = exp(pooled_effect),
                                  lb = exp(lb),
                                  ub = exp(ub),
                                  tau)], digits = 2)
```

| type | study_count | pooled_effect | lb | ub | tau |
|---|---|---|---|---|---|
| Control: Other Case: Mortality 1 | 0.61 | 0.28 1.31 0. | 00 | | |
| Control: Other Case: Infection 8 | 0.80 | 0.57 1.13 0. | 33 | | |
| Control: Admitted hospital Case: Mortality 1 0.90 | 0.32 2.55 | 0.00 | | | |
| Control: Admitted hospital Case: Severe hospital 3 0.87 | 0.49 1.54 | 0.00 | | | |
| Control: Infection Case: Mortality 2 | 2.33 1.34 | 4.04 0.26 | | | |
| Control: Infection Case: Severe hospital 1 | 1.06 0.39 | 2.88 0.00 | | | |
| Control: Infection Case: Admitted hospital 11 | 1.71 1.38 | 2.12 0.25 | | | |
| Control: General population Case: Mortality 9 1.42 | 1.04 1.93 | 0.43 | | | |
| Control: General population Case: Admitted hospital 4 1.72 | 0.89 3.29 | 0.64 | | | |
| Control: General population | | | | | |

| type | study_count | pooled_effect | lb | ub | tau |
|---|---|---|---|---|---|
| Case: Infection 6 1.77 | 1.30 2.41 | 0.32 | | | |

###### Subsample analyses by individual distinctions

```
temp_list <- list()

# Individual vs ecological measures

temp <- rma(yi = log_or,
                      sei = log_se,
                      data = meta_dt[deprivation_type == "Ecological"])

temp_list[[1]] <- data.table(distinction = "Measure",
                             type = "Ecological",
                             pooled_effect = temp$b[[1]],
                             lb = temp$ci.lb,
                             ub = temp$ci.ub,
                             tau = sqrt(temp$tau2),
                             study_count = temp$k.all)

temp <- rma(yi = log_or,
                      sei = log_se,
                      data = meta_dt[deprivation_type == "Individual"])

temp_list[[2]] <- data.table(distinction = "Measure",
                             type = "Individual",
                             pooled_effect = temp$b[[1]],
                             lb = temp$ci.lb,
                             ub = temp$ci.ub,
                             tau = sqrt(temp$tau2),
                             study_count = temp$k.all)
# Type of case outcome
temp <- rma(yi = log_or,
                      sei = log_se,
                      data = meta_dt[case_crit == "Infection"])


temp_list[[3]] <- data.table(distinction = "Case type",
                             type = "Infection",
                             pooled_effect = temp$b[[1]],
                             lb = temp$ci.lb,
                             ub = temp$ci.ub,
                             tau = sqrt(temp$tau2),
                             study_count = temp$k.all)
temp <- rma(yi = log_or,
                      sei = log_se,
                      data = meta_dt[case_crit == "Admitted hospital"])


temp_list[[4]] <- data.table(distinction = "Case type",
                             type = "Admitted hospital",
```

```r
                              pooled_effect = temp$b[[1]],
                              lb = temp$ci.lb,
                              ub = temp$ci.ub,
                              tau = sqrt(temp$tau2),
                              study_count = temp$k.all)
temp <- rma(yi = log_or,
                       sei = log_se,
                       data = meta_dt[case_crit == "Severe hospital"])


temp_list[[5]] <- data.table(distinction = "Case type",
                              type = "Severe hospital",
                              pooled_effect = temp$b[[1]],
                              lb = temp$ci.lb,
                              ub = temp$ci.ub,
                              tau = sqrt(temp$tau2),
                              study_count = temp$k.all)
temp <- rma(yi = log_or,
                       sei = log_se,
                       data = meta_dt[case_crit == "Mortality"])


temp_list[[6]] <- data.table(distinction = "Case type",
                              type = "Mortality",
                              pooled_effect = temp$b[[1]],
                              lb = temp$ci.lb,
                              ub = temp$ci.ub,
                              tau = sqrt(temp$tau2),
                              study_count = temp$k.all)
# Type of Control outcome

temp <- rma(yi = log_or,
                       sei = log_se,
                       data = meta_dt[control_crit == "Other"])


temp_list[[7]] <- data.table(distinction = "Control type",
                              type = "Other",
                              pooled_effect = temp$b[[1]],
                              lb = temp$ci.lb,
                              ub = temp$ci.ub,
                              tau = sqrt(temp$tau2),
                              study_count = temp$k.all)

temp <- rma(yi = log_or,
                       sei = log_se,
                       data = meta_dt[control_crit == "General population"])


temp_list[[8]] <- data.table(distinction = "Control type",
                              type = "General population",
                              pooled_effect = temp$b[[1]],
                              lb = temp$ci.lb,
```

```
                              ub = temp$ci.ub,
                              tau = sqrt(temp$tau2),
                              study_count = temp$k.all)

temp <- rma(yi = log_or,
                      sei = log_se,
                      data = meta_dt[control_crit == "Infection"])


temp_list[[9]] <- data.table(distinction = "Control type",
                              type = "Infection",
                              pooled_effect = temp$b[[1]],
                              lb = temp$ci.lb,
                              ub = temp$ci.ub,
                              tau = sqrt(temp$tau2),
                              study_count = temp$k.all)

temp <- rma(yi = log_or,
                      sei = log_se,
                      data = meta_dt[control_crit == "Admitted hospital"])


temp_list[[10]] <- data.table(distinction = "Control type",
                              type = "Admitted hospital",
                              pooled_effect = temp$b[[1]],
                              lb = temp$ci.lb,
                              ub = temp$ci.ub,
                              tau = sqrt(temp$tau2),
                              study_count = temp$k.all)

# Period
temp <- rma(yi = log_or,
                      sei = log_se,
                      data = meta_dt[period == "1918"])


temp_list[[11]] <- data.table(distinction = "Period",
                              type = "1918",
                              pooled_effect = temp$b[[1]],
                              lb = temp$ci.lb,
                              ub = temp$ci.ub,
                              tau = sqrt(temp$tau2),
                              study_count = temp$k.all)


temp <- rma(yi = log_or,
                      sei = log_se,
                      data = meta_dt[period == "2009"])

temp_list[[12]] <- data.table(distinction = "Period",
                              type = "2009",
                              pooled_effect = temp$b[[1]],
                              lb = temp$ci.lb,
```

```
                             ub = temp$ci.ub,
                             tau = sqrt(temp$tau2),
                             study_count = temp$k.all)


# Method
temp <- rma(yi = log_or,
                      sei = log_se,
                      data = meta_dt[method == "relative risk"])


temp_list[[13]] <- data.table(distinction = "Method",
                             type = "relative risk",
                             pooled_effect = temp$b[[1]],
                             lb = temp$ci.lb,
                             ub = temp$ci.ub,
                             tau = sqrt(temp$tau2),
                             study_count = temp$k.all)



temp <- rma(yi = log_or,
                      sei = log_se,
                      data = meta_dt[method == "odds ratio"])


temp_list[[14]] <- data.table(distinction = "Method",
                             type = "odds ratio",
                             pooled_effect = temp$b[[1]],
                             lb = temp$ci.lb,
                             ub = temp$ci.ub,
                             tau = sqrt(temp$tau2),
                             study_count = temp$k.all)


subsample_results <- rbindlist(temp_list)
subsample_results[, ':='(type_fact = factor(type, levels = c("1918",
                                                              "2009",
                                                              "Ecological",
                                                              "Individual",
                                                              "General population",
                                                              "Infection",
                                                              "Admitted hospital",
                                                              "Severe hospital",
                                                              "Mortality",
                                                              "relative risk",
                                                              "odds ratio",
                                                              "Other"),
                                               ordered = T))]
```

```
subsample_results[!(distinction %in% c("Case type", "Control type"))]
```

```
##    distinction          type pooled_effect          lb        ub       tau
## 1:     Measure    Ecological     0.3196963 0.05051038 0.5888823 0.5330767
```

```
## 2:     Measure    Individual     0.3735033 0.17939487 0.5676117 0.4056332
## 3:      Period          1918     0.3484311 0.09173299 0.6051292 0.3024308
## 4:      Period          2009     0.3680093 0.17924437 0.5567743 0.5008664
## 5:      Method relative risk     0.4759048 0.23043583 0.7213737 0.3537957
## 6:      Method    odds ratio     0.3288070 0.13516827 0.5224458 0.4858710
##    study_count     type_fact
## 1:          20    Ecological
## 2:          26    Individual
## 3:           7          1918
## 4:          39          2009
## 5:          10 relative risk
## 6:          36    odds ratio
```

```r
ggplot(subsample_results[!(distinction %in% c("Case type", "Control type"))],
       aes(type_fact, exp(pooled_effect))) +
  geom_point(aes(size = study_count),
             col = "gray") +
  geom_point(aes(size = 0.5)) +
  geom_errorbar(aes(ymin = exp(lb),
                    ymax = exp(ub))) +
  facet_wrap(~ distinction,
             scales = "free") +
  geom_hline(yintercept = 1, linetype = "dashed") +
  theme(axis.text.x = element_text(angle = 90, vjust = 0.5, hjust=1)) +
  labs(x = "", y = "Odds ratio") +
  guides(size = guide_legend(override.aes = list(col = "grey")))
```

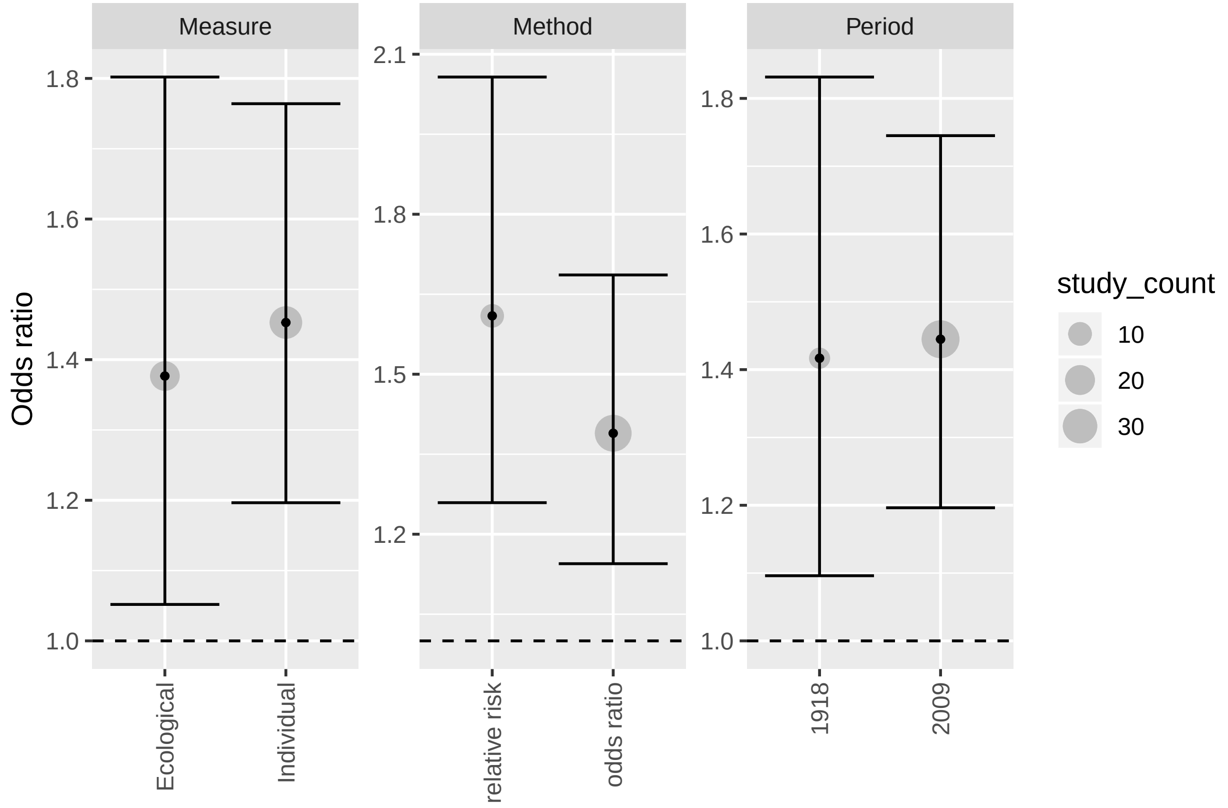


```r
knitr::kable(subsample_results[!(distinction %in% c("Case type", "Control type")),
                               list(distinction,
```

```
                                    type,
                                    study_count,
                                    pooled_effect = exp(pooled_effect),
                                    lb = exp(lb),
                                    ub = exp(ub),
                                    tau)], digits = 2)
```

| distinction | type | study_count | pooled_effect | lb | ub | tau |
|---|---|---|---|---|---|---|
| Measure | Ecological | 20 | 1.38 | 1.05 | 1.80 | 0.53 |
| Measure | Individual | 26 | 1.45 | 1.20 | 1.76 | 0.41 |
| Period | 1918 | 7 | 1.42 | 1.10 | 1.83 | 0.30 |
| Period | 2009 | 39 | 1.44 | 1.20 | 1.75 | 0.50 |
| Method | relative risk | 10 | 1.61 | 1.26 | 2.06 | 0.35 |
| Method | odds ratio | 36 | 1.39 | 1.14 | 1.69 | 0.49 |

#### Bayesian - for comparison

##### Data preparation

We prepare the data for a simple Bayesian hierarchical model.

```
use_data <- meta_dt[, list(study_index, or, lb, ub, log_se)]
apply(use_data, 2, function(x) mean(is.na(x)))
```

```
## study_index          or          lb          ub      log_se
##           0           0           0           0           0
```

```
stan_data_base = list(N = nrow(use_data),
                      y = log(use_data$or),
                      se = use_data$log_se)
```

##### Model code and prior choices

The model has two substantive priors - for the mean and standard deviation of the effect distribution.

Both are assigned a normal prior with mean zero and standard deviation of 0.4. For the effect mean, this expresses a belief that the true average effect across studies will most likely be in the range of exp(-0.8, 0.8) = 0.45 to 2.25. Put differently, we expect that low socioeconomic status (relative to high) is unlikely to predict a change in flu outcome risks by more than a factor of two on average. The prior for the standard deviation similarly expresses a belief that the effect estimated by a single study is unlikely to differ from the average effect by more than a factor of 2.

The estimation code:

```
data {
  int<lower=0> N;
  vector[N] y;
  vector[N] se;
}
parameters {
  real mu;
  real<lower = 0> sigma;
```

```
  vector[N] study_std;
}
transformed parameters {
  vector[N] study_re;
  vector[N] study_estimate;

  study_re = mu + sigma * study_std;
  study_estimate = exp(study_re);
}
model {
  mu ~ normal(0, 0.4);
  sigma ~ normal(0, 0.4);
  study_std ~ std_normal();

  y ~ normal(study_re, se);
}
generated quantities {
  real mu_exp = exp(mu);
}
```

```
base_bayes <- stan("random effect.stan",
                   data = stan_data_base,
                   iter = 10000,
                   refresh = 10000)
```

The model performs well, with no signs of divergence or other estimation issues.

##### Results

```
round(summary(base_bayes, pars = c("mu_exp", "sigma"))$summary, 2)
```

```
##        mean se_mean   sd 2.5%  25%  50% 75% 97.5%   n_eff Rhat
## mu_exp 1.42       0 0.11 1.21 1.35 1.42 1.5   1.66 3925.96    1
## sigma  0.46       0 0.07 0.33 0.41 0.45 0.5   0.61 4739.25    1
```

This is essentially the same result as the results using the standard random effects model.

#### Bayesian - with study level features

##### Data preparation

```
use_data <- meta_dt[, list(study_index, or, lb, ub, log_se, deprivation_type, case_crit, control_crit, 
apply(use_data, 2, function(x) mean(is.na(x)))
```

```
##      study_index               or               lb               ub
##                0                0                0                0
##           log_se deprivation_type        case_crit     control_crit
##                0                0                0                0
##           period          country           method  comparison_type
##                0                0                0                0
```

```r
use_data[, ses_index := .GRP, keyby = deprivation_type]
use_data[, case_type_index := .GRP, keyby = case_crit]
use_data[, control_type_index := .GRP, keyby = control_crit]
use_data[, country_index := .GRP, keyby = country]
use_data[, method_index := .GRP, keyby = method]
use_data[, period_index := .GRP, keyby = period]

setkey(use_data, log_se)

stan_data_cov = list(N = nrow(use_data),
                     ses_measure_n = max(use_data$ses_index),
                     case_type_n = max(use_data$case_type_index),
                     control_type_n = max(use_data$control_type_index),
                     country_n = max(use_data$country_index),
                     period_n = max(use_data$period_index),
                     comparison_combo_n = max(use_data$comparison_type),
                     y = log(use_data$or),
                     se = use_data$log_se,
                     ses_measure = use_data$ses_index,
                     case_type = use_data$case_type_index,
                     control_type = use_data$control_type_index,
                     country = use_data$country_index,
                     period = use_data$period_index,
                     method = use_data$method_index,
                     comparison_combo = use_data$comparison_type)
```

##### Model code and prior choices

This model has two additional sets of parameters expressing the extent to which different study level indicators are associated with the outcome. The priors for the mean and standard deviation of the effect distribution remain as before.

In addition, there is a block of parameters associated with study level indicators (e.g., period, country etc). These are given the same "factor of two" prior, but normalized for each set of indicators, so that the pooled mean expresses the "average" across the countries in the sample, across the two periods, and so on.

Finally, there is a block of parameters for the combinations (interactions) of case and control outcomes. These are given a more conservative prior (normal, mean zero, standard deviation 0.2).

The estimation code:

```stan
data {
  int<lower=0> N;
  int<lower = 0> ses_measure_n;
  int<lower = 0> case_type_n;
  int<lower = 0> control_type_n;
  // int<lower = 0> comparison_combo_n;
  int<lower = 0> country_n;
  int<lower = 0> period_n;

  vector[N] y;
  vector[N] se;
  int ses_measure[N];
  int case_type[N];
```

```
  int control_type[N];
  int comparison_combo[N];
  int country[N];
  int period[N];
  int method[N];
}
transformed data {
  int method_n = 2;
  int out_n = 2* period_n * ses_measure_n * case_type_n * control_type_n * country_n;

}
parameters {
  real mu;
  real<lower = 0> sigma;

  real<lower = 0> group_sigma;
  real<lower = 0> combo_sigma;

  vector[N] study_std;
  vector[2 + period_n + ses_measure_n + case_type_n + control_type_n + country_n] group_std;
  matrix[control_type_n, case_type_n] combo_std;
}
transformed parameters {
  vector[N] study_re;
  vector[N] study_estimate;
  vector[ses_measure_n] ses_measure_re;
  vector[case_type_n] case_type_re;
  vector[control_type_n] control_type_re;
  matrix[control_type_n, case_type_n] comparison_combo_re;
  vector[country_n] country_re;
  vector[period_n] period_re;
  vector[2] method_re;

  {
    int passed_vars = 0;

    period_re = group_sigma * (group_std[(passed_vars +1):(passed_vars + period_n)] -
      mean(group_std[(passed_vars +1):(passed_vars + period_n)]));
    passed_vars += period_n;

    ses_measure_re = group_sigma * (group_std[(passed_vars +1):(passed_vars + ses_measure_n)] -
      mean(group_std[(passed_vars +1):(passed_vars + ses_measure_n)]));
    passed_vars += ses_measure_n;

    case_type_re = group_sigma * (group_std[(passed_vars +1):(passed_vars + case_type_n)] -
      mean(group_std[(passed_vars +1):(passed_vars + case_type_n)]));
    passed_vars += case_type_n;

    control_type_re = group_sigma * (group_std[(passed_vars +1):(passed_vars + control_type_n)] -
      mean(group_std[(passed_vars +1):(passed_vars + control_type_n)]));
    passed_vars += control_type_n;

    country_re = group_sigma * (group_std[(passed_vars +1):(passed_vars + country_n)] -
```

```
      mean(group_std[(passed_vars +1):(passed_vars + country_n)]));
    passed_vars += country_n;

    method_re = group_sigma * (group_std[(passed_vars +1):(passed_vars + method_n)] -
      mean(group_std[(passed_vars +1):(passed_vars + method_n)]));
    passed_vars += method_n;


  }

    comparison_combo_re = combo_sigma * (combo_std -  mean(combo_std)) +
      rep_matrix(control_type_re, case_type_n) +
      rep_matrix(to_row_vector(case_type_re), control_type_n);


  for (i in 1:N){
    study_re[i] = mu + sigma * study_std[i] +
                  ses_measure_re[ses_measure[i]] +
                  country_re[country[i]] +
                  period_re[period[i]] +
                  method_re[method[i]] +
                  comparison_combo_re[control_type[i], case_type[i]];

  }
  study_estimate = exp(study_re);

}
model {
  mu ~ normal(0, 0.4);
  sigma ~ normal(0, 0.4);
  group_sigma ~ normal(0, 0.4);
  combo_sigma ~ normal(0, 0.2);
  study_std ~ std_normal();
  group_std ~ std_normal();
  to_vector(combo_std) ~ std_normal();

  y ~ normal(study_re, se);
}
generated quantities {
  real mu_exp = exp(mu);
}
```

```
cov_bayes <- stan("random effect cov_4.stan",
                   data = stan_data_cov,
                  iter = 10000,
                  pars = c("study_std", "group_std"),
                  include = FALSE,
                  refresh = 10000,
                  control = list(adapt_delta = 0.99))
```

**Results**

```
plot(cov_bayes, pars = c("mu_exp"), plotfun = "stan_hist") + labs(title = "Effect distribution mean")
```

```
## `stat_bin()` using `bins = 30`. Pick better value with `binwidth`.
```

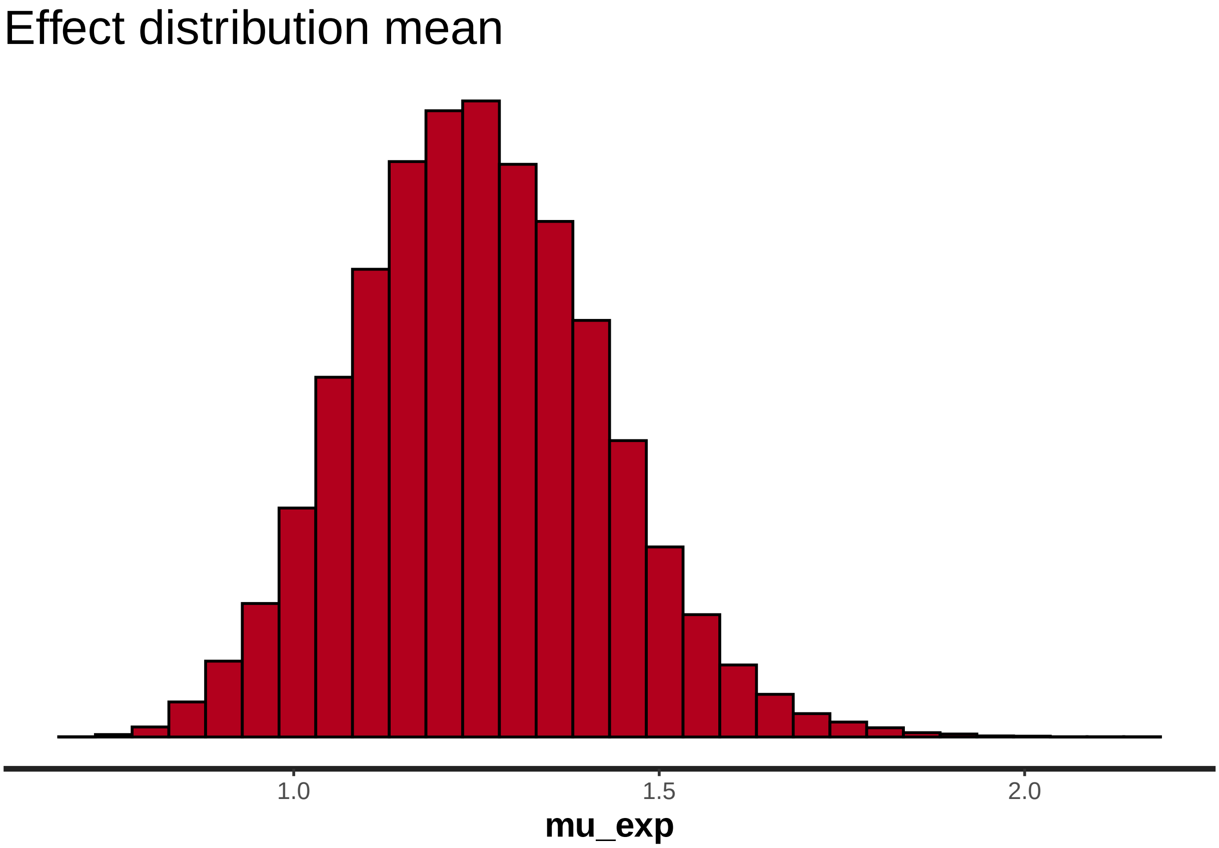


```
all_effects <- summary(cov_bayes, pars = c("ses_measure_re",
                                           # "case_type_re",
                                           # "control_type_re",
                                           "comparison_combo_re",
                                           "country_re",
                                           "period_re",
                                           "method_re"))$summary

all_effects <- cbind(data.table(all_effects),
                     data.table(variable = rownames(all_effects)))

setnames(all_effects, make.names(colnames(all_effects)))

all_effects[, variable_name := str_split_fixed(variable,  "\\[", 2)[,1]]
all_effects[, remainder := str_split_fixed(variable,  "\\[", 2)[,2]]

all_effects[ variable_name == "comparison_combo_re",
             control_index := as.integer(str_split_fixed(remainder,  ",", 2)[,1])]
all_effects[ variable_name == "comparison_combo_re",
             remainder := str_split_fixed(remainder,  ",", 2)[, 2]]
all_effects[ variable_name == "comparison_combo_re",
```

```
             case_index := as.integer(str_split_fixed(remainder,  "\\]", 2)[, 1])]

all_effects[ variable_name != "comparison_combo_re",
             variable_index := as.integer(str_split_fixed(remainder, "\\]", 2)[, 1])]

# Assigning names
temp_labels <- use_data[, list(control_crit = control_crit[1]), keyby = control_type_index]$control_cri

all_effects[ variable_name == "comparison_combo_re",
             control_criteria := temp_labels[control_index]]

temp_labels <- use_data[, list(case_crit = case_crit[1]), keyby = case_type_index]$case_crit

all_effects[ variable_name == "comparison_combo_re",
             case_criteria := temp_labels[case_index]]

all_effects[ variable_name == "comparison_combo_re",
             variable_label := paste("Control: ", control_criteria, "\n", "Case: ", case_criteria)]

temp_labels <- use_data[, list(temp = deprivation_type[1]), keyby = ses_index]$temp

all_effects[ variable_name == "ses_measure_re",
             variable_label := temp_labels[variable_index]]

temp_labels <- use_data[, list(temp = method[1]), keyby = method_index]$temp

all_effects[ variable_name == "method_re",
             variable_label := temp_labels[variable_index]]

temp_labels <- use_data[, list(temp = country[1]), keyby = country_index]$temp

all_effects[ variable_name == "country_re",
             variable_label := temp_labels[variable_index]]

temp_labels <- use_data[, list(temp = period[1]), keyby = period_index]$temp

all_effects[ variable_name == "period_re",
             variable_label := temp_labels[variable_index]]

all_effects[, variable_fact := factor(variable_name,
                                      levels = c("ses_measure_re",
                                                 "comparison_combo_re",
                                                 "country_re",
                                                 "period_re",
                                                 "method_re"),
                                      labels = c("Level of SES measure",
                                                 "Type of comparison",
                                                 "Country/region",
                                                 "Period",
                                                 "Method"))]

ggplot(all_effects[variable_name != "comparison_combo_re"],
       aes(variable, exp(mean), col = variable_fact)) +
```

```
  geom_point() +
  geom_errorbar(aes(ymin = exp(X2.5.),
                    ymax = exp(X97.5.))) +
  geom_hline(yintercept = 1, linetype = "dashed") +
  # facet_wrap(~variable_name) +
  scale_x_discrete(breaks = all_effects$variable,
                   labels = all_effects$variable_label) +
  coord_flip() +
  labs(y = "Odds ratio relative to average across distinction",
       x = "") +
  guides(col = guide_legend(title = "Variable group",
                            reverse = T))
```

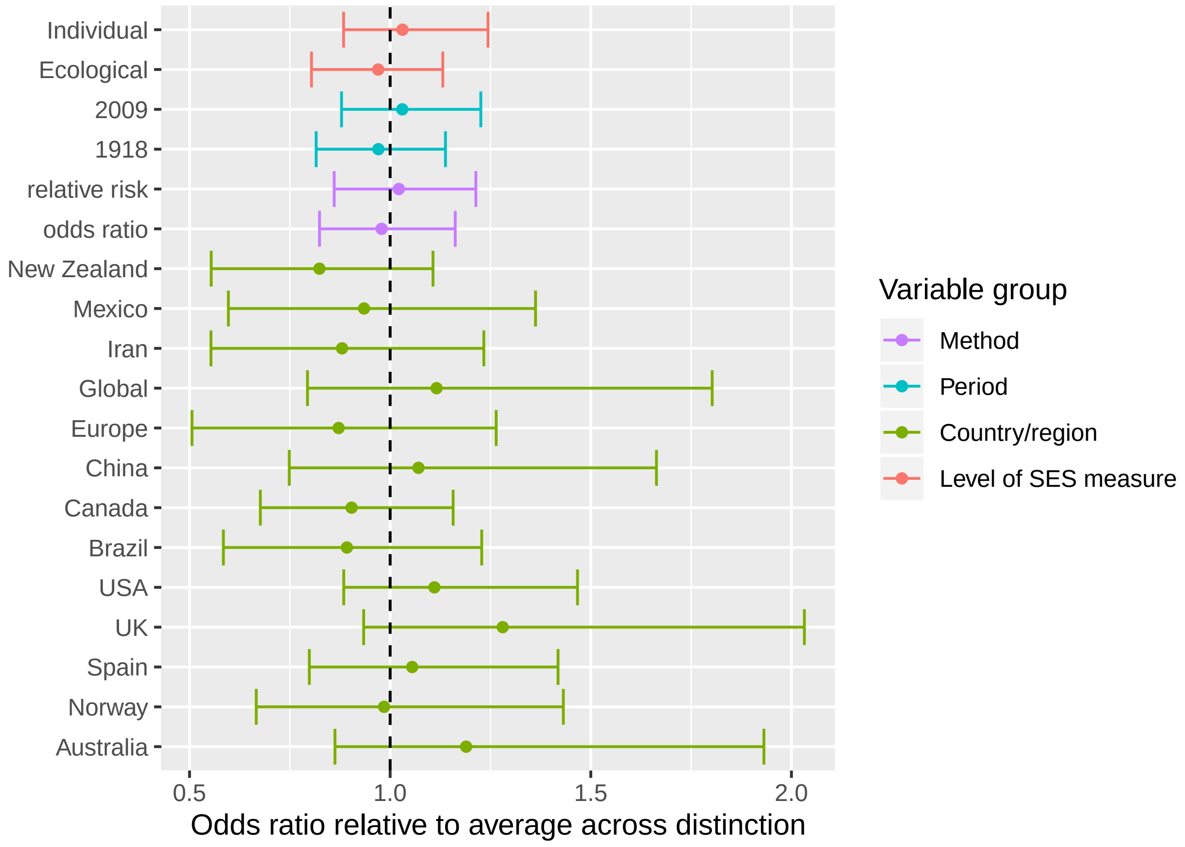


```
  NULL
```

```
## NULL
```

```
combos_seen <- unique(use_data[, paste0(control_type_index,"-",case_type_index)])
all_effects[variable_name == "comparison_combo_re",
            in_data := paste0(control_index, "-",case_index) %in% combos_seen]

all_effects[variable_name == "comparison_combo_re" & in_data == T,
            combo_number := case_when(control_index == 2 & case_index == 2 ~ 10,
                                      control_index == 2 & case_index == 1 ~ 9,
                                      control_index == 2 & case_index == 3 ~ 8,
                                      control_index == 3 & case_index == 1 ~ 7,
                                      control_index == 3 & case_index == 4 ~ 6,
                                      control_index == 3 & case_index == 3 ~ 5,
```

```
                                        control_index == 1 & case_index == 4 ~ 4,
                                        control_index == 1 & case_index == 3 ~ 3,
                                        control_index == 4 & case_index == 2 ~ 2,
                                        control_index == 4 & case_index == 3 ~ 1,
                                        TRUE ~ NA_real_)]

ggplot(all_effects[variable_name == "comparison_combo_re" & in_data == T],
       aes(combo_number, exp(mean))) +
  geom_point() +
  geom_errorbar(aes(ymin = exp(X2.5.),
                    ymax = exp(X97.5.))) +
  geom_hline(yintercept = 1, linetype = "dashed") +
  # facet_wrap(~variable_name) +
  scale_x_continuous(breaks = all_effects$combo_number,
                     labels = all_effects$variable_label) +
  coord_flip() +
  labs(y = "Odds ratio relative to average across all comparisons",
       x = "")
```

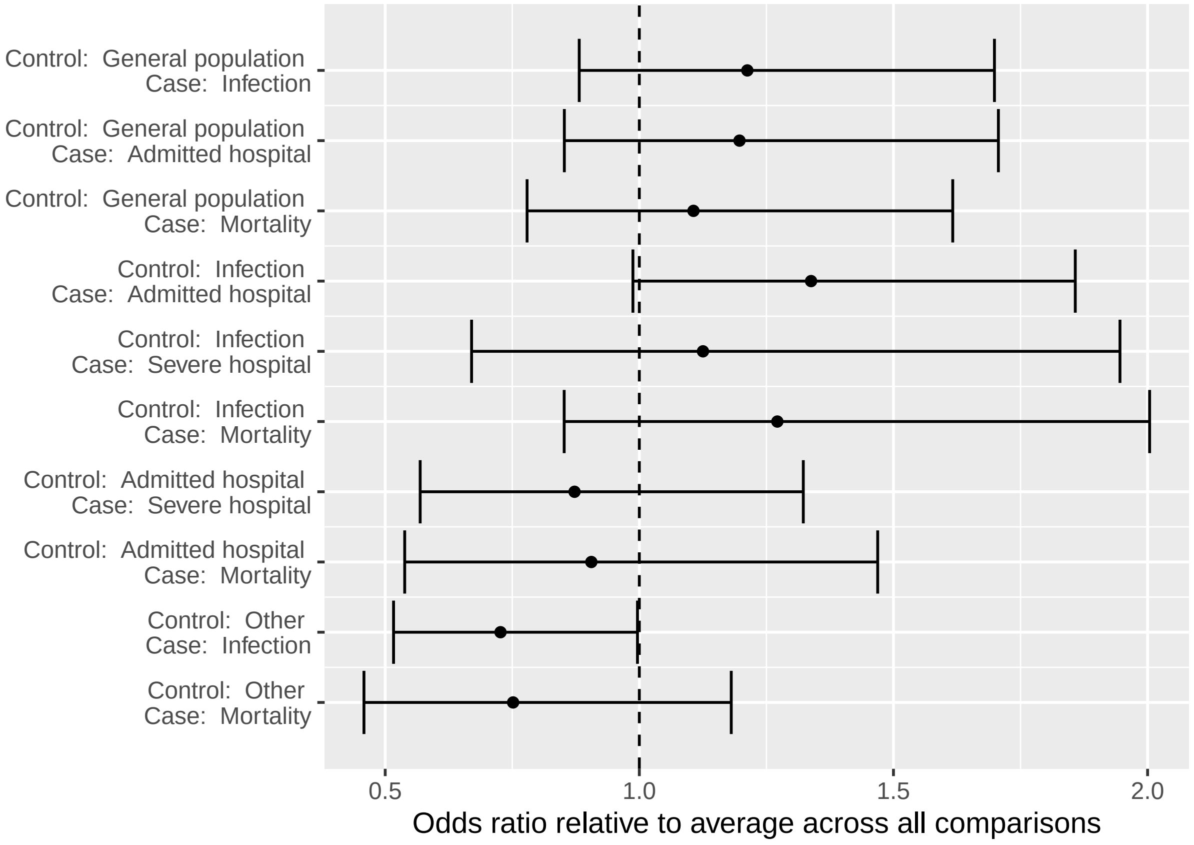


#### Comparing bayesian models

```
round(summary(base_bayes, pars = c("mu_exp", "sigma"))$summary, 2)
```

```
##        mean se_mean   sd 2.5%  25%  50% 75% 97.5%   n_eff Rhat
## mu_exp 1.42       0 0.11 1.21 1.35 1.42 1.5  1.66 3925.96    1
```

```
## sigma  0.46       0 0.07 0.33 0.41 0.45 0.5  0.61 4739.25    1
```

```
round(summary(cov_bayes, pars = c("mu_exp", "sigma", "group_sigma", "combo_sigma"))$summary, 2)
```

```
##             mean se_mean   sd 2.5%  25%  50%  75% 97.5%    n_eff Rhat
## mu_exp      1.25       0 0.17 0.94 1.13 1.25 1.36  1.61 10527.12    1
## sigma       0.33       0 0.08 0.20 0.28 0.33 0.38  0.50  5607.20    1
## group_sigma 0.21       0 0.10 0.02 0.14 0.20 0.27  0.42  3716.73    1
## combo_sigma 0.14       0 0.10 0.01 0.06 0.13 0.21  0.37 10614.31    1
```

```
plot(base_bayes, pars = c( "sigma"), plotfun = "stan_hist")
```

```
## `stat_bin()` using `bins = 30`. Pick better value with `binwidth`.
```

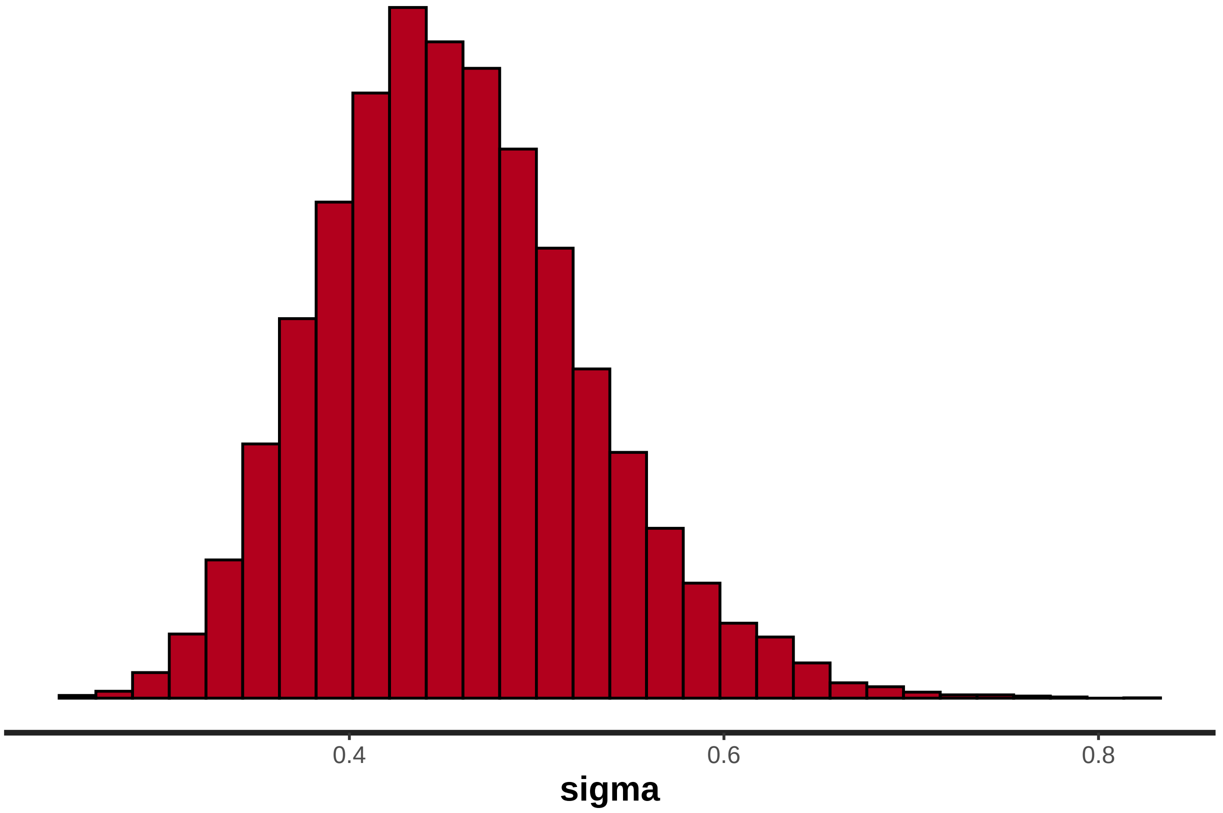


```
plot(cov_bayes, pars = c("sigma", "group_sigma", "combo_sigma"), plotfun = "stan_hist")
```

```
## `stat_bin()` using `bins = 30`. Pick better value with `binwidth`.
```

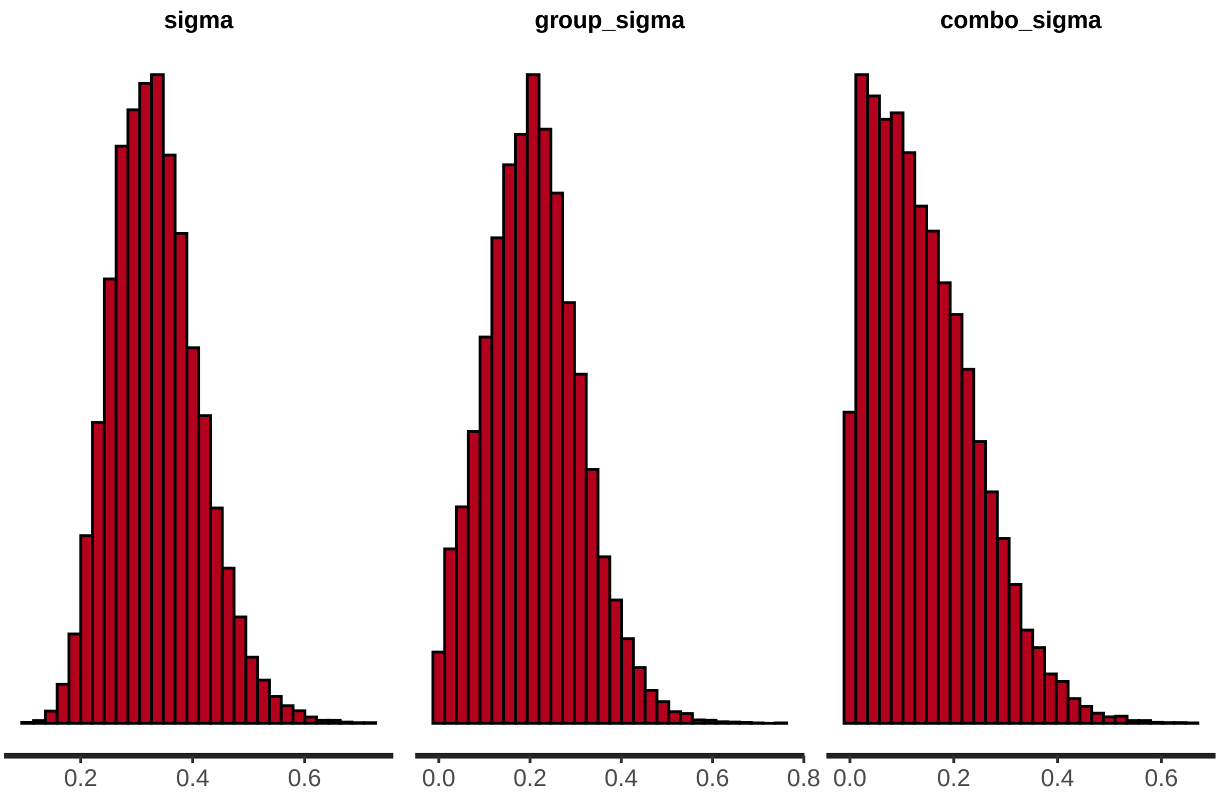


**Comparing the priors to the posteriors**

```
compare_het <- rbind(data.table(model = "Without study level variables",
                                posterior_draw = extract(base_bayes, pars = c("sigma"))$sigma),
                     data.table(model = "With study level variables",
                                posterior_draw = extract(cov_bayes, pars = c("sigma"))$sigma))

ggplot(compare_het, aes(posterior_draw, fill = model)) +
  geom_histogram(position = "dodge")  +
  labs(title = "Unexplained heterogeneity (tau)")
```

```
## `stat_bin()` using `bins = 30`. Pick better value with `binwidth`.
```

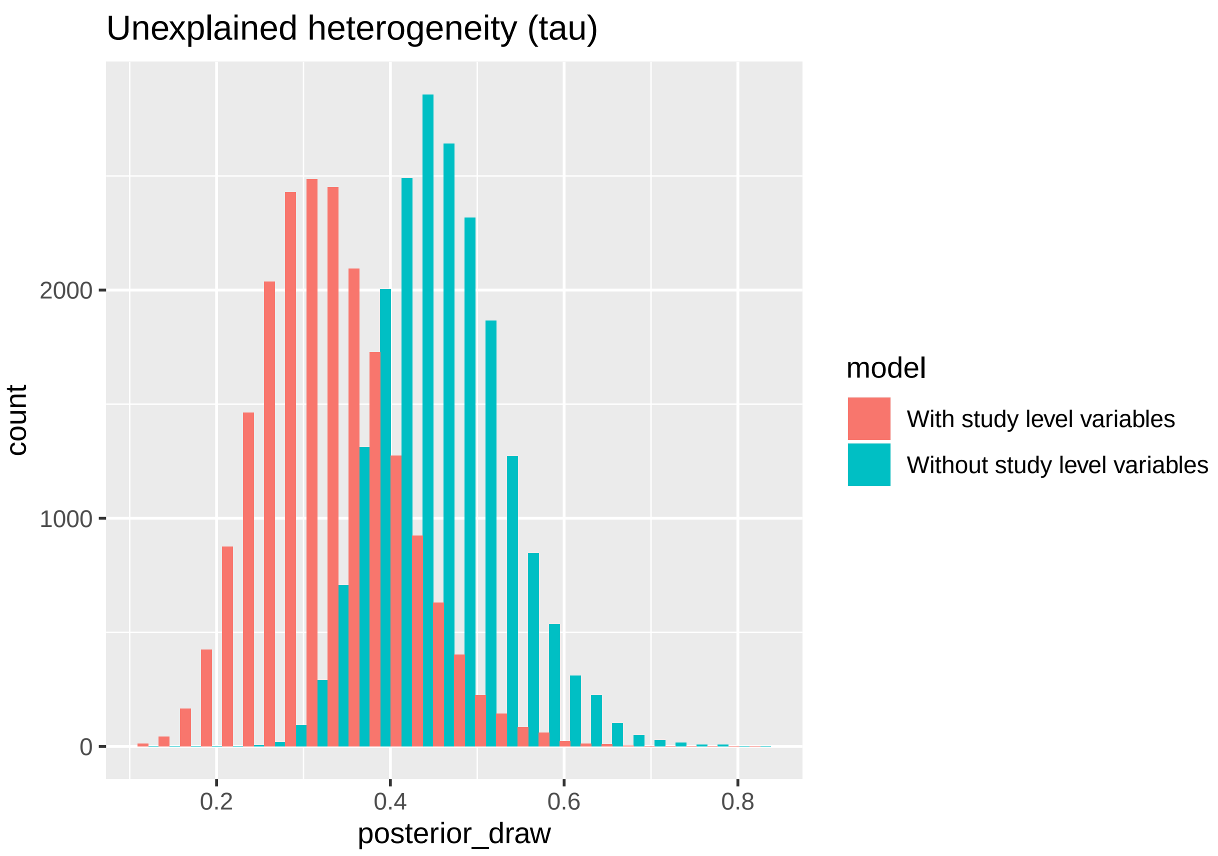


```
prior_draws <- rnorm(100000, 0, 0.4)
prior_draws <- prior_draws[prior_draws > 0][1:20000]
compare_prior_post <- rbind(data.table(type = "Posterior",
                                       parameter_value = extract(cov_bayes, pars = c("sigma"))$sigma),
                             data.table(type = "Prior",
                                       parameter_value = prior_draws))

ggplot(compare_prior_post, aes(parameter_value, fill = type)) +
  geom_histogram(position = "dodge") +
  labs(title = "Prior and posterior distribution - unexplained study heterogeneity (tau)")
```

```
## `stat_bin()` using `bins = 30`. Pick better value with `binwidth`.
```

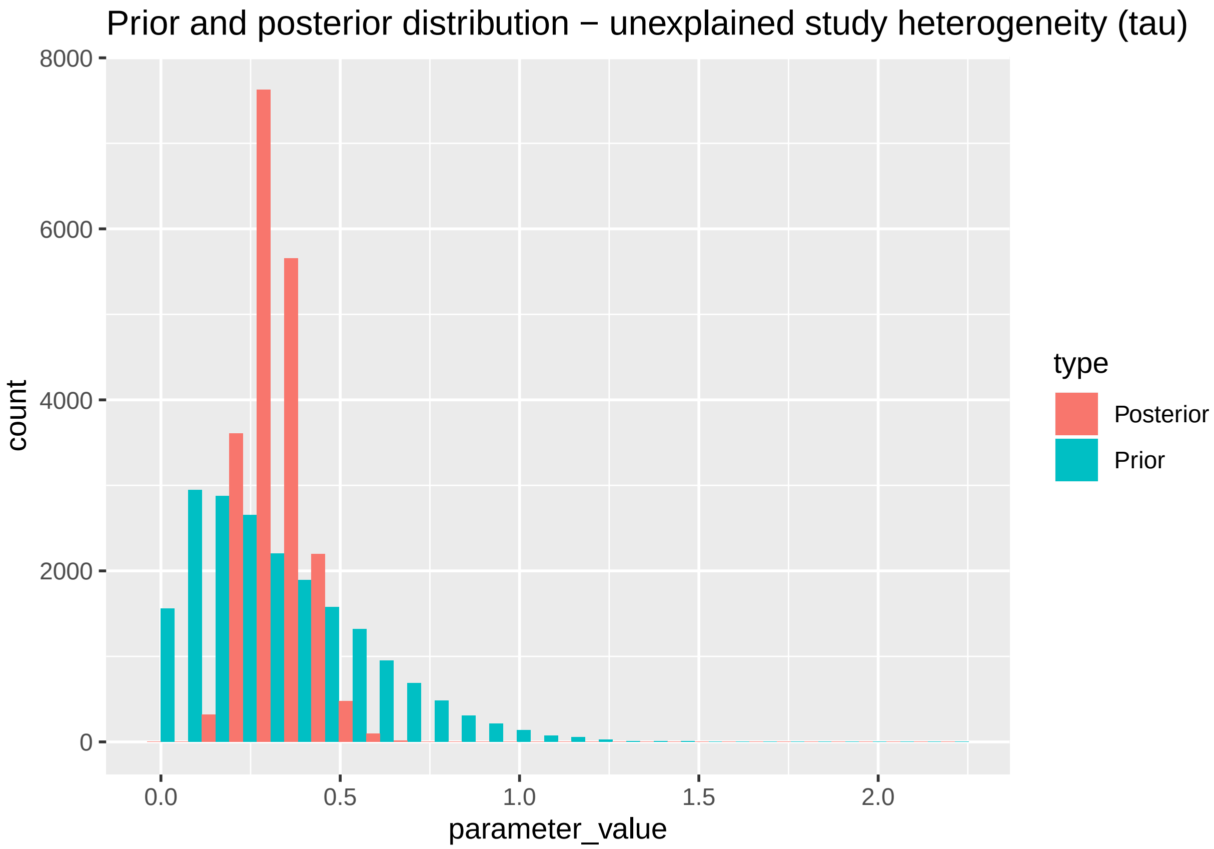


```
prior_draws <- rnorm(100000, 0, 0.4)
prior_draws <- prior_draws[prior_draws > 0][1:20000]
compare_prior_post <- rbind(data.table(type = "Posterior",
                                       parameter_value = extract(cov_bayes, pars = c("group_sigma"))$gr
                            data.table(type = "Prior",
                                       parameter_value = prior_draws))

ggplot(compare_prior_post, aes(parameter_value, fill = type)) +
  geom_histogram(position = "dodge") +
  labs(title = "Prior and posterior distribution - sigma for study level indicators")
```

```
## `stat_bin()` using `bins = 30`. Pick better value with `binwidth`.
```

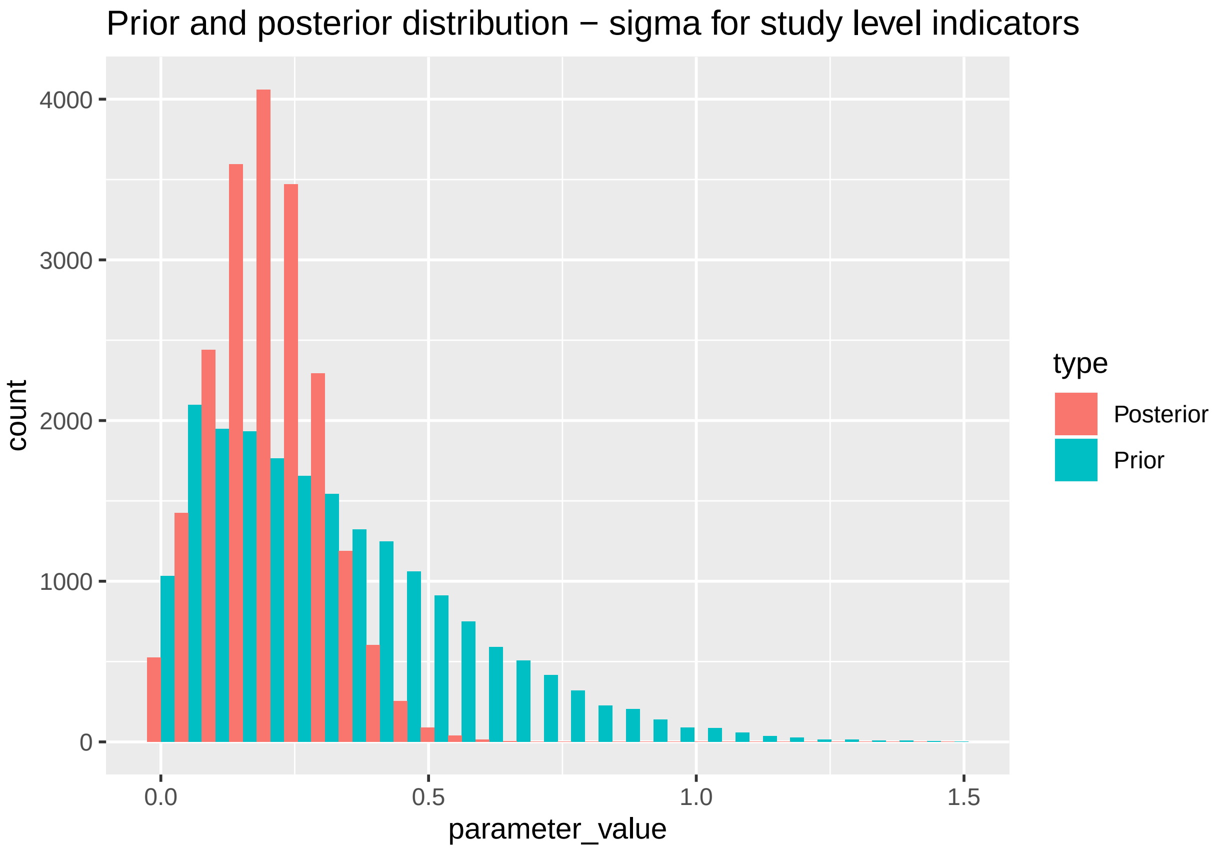


```
prior_draws <- rnorm(100000, 0, 0.2)
prior_draws <- prior_draws[prior_draws > 0][1:20000]
compare_prior_post <- rbind(data.table(type = "Posterior",
                                       parameter_value = extract(cov_bayes, pars = c("combo_sigma"))$com
                            data.table(type = "Prior",
                                       parameter_value = prior_draws))

ggplot(compare_prior_post, aes(parameter_value, fill = type)) +
  geom_histogram(position = "dodge") +
  labs(title = "Prior and posterior distribution - sigma for comparison combinations")
```

```
## `stat_bin()` using `bins = 30`. Pick better value with `binwidth`.
```

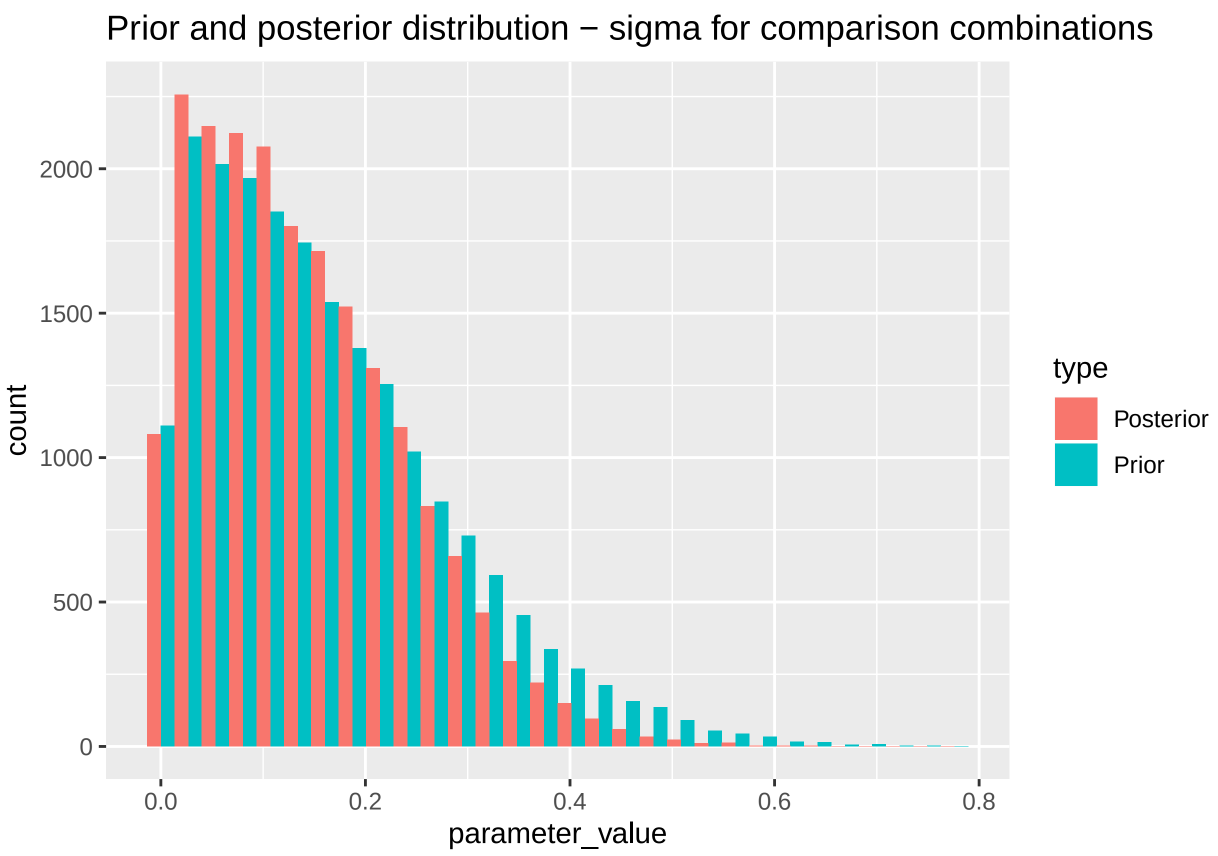


#### Subsamples

##### By case outcome_type

```
measures <- unique(use_data$case_crit)


rm_list <- list()

for (i in 1:length(measures)){
  rm_list[[i]] <- random_effects <- rma(yi = log_or,
                                        sei = log_se,
                                        data = meta_dt[case_crit == measures[i]])
  cat("Results for studies with case based on ", measures[i], "\n")
  print(rm_list[i])
  forest(rm_list[[i]],
               atransf = exp,
               order = "prec",
               slab = meta_dt[case_crit == measures[i], list(v = paste0(Author, ", ", Year))]$v,
         main = paste0("Type of case outcome: ",measures[i] ))
}
```

```
## Results for studies with case based on  Mortality
## [[1]]
##
```

```
## Random-Effects Model (k = 13; tau^2 estimator: REML)
##
## tau^2 (estimated amount of total heterogeneity): 0.2018 (SE = 0.1072)
## tau (square root of estimated tau^2 value):      0.4492
## I^2 (total heterogeneity / total variability):   93.62%
## H^2 (total variability / sampling variability):  15.68
##
## Test for Heterogeneity:
## Q(df = 12) = 63.3896, p-val < .0001
##
## Model Results:
##
## estimate      se    zval    pval   ci.lb   ci.ub
##   0.3502  0.1440  2.4314  0.0150  0.0679  0.6325  *
##
## ---
## Signif. codes:  0 '***' 0.001 '**' 0.01 '*' 0.05 '.' 0.1 ' ' 1
```

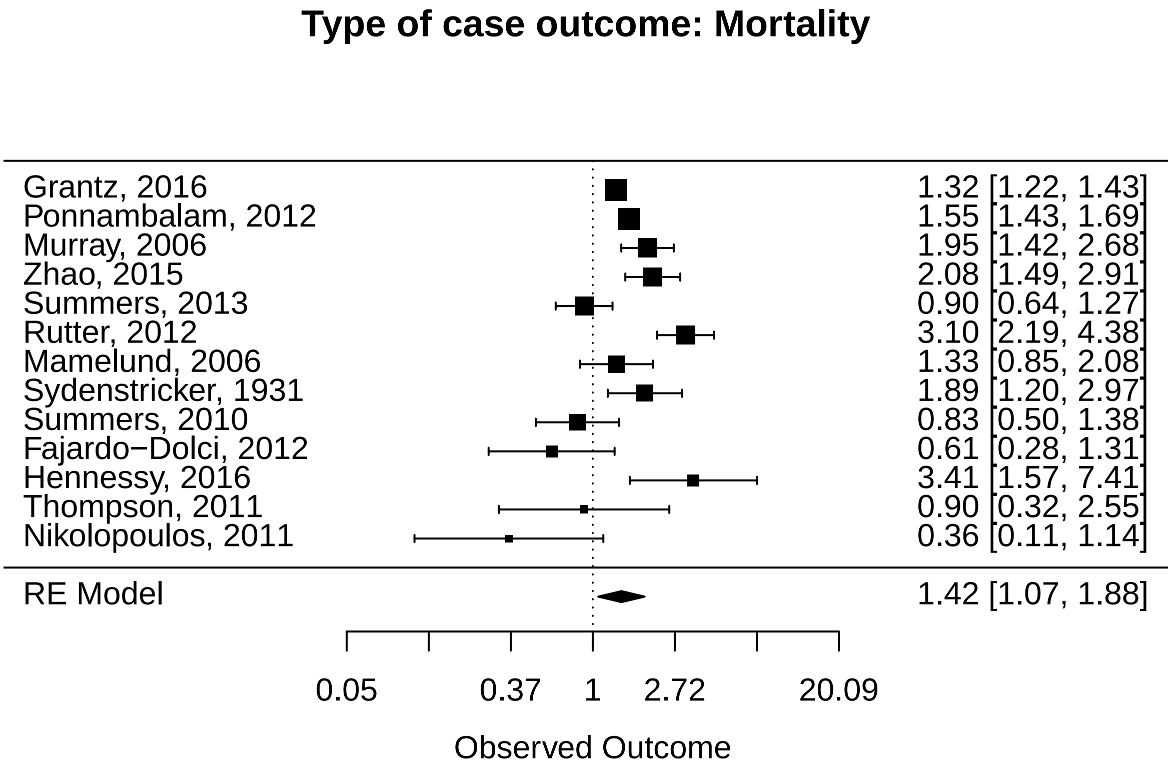


```
## Results for studies with case based on  Infection
## [[1]]
##
## Random-Effects Model (k = 14; tau^2 estimator: REML)
##
## tau^2 (estimated amount of total heterogeneity): 0.2723 (SE = 0.1446)
## tau (square root of estimated tau^2 value):      0.5219
## I^2 (total heterogeneity / total variability):   93.18%
## H^2 (total variability / sampling variability):  14.66
##
## Test for Heterogeneity:
## Q(df = 13) = 137.3100, p-val < .0001
```

```
##
## Model Results:
##
## estimate      se    zval    pval    ci.lb   ci.ub
##   0.2000  0.1664  1.2020  0.2294  -0.1261  0.5262
##
## ---
## Signif. codes:  0 '***' 0.001 '**' 0.01 '*' 0.05 '.' 0.1 ' ' 1
```

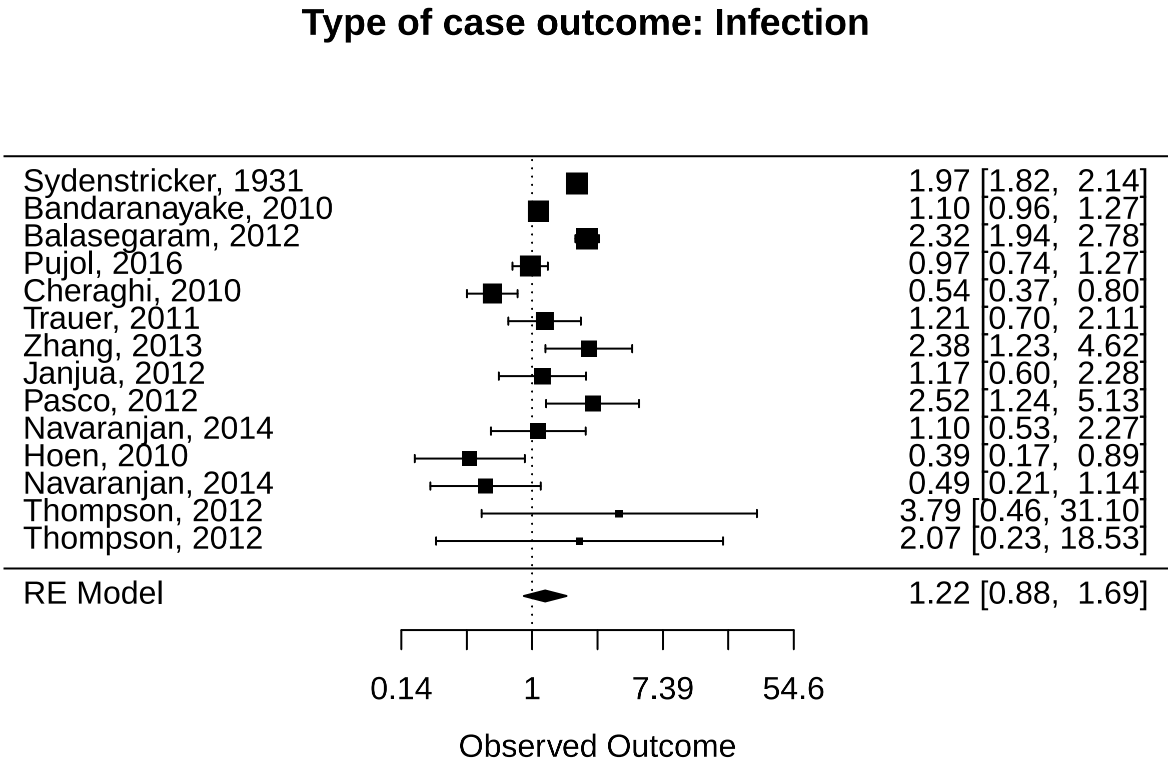


```
## Results for studies with case based on  Admitted hospital
## [[1]]
##
## Random-Effects Model (k = 15; tau^2 estimator: REML)
##
## tau^2 (estimated amount of total heterogeneity): 0.1656 (SE = 0.0857)
## tau (square root of estimated tau^2 value):      0.4069
## I^2 (total heterogeneity / total variability):   85.33%
## H^2 (total variability / sampling variability):  6.82
##
## Test for Heterogeneity:
## Q(df = 14) = 93.0124, p-val < .0001
##
## Model Results:
##
## estimate      se    zval    pval   ci.lb   ci.ub
##   0.5573  0.1247  4.4687  <.0001  0.3129  0.8017  ***
##
## ---
## Signif. codes:  0 '***' 0.001 '**' 0.01 '*' 0.05 '.' 0.1 ' ' 1
```

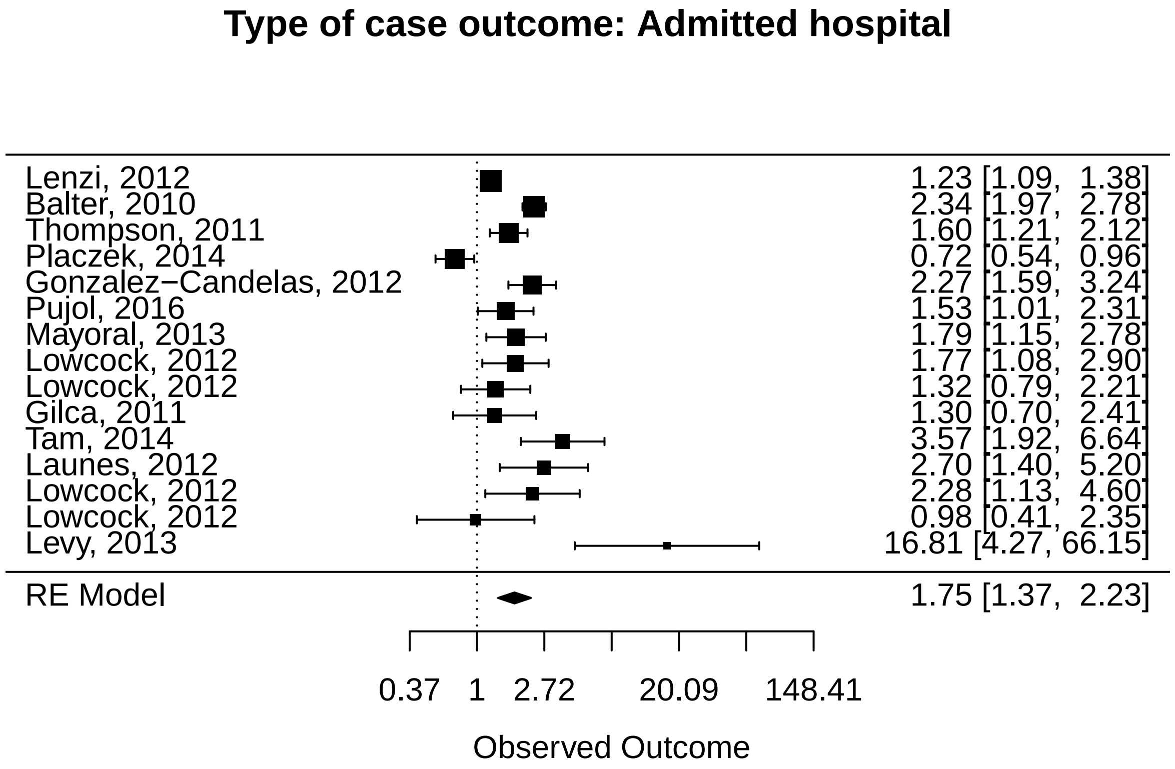


```
## Results for studies with case based on  Severe hospital
## [[1]]
##
## Random-Effects Model (k = 4; tau^2 estimator: REML)
##
## tau^2 (estimated amount of total heterogeneity): 0 (SE = 0.2158)
## tau (square root of estimated tau^2 value):      0
## I^2 (total heterogeneity / total variability):   0.00%
## H^2 (total variability / sampling variability):  1.00
##
## Test for Heterogeneity:
## Q(df = 3) = 1.4244, p-val = 0.6998
##
## Model Results:
##
## estimate      se     zval    pval    ci.lb   ci.ub
##  -0.0911  0.2543  -0.3582  0.7202  -0.5896  0.4074
##
## ---
## Signif. codes:  0 '***' 0.001 '**' 0.01 '*' 0.05 '.' 0.1 ' ' 1
```

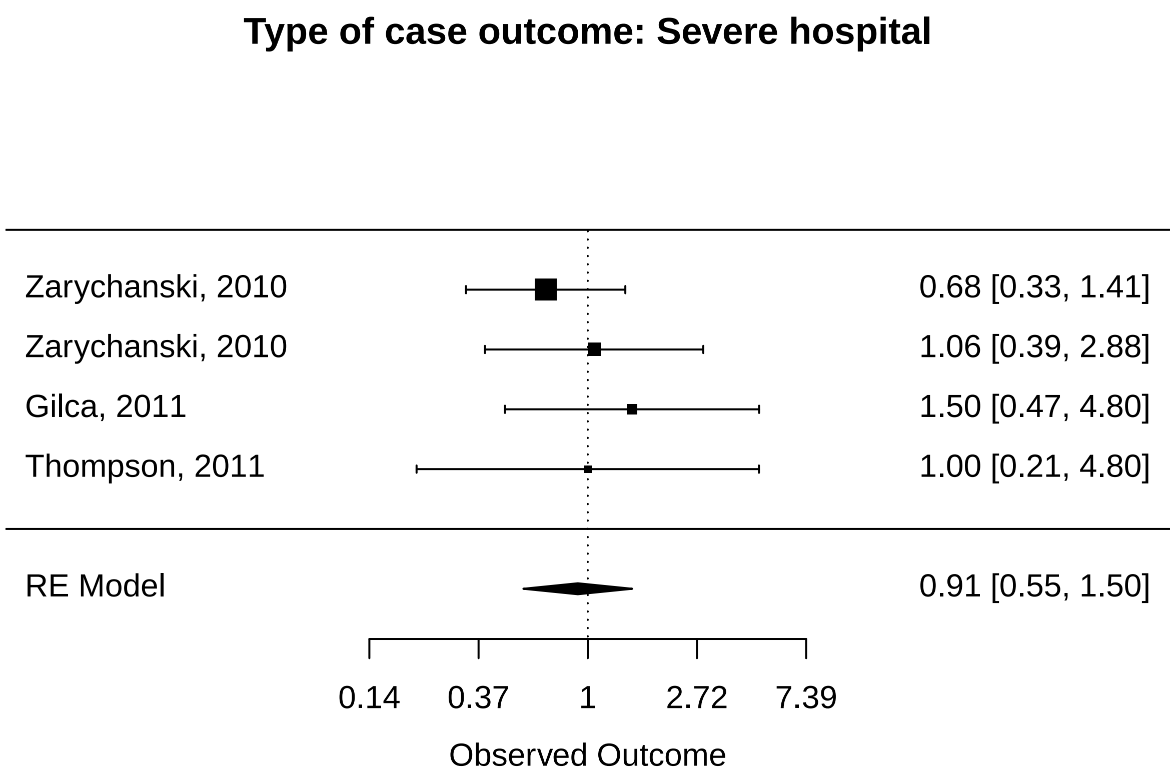


##### By control outcome_type

```
measures <- unique(use_data$control_crit)


rm_list <- list()

for (i in 1:length(measures)){
  rm_list[[i]] <- random_effects <- rma(yi = log_or,
                                        sei = log_se,
                                        data = meta_dt[control_crit == measures[i]])
  cat("Results for studies with SES-index based on ", measures[i], "\n")
  print(rm_list[i])
  forest(rm_list[[i]],
               atransf = exp,
               order = "prec",
               slab = meta_dt[control_crit == measures[i], list(v = paste0(Author, ", ", Year))]$v,
        main = paste0("Type of control outcome: ",measures[i] ))
}
```

```
## Results for studies with SES-index based on  General population
## [[1]]
##
## Random-Effects Model (k = 19; tau^2 estimator: REML)
##
## tau^2 (estimated amount of total heterogeneity): 0.1747 (SE = 0.0708)
## tau (square root of estimated tau^2 value):      0.4180
## I^2 (total heterogeneity / total variability):   95.25%
```

```
## H^2 (total variability / sampling variability):  21.03
##
## Test for Heterogeneity:
## Q(df = 18) = 193.0825, p-val < .0001
##
## Model Results:
##
## estimate      se    zval    pval   ci.lb   ci.ub
##   0.4572  0.1068  4.2787  <.0001  0.2477  0.6666  ***
##
## ---
## Signif. codes:  0 '***' 0.001 '**' 0.01 '*' 0.05 '.' 0.1 ' ' 1
```

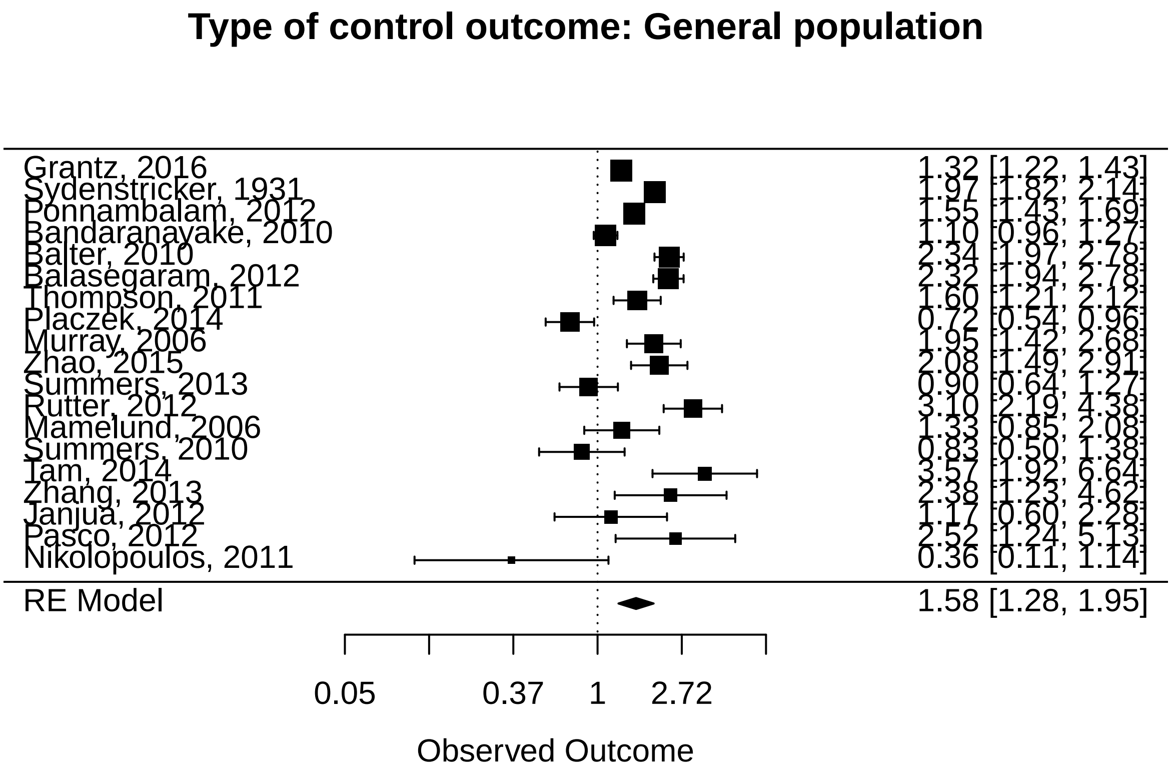


```
## Results for studies with SES-index based on  Infection
## [[1]]
##
## Random-Effects Model (k = 14; tau^2 estimator: REML)
##
## tau^2 (estimated amount of total heterogeneity): 0.0603 (SE = 0.0506)
## tau (square root of estimated tau^2 value):      0.2455
## I^2 (total heterogeneity / total variability):   54.93%
## H^2 (total variability / sampling variability):  2.22
##
## Test for Heterogeneity:
## Q(df = 13) = 40.0613, p-val = 0.0001
##
## Model Results:
##
## estimate      se    zval    pval   ci.lb   ci.ub
##   0.5614  0.0998  5.6258  <.0001  0.3658  0.7570  ***
```

```
##
## ---
## Signif. codes:  0 '***' 0.001 '**' 0.01 '*' 0.05 '.' 0.1 ' ' 1
```

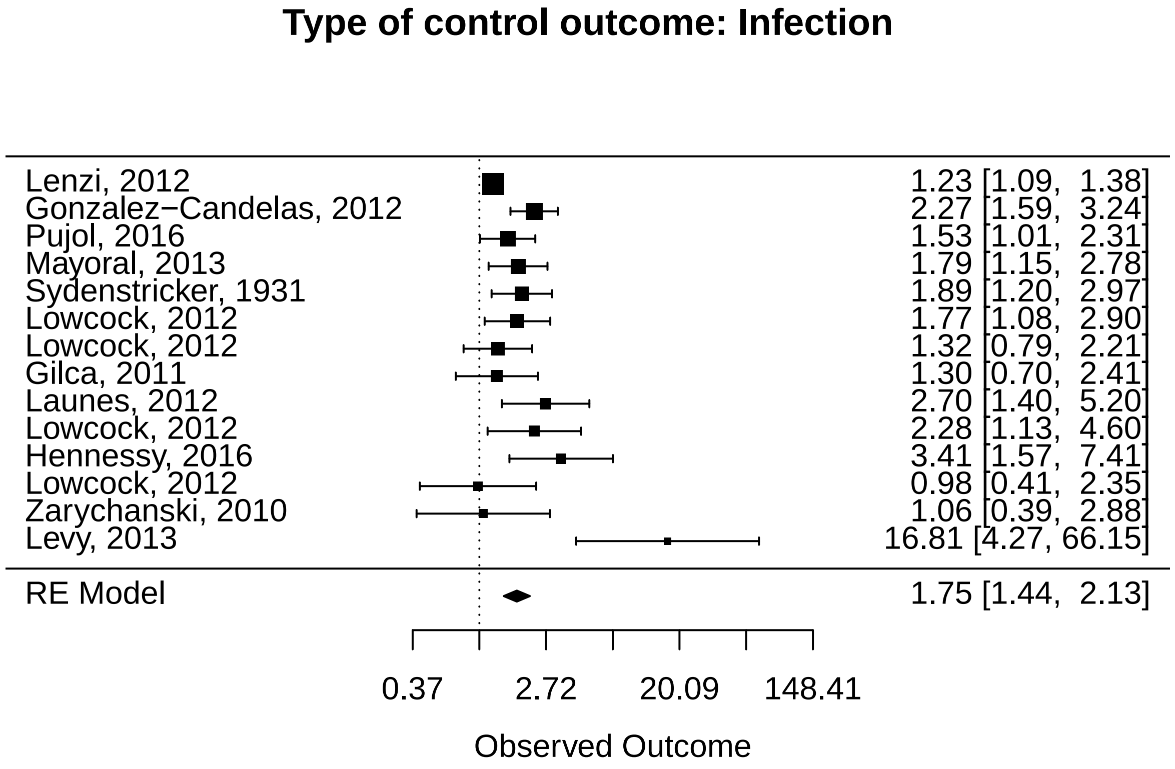


```
## Results for studies with SES-index based on  Other
## [[1]]
##
## Random-Effects Model (k = 9; tau^2 estimator: REML)
##
## tau^2 (estimated amount of total heterogeneity): 0.0868 (SE = 0.0991)
## tau (square root of estimated tau^2 value):      0.2946
## I^2 (total heterogeneity / total variability):   48.06%
## H^2 (total variability / sampling variability):  1.93
##
## Test for Heterogeneity:
## Q(df = 8) = 16.1789, p-val = 0.0399
##
## Model Results:
##
## estimate      se     zval    pval    ci.lb   ci.ub
##  -0.2486  0.1562  -1.5911  0.1116  -0.5548  0.0576
##
## ---
## Signif. codes:  0 '***' 0.001 '**' 0.01 '*' 0.05 '.' 0.1 ' ' 1
```

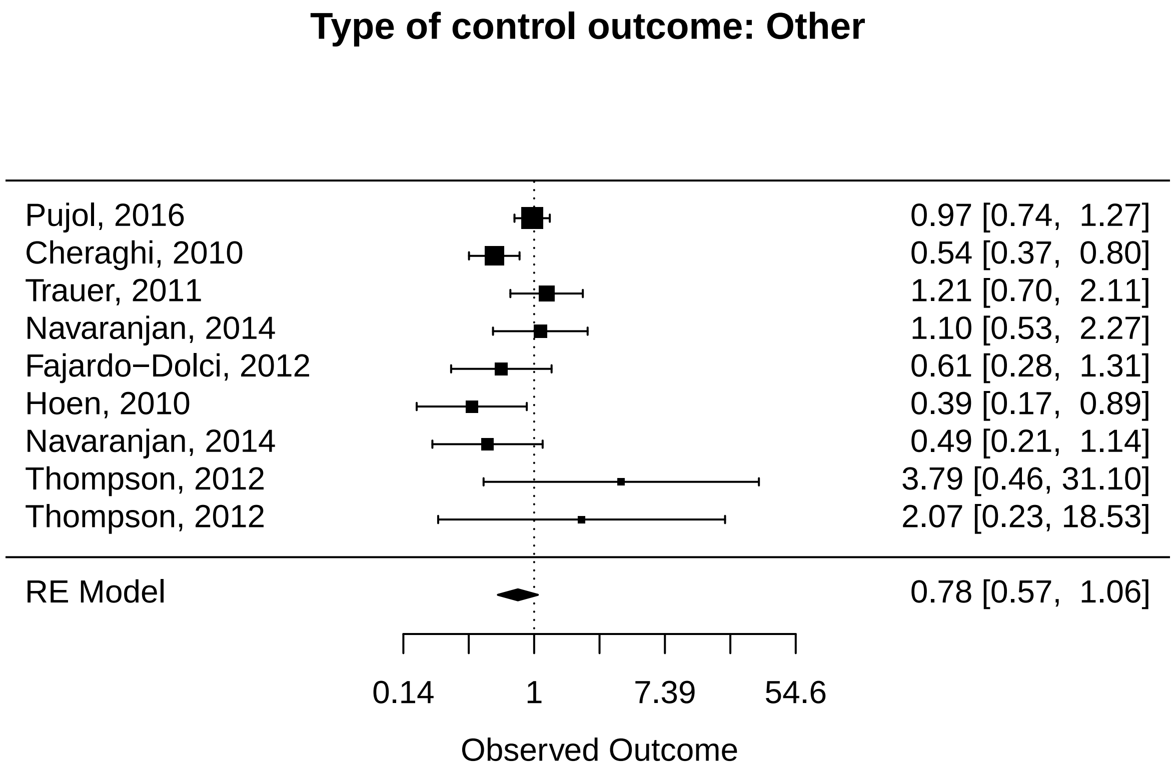


```
## Results for studies with SES-index based on  Admitted hospital
## [[1]]
##
## Random-Effects Model (k = 4; tau^2 estimator: REML)
##
## tau^2 (estimated amount of total heterogeneity): 0 (SE = 0.2214)
## tau (square root of estimated tau^2 value):      0
## I^2 (total heterogeneity / total variability):   0.00%
## H^2 (total variability / sampling variability):  1.00
##
## Test for Heterogeneity:
## Q(df = 3) = 1.3136, p-val = 0.7259
##
## Model Results:
##
## estimate      se     zval    pval    ci.lb   ci.ub
##  -0.1323  0.2568  -0.5153  0.6064  -0.6355  0.3709
##
## ---
## Signif. codes:  0 '***' 0.001 '**' 0.01 '*' 0.05 '.' 0.1 ' ' 1
```

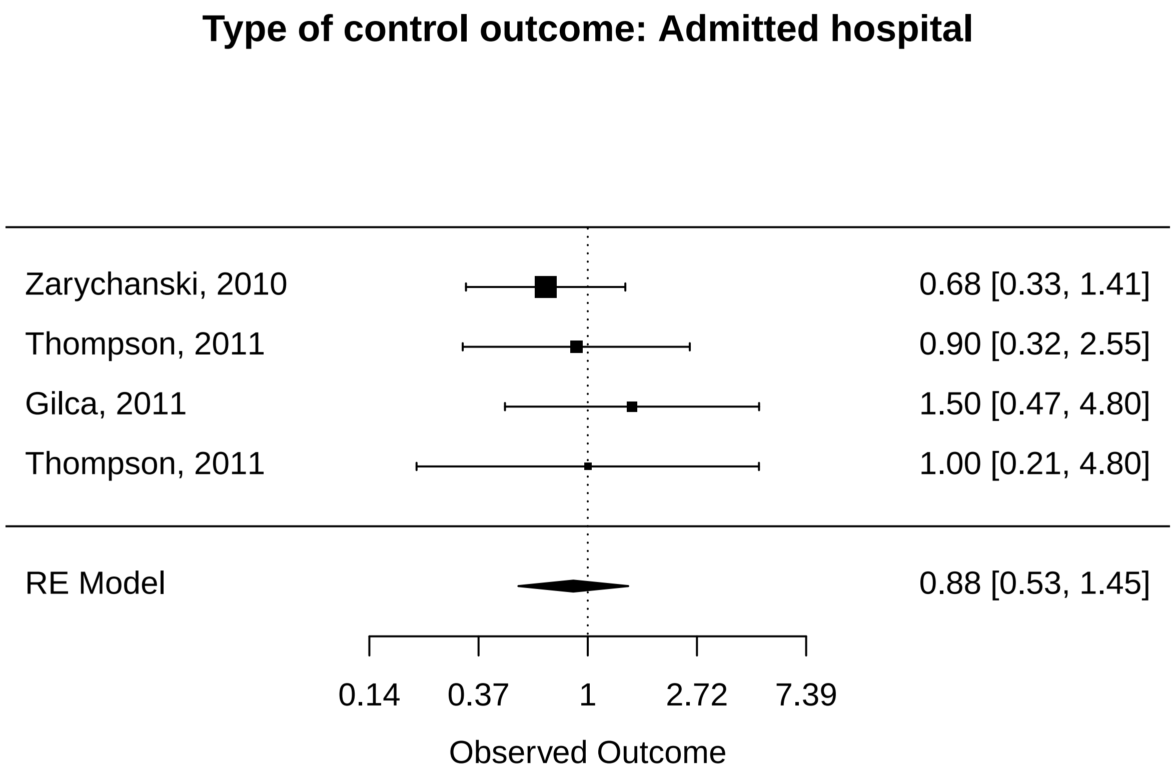


##### By period

```
measures <- unique(use_data$period)


rm_list <- list()

for (i in 1:length(measures)){
  rm_list[[i]] <- random_effects <- rma(yi = log_or,
                                        sei = log_se,
                                        data = meta_dt[period == measures[i]])
  cat("Results for studies with SES-index based on ", measures[i], "\n")
  print(rm_list[i])
  forest(rm_list[[i]],
               atransf = exp,
               order = "prec",
               slab = meta_dt[period == measures[i], list(v = paste0(Author, ", ", Year))]$v,
        main = paste0("Period: ",measures[i] ))
}
```

```
## Results for studies with SES-index based on  1918
## [[1]]
##
## Random-Effects Model (k = 7; tau^2 estimator: REML)
##
## tau^2 (estimated amount of total heterogeneity): 0.0915 (SE = 0.0687)
## tau (square root of estimated tau^2 value):      0.3024
## I^2 (total heterogeneity / total variability):   92.31%
```

```
## H^2 (total variability / sampling variability):  13.01
##
## Test for Heterogeneity:
## Q(df = 6) = 68.3824, p-val < .0001
##
## Model Results:
##
## estimate      se    zval    pval   ci.lb   ci.ub
##   0.3484  0.1310  2.6604  0.0078  0.0917  0.6051  **
##
## ---
## Signif. codes:  0 '***' 0.001 '**' 0.01 '*' 0.05 '.' 0.1 ' ' 1
```

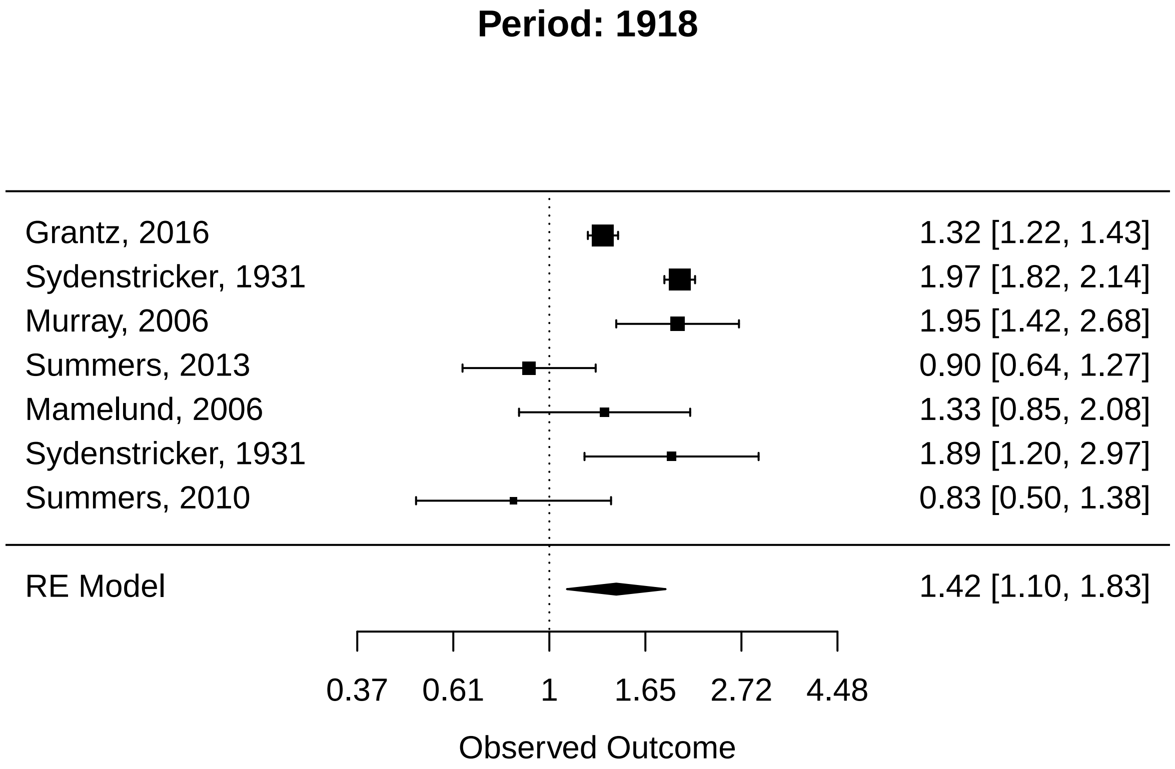


```
## Results for studies with SES-index based on  2009
## [[1]]
##
## Random-Effects Model (k = 39; tau^2 estimator: REML)
##
## tau^2 (estimated amount of total heterogeneity): 0.2509 (SE = 0.0793)
## tau (square root of estimated tau^2 value):      0.5009
## I^2 (total heterogeneity / total variability):   90.70%
## H^2 (total variability / sampling variability):  10.76
##
## Test for Heterogeneity:
## Q(df = 38) = 236.6249, p-val < .0001
##
## Model Results:
##
## estimate      se    zval    pval   ci.lb   ci.ub
##   0.3680  0.0963  3.8211  0.0001  0.1792  0.5568  ***
```

```
##
## ---
## Signif. codes:  0 '***' 0.001 '**' 0.01 '*' 0.05 '.' 0.1 ' ' 1
```

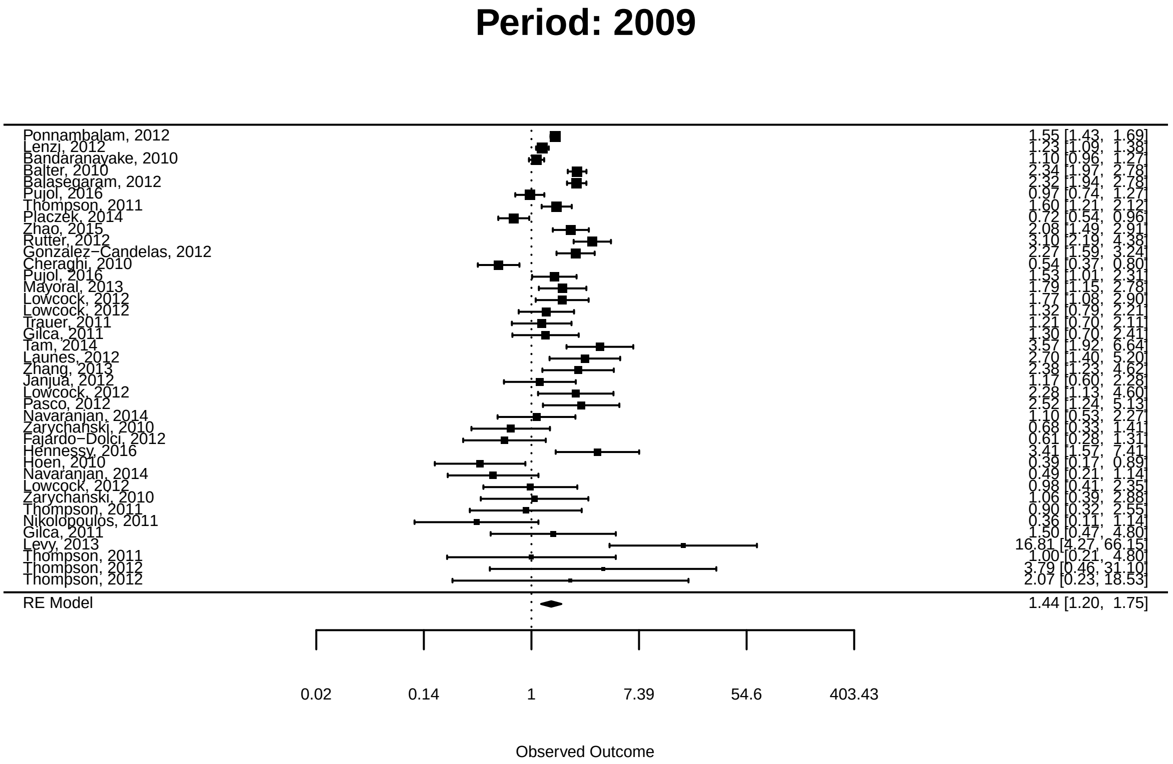


#### By method

```
measures <- unique(meta_dt$method)


rm_list <- list()

for (i in 1:length(measures)){
  rm_list[[i]] <- random_effects <- rma(yi = log_or,
                                        sei = log_se,
                                        data = meta_dt[method == measures[i]])
  cat("Results for studies with SES-index based on ", measures[i], "\n")
  print(rm_list[i])
  forest(rm_list[[i]],
              atransf = exp,
              order = "prec",
              slab = meta_dt[method == measures[i], list(v = paste0(Author, ", ", Year))]$v,
         main = paste0("Measure type: ",measures[i] ))
}
```

```
## Results for studies with SES-index based on  relative risk
## [[1]]
##
## Random-Effects Model (k = 10; tau^2 estimator: REML)
```

```
##
## tau^2 (estimated amount of total heterogeneity): 0.1252 (SE = 0.0723)
## tau (square root of estimated tau^2 value):      0.3538
## I^2 (total heterogeneity / total variability):   93.41%
## H^2 (total variability / sampling variability):  15.18
##
## Test for Heterogeneity:
## Q(df = 9) = 70.7540, p-val < .0001
##
## Model Results:
##
## estimate      se    zval    pval   ci.lb   ci.ub
##   0.4759  0.1252  3.7999  0.0001  0.2304  0.7214  ***
##
## ---
## Signif. codes:  0 '***' 0.001 '**' 0.01 '*' 0.05 '.' 0.1 ' ' 1
```

**Measure type: relative risk**

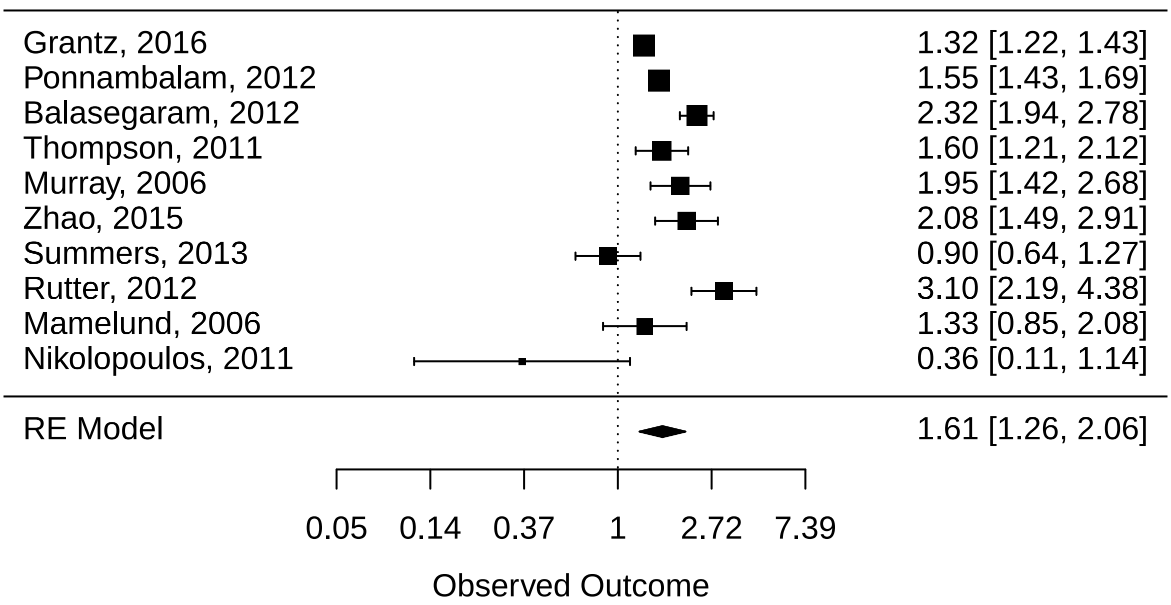


```
## Results for studies with SES-index based on  odds ratio
## [[1]]
##
## Random-Effects Model (k = 36; tau^2 estimator: REML)
##
## tau^2 (estimated amount of total heterogeneity): 0.2361 (SE = 0.0802)
## tau (square root of estimated tau^2 value):      0.4859
## I^2 (total heterogeneity / total variability):   89.61%
## H^2 (total variability / sampling variability):  9.62
##
## Test for Heterogeneity:
## Q(df = 35) = 237.6958, p-val < .0001
##
```

```
## Model Results:
##
## estimate      se    zval    pval   ci.lb   ci.ub
##   0.3288  0.0988  3.3281  0.0009  0.1352  0.5224  ***
##
## ---
## Signif. codes:  0 '***' 0.001 '**' 0.01 '*' 0.05 '.' 0.1 ' ' 1
```

### Measure type: odds ratio

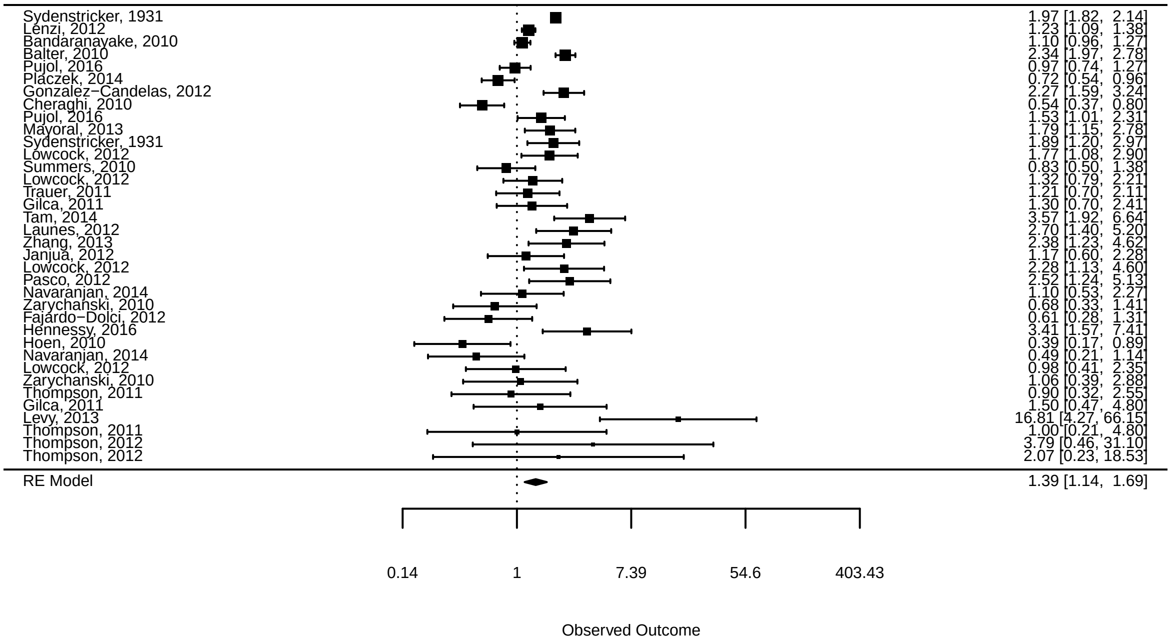
